# Exploring the causal effect of maternal pregnancy adiposity on offspring adiposity: Mendelian randomization using polygenic risk scores

**DOI:** 10.1101/2021.04.01.21251414

**Authors:** Tom A Bond, Rebecca C Richmond, Ville Karhunen, Gabriel Cuellar-Partida, Maria Carolina Borges, Verena Zuber, Alexessander Couto Alves, Dan Mason, Tiffany C Yang, Marc J Gunter, Abbas Dehghan, Ioanna Tzoulaki, Sylvain Sebert, David M Evans, Alex M Lewin, Paul F O’Reilly, Deborah A Lawlor, Marjo-Riitta Järvelin

## Abstract

**Background:** Greater maternal adiposity before or during pregnancy is associated with greater offspring adiposity throughout childhood, but the extent to which this is due to causal intrauterine or periconceptional mechanisms remains unclear. Here we use Mendelian Randomization (MR) with polygenic risk scores (PRS) to investigate whether associations between maternal pre-/early pregnancy body mass index (BMI) and offspring adiposity from birth to adolescence are causal.

**Methods:** We undertook confounder adjusted multivariable (MV) regression and MR using mother-offspring pairs from two UK cohorts: Avon Longitudinal Study of Parents and Children (ALSPAC) and Born in Bradford (BiB). In ALSPAC and BiB the outcomes were birthweight (BW; *N* = 9339) and BMI at age 1 and 4 years (*N* = 8659 to 7575). In ALSPAC only we investigated BMI at 10 and 15 years (*N* = 4476 to 4112) and dual-energy X-ray absorptiometry (DXA) determined fat mass index (FMI) from age 10–18 years (*N* = 2659 to 3855). We compared MR results from several PRS, calculated from maternal non-transmitted alleles at between 29 and 80,939 single nucleotide polymorphisms (SNPs).

**Results:** MV and MR consistently showed a positive association between maternal BMI and BW, supporting a moderate causal effect. For adiposity at most older ages, although MV estimates indicated a strong positive association, MR estimates did not support a causal effect. For the PRS with few SNPs, MR estimates were statistically consistent with the null, but had wide confidence intervals so were often also statistically consistent with the MV estimates. In contrast, the largest PRS yielded MR estimates with narrower confidence intervals, providing strong evidence that the true causal effect on adolescent adiposity is smaller than the MV estimates (*P*_difference_ = 0.001 for 15 year BMI). This suggests that the MV estimates are affected by residual confounding, therefore do not provide an accurate indication of the causal effect size.

**Conclusions:** Our results suggest that higher maternal pre-/early-pregnancy BMI is not a key driver of higher adiposity in the next generation. Thus, they support interventions that target the whole population for reducing overweight and obesity, rather than a specific focus on women of reproductive age.

## Background

It has been hypothesised that prenatal exposure to greater maternal adiposity during or prior to pregnancy causes greater adiposity in the offspring throughout life, via intrauterine effects or periconceptional mechanisms (for example effects on the oocyte) (1-4). There are well replicated observational associations between maternal body mass index (BMI) before or during pregnancy and offspring adiposity and cardiometabolic outcomes in childhood, adolescence and adulthood (5-8). Furthermore, evidence from animal experiments suggests that such associations are plausibly due to causal biological effects in the intrauterine period (9, 10). If true, this could have important implications for obesity prevention policy, because interventions to reduce maternal obesity before pregnancy might reduce offspring obesity risk in later life (1, 2, 6).

Triangulated epidemiological evidence from different study designs (11) suggests that associations between maternal BMI and offspring childhood/adolescent adiposity may *not* reflect a causal effect. For example, negative paternal exposure control studies (12-18) and studies examining associations within sibling groups (19, 20) suggest that confounding by genetic and/or environmental factors shared within families may be an important explanation for the associations. In addition, two Mendelian randomization (MR) (21, 22) studies, which used genetic variants as instrumental variables (IVs) for maternal BMI, provided no strong evidence for a causal effect (14, 23). However, in order to avoid bias due to genetic inheritance the primary analysis in the most recent MR study (23) was adjusted for an offspring weighted allele score, and simulations suggest that the use of a *weighted* allele score may not be the optimal approach to avoid bias (Personal communication, Wang G, Warrington N, Evans DM, 2020). In addition, both previous studies (14, 23) were unable to adjust for paternal genetic variants, which may be necessary to avoid collider bias (24). Furthermore, the causal estimates from previous MR studies were imprecise (14, 23, 24). For example, in the largest study (*N* = 6057) a one standard deviation (SD) higher maternal BMI was associated with a 0.05 SD increase in mean offspring BMI at age 7, but the 95% confidence interval was consistent with a 0.11 SD reduction or a 0.21 SD increase (23). If a positive causal effect is present this could have important public health implications, because it could lead to an accelerating intergenerational cycle of obesity that is difficult to break (1, 25). It is therefore important to conduct further MR investigations with improved methods, in order to obtain more precise estimates that are not subject to the aforementioned biases.

We aimed to use maternal non-transmitted allele polygenic risk scores (PRS) as IVs in a one-sample MR design, to explore the causal effect of maternal BMI on offspring adiposity from birth to adolescence, and to compare those results with confounder adjusted multivariable (MV) regression estimates. Because we used only maternal alleles that were not inherited by the offspring, we did not need to adjust for offspring or paternal genotype and thereby avoided biases that may have affected previous studies. We included thousands of genetic variants (hereafter referred to as single nucleotide polymorphisms [SNPs]) in the PRS, affording increased precision over previous MR studies which used only genome wide significant (GWS; *P* <5e-8) SNPs (14, 23). Based on previous MR studies (23, 26, 27) we hypothesised that greater maternal BMI would cause increased offspring birth weight (BW), but that the causal effect would attenuate over childhood and adolescence.

## Methods

### Study design

We have followed the MR-STROBE reporting guidelines in this paper (28). We conducted one-sample MR and compared these results with confounder adjusted multivariable (MV) regression analyses. We analysed data from two British population based prospective birth cohorts: the Avon Longitudinal Study of Parents and Children (ALSPAC) and Born in Bradford (BiB). These cohorts are described in **Additional file 1: Supplementary information S1** (14, 22-24, 29-75) and details of the study methodology have been reported previously (29-31).

### Selection of participants

Full details of sample selection for each cohort are given in **Additional file 1: Supplementary information S2**, and selection flow charts are presented in **Additional file 1: Supplementary information S3**. We included live-born singletons with non-missing data for the variables required for MR analyses, and excluded one offspring from any sibling groups present (chosen at random in ALSPAC or to maximise the sample size with data available in BiB). As the effects we were exploring may differ by ethnicity (32) we limited analyses to two ethnic groups: White European and South Asian, which comprised 40% and 51% of the sample with offspring genotype data available respectively. There were very few participants from other ethnic groups in either cohort. ALSPAC (93% White European) contributed only to the analyses in White Europeans and we meta-analysed these results with those from models fitted separately for BiB South Asians and BiB White Europeans. Derivation of ethnicity variables is described in **Additional file 1: Supplementary information S4**. The overall sample size for MR analyses ranged from 2659–5085 for ALSPAC, 1566–2262 for BiB South Asians and 1339–1992 for BiB White Europeans. The sample sizes for confounder adjusted MV estimates were somewhat smaller due to missing confounder data (1884–3265 for ALSPAC, 325–449 for BiB South Asians and 442–604 for BiB White Europeans). To enable comparison between the confounder adjusted MV estimates from models that adjusted for different covariates, we fitted all the models for each outcome using an identical sample with non-missing data for all relevant variables.

### Parental anthropometric variables

In ALSPAC, maternal pre-pregnancy weight and height were retrospectively reported by the women during pregnancy (at a mean gestational age of 24.7 weeks [SD 6.3]) or postnatally for 11.2% of mothers (at a mean of 22.0 weeks after birth [SD 6.7]). The reported weights correlated highly with weight recorded at the first antenatal clinic (Pearson correlation coefficient = 0.96). Paternal height and weight were reported by the fathers during their partner’s pregnancy (or postnatally for a minority of fathers). In BiB, early pregnancy BMI was calculated from height reported by the mothers at recruitment (26–28 weeks gestation) and weight extracted from the first antenatal clinic records (median 12 weeks gestation). Paternal height and weight were reported by the fathers at recruitment, which for the majority of fathers was at the time of their partner’s pregnancy.

### Offspring anthropometric variables

Offspring outcomes included BW and BMI at age 1 and 4 years (in ALSPAC and BiB), BMI at age 10 and 15 years (ALSPAC only) and fat mass index (FMI) at age 10, 12, 14, 16 and 18 years (ALSPAC only). The assessment of these outcomes is described in **Additional file 1: Supplementary information S5** and **Additional file 1: Supplementary information S6**, and included extraction of measurements from routine data sources (birth records/notifications, child health records, primary care records and school nurse records), clinical measurement by research staff or UK Government National Child Measurement Programme (NCMP) staff, and maternal/offspring questionnaire responses. In ALSPAC, we calculated FMI as fat mass (kg) / height (m)^2^ using fat mass measured by whole body dual-energy X-ray absorptiometry (DXA) (**Additional file 1: Supplementary information S5**).

### Anthropometric variable standardisation

In each of the three samples (ALSPAC, BiB White Europeans and BiB South Asians) we internally standardised exposure and outcome variables to give measures in standard deviation (SD) units. We standardised maternal BMI by maternal age (at delivery), in one year age categories. We standardised offspring BW by sex, and offspring BMI and FMI by sex and age (in one month categories).

### Confounder adjusted multivariable regression

We considered the following variables to be potential confounders: maternal age (which was adjusted for in the standardised exposure by calculating *z*-scores within maternal age strata), parity, maternal smoking during pregnancy, parental occupation, maternal educational attainment, paternal educational attainment and paternal BMI. Standard protocols for assessing these variables were used in each cohort, and full details are provided in **Additional file 1: Supplementary information S7**. We fitted three MV regression models: in model one we adjusted for maternal age, offspring age and offspring sex, in model two we additionally adjusted for the potential confounders listed above except for paternal BMI, and in model three (which was the main multivariable model of interest and is presented in **Results**) we additionally adjusted for paternal BMI. We took a complete case approach and excluded individuals with any missing data, therefore models one to three were fitted using identical samples. In sensitivity analyses we adjusted all models for gestational age at delivery, and for 20 genetic principal components (PCs) which we calculated from genome-wide SNPs separately for each of the three samples (**Additional file 1: Supplementary information S8**), in order to adjust for ancestry. In BiB we had to exclude a large number of individuals from the main MV models due to missing paternal BMI data. We therefore refitted models one and two without first excluding individuals with missing paternal BMI data (i.e. on larger samples), in order to explore potential selection bias.

### Genotyping, quality control and imputation

Mothers, offspring and (in ALSPAC only) fathers were genotyped using genome-wide arrays, followed by standard quality control (QC) measures (**Additional file 1: Supplementary information S9**). Array genotypes were then imputed to the Haplotype Reference Consortium (HRC), 1000 Genomes or UK10K reference panels (46-48) (**Additional file 1: Supplementary information S9**). In order to maximize the sample size we did not exclude cryptically related individuals for the primary analyses. As a sensitivity analysis we removed cryptic relatedness at a level corresponding to first cousins (dropping 6.7%, 13.5% and 9.1% of individuals in ALSPAC, BiB South Asians and BiB White Europeans respectively) by applying a KING (48) kinship coefficient threshold of 0.044 to the offspring using the PLINK software package version 2.00 (49, 50).

### Inference of maternal non-transmitted alleles

Our MR analyses used maternal PRS as IVs for maternal pre-pregnancy BMI. MR assumes that the IV is only associated with the outcome via its association with the exposure (**Additional file 1: Supplementary information S10**). For this to be true, the maternal PRS must be independent of the offspring’s genotype, but due to genetic inheritance this is not the case for PRS calculated in the usual way from all maternal alleles. We therefore calculated maternal PRS from only those maternal alleles that were not inherited by the offspring (maternal non-transmitted alleles (34)). After conversion of imputed genotypes to hard calls (integer valued allele dosages) and application of QC filters (**Additional file 1: Supplementary information S9**), we phased offspring imputed SNPs (for the sample of genotyped mother-offspring duos) using the duoHMM method implemented in the SHAPEIT v2 (r904) software package, with a window size of 5 Mb as per the authors recommendations for parent-offspring duos (76). This yielded maternal transmitted alleles (i.e. maternal alleles that were inherited by the offspring), which we used (along with the maternal genotypes) to infer the maternal non-transmitted alleles, from which we calculated maternal PRS, having first estimated SNP weights using maternal genotypes (see below).

### Polygenic risk score (PRS) calculation

Previous MR studies followed the widely used practice of using up to 97 GWS (*P* <5e-8) SNPs, but for polygenic traits such as BMI it is known that substantially improved phenotypic prediction can be achieved by including many more SNPs in the genetic risk score (i.e. more weakly associated SNPs that individually are not GWS) (61, 77-79). In order to maximise statistical power we used thousands of genome-wide SNPs to calculate a BMI PRS, as a weighted sum of BMI-increasing maternal non-transmitted alleles at SNPs across the genome. We tested four PRS methods (clumping and thresholding (80), LDPred (52), lassosum (53, 81) and the BOLT-LMM linear predictor (54)) (**Additional file 1: Supplementary information S11** provides further information for each of these). Of these four methods, lassosum explained the highest proportion of variance (*R*^2^) for maternal BMI in both ALSPAC and BiB (which we refer to as the target datasets), therefore we used the lassosum PRS for subsequent MR analyses. Lassosum requires summary statistics from a genome wide association study (GWAS), which we refer to as the base dataset. We conducted a GWAS in the UK Biobank (UKB), a prospective cohort of 502,628 volunteers (with 5% response rate of those invited), recruited from across the UK at age 40–69 years between 2006 and 2010 (58, 82) (**Additional file 1: Supplementary information S11**). In order to avoid overfitting due to overlap between the base and target samples we excluded attendees of the Bristol (where ALSPAC participants would have attended) or Leeds (where Born in Bradford participants would have attended) UKB assessment centres. We meta-analysed the summary statistics from the UKB GWAS with a published BMI GWAS from the GIANT consortium (60, 83), giving a total base sample size of up to 756,048. We applied the lassosum algorithm to the meta-analysed base dataset; lassosum uses penalised regression to carry out shrinkage and selection on the base GWAS SNP effects and accounts for LD information from a reference panel. We used the ALSPAC or BiB datasets as the reference panels as per the authors’ recommendations) (53). PLINK was used to calculate the PRS for ALSPAC and BiB individuals using the lassosum SNP weights for around 80,000 SNPs (see **Table 2** for the exact number of SNPs for each cohort). We also calculated three PRS from fewer SNPs, to be used in sensitivity analyses to explore potential pleiotropic effects (we would expect that the risk of pleiotropic bias might decrease as fewer SNPs are included in the IV; see below). These PRS used (i) around 30 GWS SNPs identified in a 2010 BMI GWAS (67), (ii) around 90 GWS SNPs identified in a 2015 BMI GWAS (60), and (iii) around 500 GWS SNPs identified as primary signals in a 2018 BMI GWAS (61). Full details of these analyses, including the exact number of SNPs used to calculate each PRS (which varied between samples) are given in **Additional file 1: Supplementary information S12**.

**Table 1:** Characteristics of the mothers and offspring in ALSPAC and BiB

|  | ALSPAC |  |  |  | BiB (WE) |  |  |  | BiB (SA) |  |  |  |
| --- | --- | --- | --- | --- | --- | --- | --- | --- | --- | --- | --- | --- |
|  | Mean | SD | N | Female offspring (%) | Mean | SD | N | Female offspring (%) | Mean | SD | N | Female offspring (%) |
| Maternal BMI (kg/m<sup>2</sup>) | 23.0 | 3.8 |  |  | 26.9 | 6.0 |  |  | 25.8 | 5.5 |  |  |
| Maternal age (years) | 29.3 | 4.5 |  |  | 27.4 | 6.0 |  |  | 28.8 | 5.0 |  |  |
| Birth weight (z score) | 0.10 | 0.95 | 5085 | 50.5 | -0.05 | 0.97 | 1992 | 47.9 | -0.56 | 0.92 | 2262 | 47.9 |
| Gestational age (weeks) | 39.6 | 1.7 |  |  | 39.4 | 1.6 |  |  | 39.1 | 1.5 |  |  |
| 1yr weight for length percentile* | 0.64 | 0.27 | 4838 | 50.6 | 0.61 | 0.28 | 1798 | 47.8 | 0.48 | 0.30 | 2023 | 48.1 |
| Age at measurement (years) | 0.9 | 0.2 |  |  | 0.8 | 0.3 |  |  | 0.8 | 0.3 |  |  |
| 4yr BMI (z score) | 0.46 | 1.05 | 4670 | 50.2 | 0.50 | 0.97 | 1339 | 48.6 | 0.10 | 1.21 | 1566 | 48.5 |
| Age at measurement (years) | 4.1 | 0.7 |  |  | 4.5 | 0.7 |  |  | 4.5 | 0.7 |  |  |
| 10yr BMI (z score) | 0.26 | 1.12 | 4476 | 51.3 |  |  |  |  |  |  |  |  |
| Age at measurement (years) | 9.9 | 0.5 |  |  |  |  |  |  |  |  |  |  |
| 15yr BMI (z score) | 0.30 | 1.11 | 4112 | 51.7 |  |  |  |  |  |  |  |  |
| Age at measurement (years) | 15.0 | 0.8 |  |  |  |  |  |  |  |  |  |  |

**SA:** South Asians, **WE:** White Europeans, **SD:** standard deviation, **z score:** sex- and gestational age-adjusted standard deviation score (UK-WHO Growth Reference (88)), \*Sex-adjusted percentile calculated using the WHO Child Growth Standards (89). All z scores and percentiles were calculated via the *zanthro* Stata package (90). Data for absolute values (as opposed to z scores) are presented in **Additional file 1: Supplementary information S18**

**Table 2:**
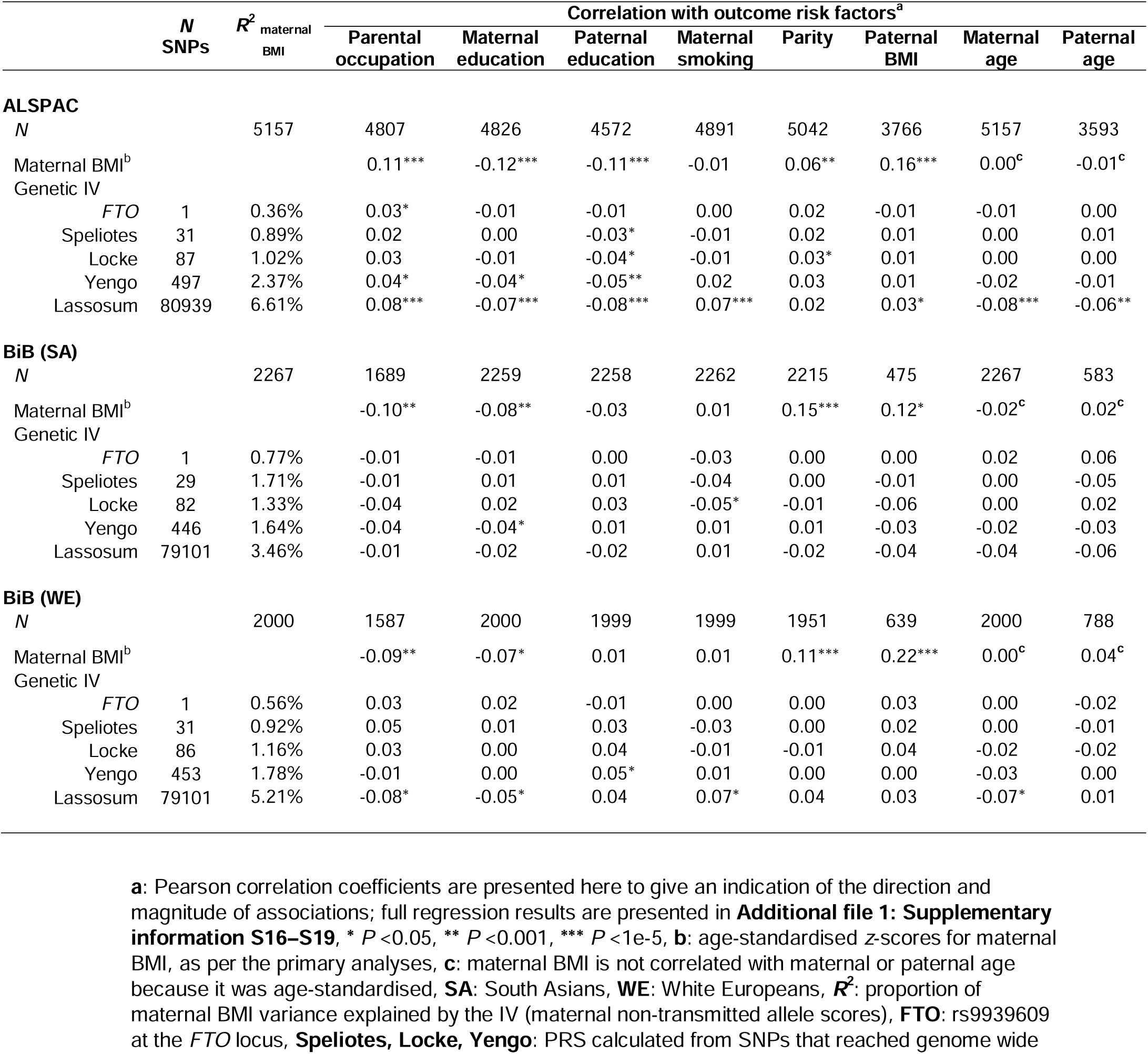
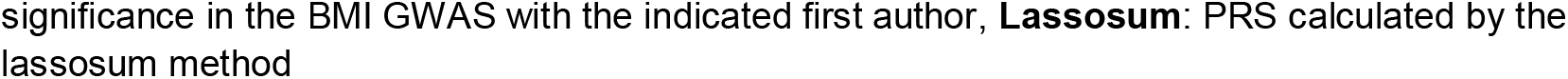
Associations of maternal BMI with outcome risk factors, and of the genetic IVs (maternal non-transmitted alleles) with maternal BMI and outcome risk factors

### Mendelian randomization

For the primary MR analyses we used the lassosum non-transmitted allele BMI PRS as an IV for maternal BMI and fitted models using the two-stage least squares (TSLS) method (22) (i.e. one sample MR). **Additional file 1: Supplementary information S10** shows our MR analyses diagrammatically. We included 20 genetic PCs as covariates in order to adjust for population stratification. We tested for a difference between the most extensively confounder adjusted MV estimates (model three) and MR estimates using a *z*-test (**Additional file 1: Supplementary information S13**), and used a bootstrapping procedure to estimate the covariance between MV and MR estimates in order to calculate the *z*-statistic. Evidence for a difference between the two could reflect residual confounding in the MV analyses or violation of one or more of the MR assumptions.

### Meta-analysis

We examined the point estimates, *I*^2^ statistics and Cochran’s *Q* test P-values for the MV and MR associations and found little evidence for heterogeneity between ALSPAC, BiB South Asians and BiB White Europeans (**Additional file 1: Supplementary information S14**). We therefore meta-analysed estimates from the three samples using a fixed effects model. Results were similar when we instead used a random effects model. For the meta-analyses we used the ratio estimator (calculated as the meta-analysed PRS-outcome regression coefficient 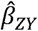 divided by the meta-analysed PRS-exposure regression coefficient 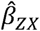; in the present study both coefficients were estimated in the same sample) which gives equivalent results to TSLS (84). We calculated the standard errors for the pooled MR estimates using a Taylor series approximation (70).

### Checking MR assumptions

We checked the assumptions made by MR analyses (**Additional file 1: Supplementary information S10**); if these assumptions are met then our MR estimates can be interpreted as causal effect estimates (22). We first assessed whether the PRS were associated with maternal BMI using the *R*^2^ and *F*-statistics. Next, we explored whether the PRS-outcome associations were confounded by ancestry (population stratification) using a linear mixed model (LMM). LMMs have been widely used in GWAS to adjust for population stratification and cryptic relatedness (71). We fitted models for the numerator and denominator of the ratio estimator separately, using the *--reml-est-fix* command in the GCTA software package (version 1.91.7beta) (43). Further details of the LMM approach are given in **Additional file 1: Supplementary information S15**. Finally, we conducted several analyses to explore whether the maternal PRS influences offspring adiposity via mechanisms other than intrauterine or periconceptional exposure to increased maternal BMI (horizontal pleiotropy). We first tested for associations of the PRS with other potential risk factors for the offspring outcomes (85). We would expect that the risk of pleiotropic bias might decrease as fewer SNPs are included in the IV. We therefore repeated MR analyses with IVs calculated from a single BMI-associated SNP (rs9939609 at the *FTO* locus, the locus at which there is currently the strongest evidence for association with BMI (61)), as well as the three PRS calculated from only strongly BMI-associated (GWS) SNPs, as described above. Furthermore, most of the SNPs included in the lassosum BMI PRS had small effect sizes, and the consequences of this for the extent of horizontal pleiotropic effects are unclear (72), so we explored how MR estimates varied with varying SNP effect size distributions. We also tested for evidence of between-SNP MR estimate heterogeneity (Cochran’s Q test) and used MR Egger regression (74) to investigate horizontal pleiotropy, for the analyses based on GWS SNPs. Finally, to investigate collider bias and bias due to assortative mating we examined the association between the maternal and paternal lassosum BMI PRS, in the subset of ALSPAC participants with paternal genotype data available (*N* = 1325).

### Other sensitivity analyses

We explored departure from linearity of the MV and MR associations by examining augmented partial residual plots with overlaid linear regression lines and nonparametric loess smoothers (86). The residuals from several models involving adolescent BMI and FMI variables were somewhat positively skewed so we repeated MV and MR analyses using the natural log of the relevant variables. We examined whether results differed for BW, BMI and ponderal index (weight [kg] / length [m]^3^) at birth (in ALSPAC only as birth length was not available in BiB). Finally, we tested for interaction by offspring sex for the MV and MR models. We carried out statistical analyses in R version 3.5.1 (87), and Stata version 13.1 (StataCorp, College Station, TX, USA).

## Results

### Participant characteristics

**Table 1** shows the participant characteristics. The prevalence of maternal obesity (maternal BMI ≥30) was 5.5% (95% confidence interval [CI]: 4.9%, 6.1%) in ALSPAC and markedly higher in BiB South Asians (20.5% [95% CI: 18.9%, 22.2%]) and BiB White Europeans (26.0% [95% CI: 24.1%, 28.0%]). The samples for our analyses were smaller than those for the full cohorts at birth due to missing data, particularly for the MV associations. Despite this there were not large differences in the distributions of BW, maternal BMI or offspring sex between the baseline samples and those from which we calculated the MV estimates (**Additional file 1: Supplementary information S16**). Furthermore, when we fitted MV models one and two on a larger sample (retaining individuals with missing paternal BMI) there were not large differences in the primary MV results (**Additional file 1: Supplementary information S17**).

### Associations of genetic IVs with maternal BMI and offspring genotype

As we included more SNPs in the IV the *R*^2^ for maternal BMI increased markedly, from <1% for the *FTO* IV to ∼3–7% for the lassosum IV (**Table 2**). First-stage *F*-statistics were >75 for all lassosum MR models (**Additional file 1: Supplementary information S19**). The lassosum maternal non-transmitted allele BMI PRS was not correlated with the offspring’s PRS (results available from the authors on request).

### Associations of maternal BMI with confounders/outcome risk factors

There was strong evidence in all three samples for associations between maternal BMI and several other potential risk factors for the offspring outcomes, including parental occupation, educational attainment, maternal parity and paternal BMI (results are summarised in **Table 2**, and full regression results including the direction of associations are given in **Additional file 1: Supplementary information S20–S23**).

### Associations of maternal BMI PRS with confounders/outcome risk factors

Genetic IVs based on fewer SNPs (i.e. <100 SNPs) were generally not associated with the outcome risk factors. In ALSPAC however there was strong evidence for association of the lassosum IV (based on 80,939 SNPs) with parental occupation, parental educational attainment, parental age, maternal smoking and paternal BMI. These associations were mostly present for BiB White Europeans but absent for BiB South Asians.

### Confounder adjusted MV regression

In confounder adjusted MV regression models maternal BMI was positively associated with all offspring outcomes (**Figure 1, Additional file 1: Supplementary information S24**; meta-analysis heterogeneity statistics are given in **Additional file 1: Supplementary information S14**). Estimates for the SD scale increase in offspring outcomes associated with a 1 SD higher age-adjusted maternal BMI ranged from 0.07 (95% CI: 0.04, 0.10) for 4 year BMI to 0.32 (95% CI: 0.29, 0.36) for 15 year BMI, and MV estimates for 10–18 year FMI were similar to those for 15 year BMI. Adjustment for potential confounders had a negligible impact on the estimates, aside from a small attenuation on adjustment for paternal BMI for outcomes after birth. Results were similar when we refitted MV models one and two on larger samples without excluding individuals with missing paternal BMI data (**Additional file 1: Supplementary information S17**). Additional adjustment for gestational age at birth or 20 genetic PCs had a negligible effect (**Additional file 1: Supplementary information S25, S26**), and there was not a large difference when BMI or ponderal index at birth was substituted for BW in ALSPAC (**Additional file 1: Supplementary information S27**).

**Figure 1:**
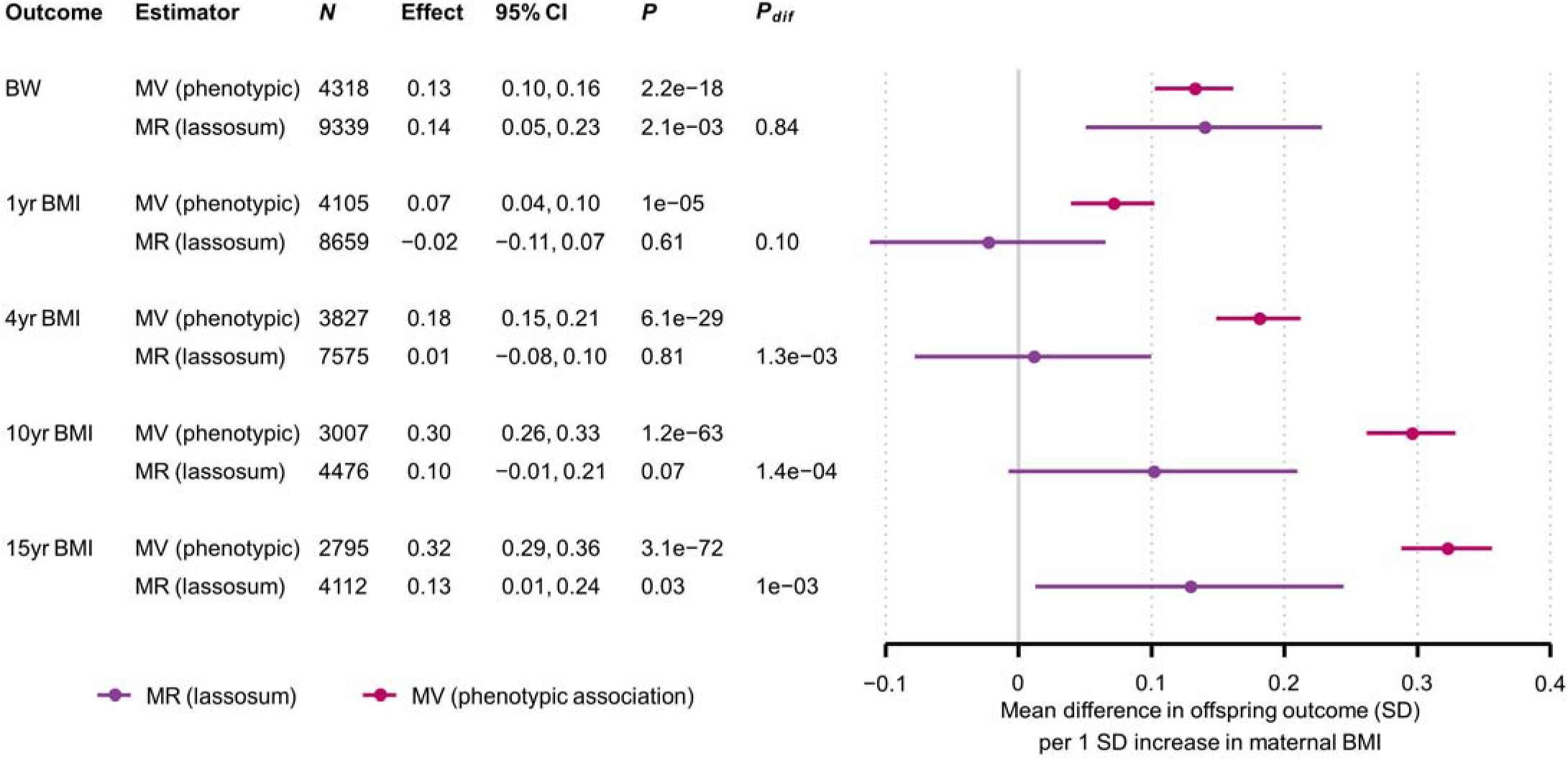
Mean difference in offspring BW and BMI (SD) per 1SD increase in maternal BMI, from MR (lassosum) and confounder adjusted multivariable regression (MV) models Confounder adjusted multivariable regression (MV) estimates are from model three (**Methods**). ***N***: Number of participants. The number of SNPs used for the MR analyses is provided separately by cohort in **Table 1. *P***: *P*-value for the null hypothesis that the effect equals zero, ***P***_**dif**_: *P*-value for the null hypothesis that MR effect equals the MV effect.

### MR results

For BW the MR estimate for the lassosum PRS for all three samples meta-analysed was 0.14 (0.05, 0.23), which was similar to the MV estimate; *P*_difference (MV vs. MR)_ = 0.84)) (**Figure 1**). The corresponding lassosum MR estimates for 1 year BMI and 4 year BMI were -0.02 (−0.11, 0.07) and 0.01 (−0.08, 0.10) respectively, and there was moderate to strong evidence for an MR-MV difference (*P*_difference_ = 0.10 and 1.3e-3 respectively). The MR estimates for 10 and 15 year BMI in ALSPAC (0.10 [-0.01, 0.21] and 0.13 [0.01, 0.24] respectively) were also smaller than the MV estimates (*P*_difference_ = 1.4e-4 and 1.0e-3 respectively). Results for adolescent FMI (**Figure 2**) were similar to those for adolescent BMI: MR estimates ranged between 0.09 and 0.19, and there was strong evidence that the MR estimates were smaller than the MV estimates, with *P*_difference_ ranging between 0.05 and 4.7e-4). We did not observe strong evidence for non-linearity or interaction by sex for either the MV or MR models (results available from the authors), and results were similar when we (i) substituted BMI or ponderal index at birth for BW (**Additional file 1: Supplementary information S28**), (ii) natural log transformed skewed variables (results available from the authors), (iii) removed cryptic relatedness from the sample (results available from the authors), and (iv) used linear mixed models to adjust for population structure (**Additional file 1: Supplementary information S29**). In linear regression models (as opposed to two-stage least squares regression) there was strong to moderate evidence that the lassosum maternal non-transmitted allele BMI PRS was associated with offspring BW and adolescent adiposity (**Additional file 1: Supplementary information S30, S31**).

**Figure 2:**
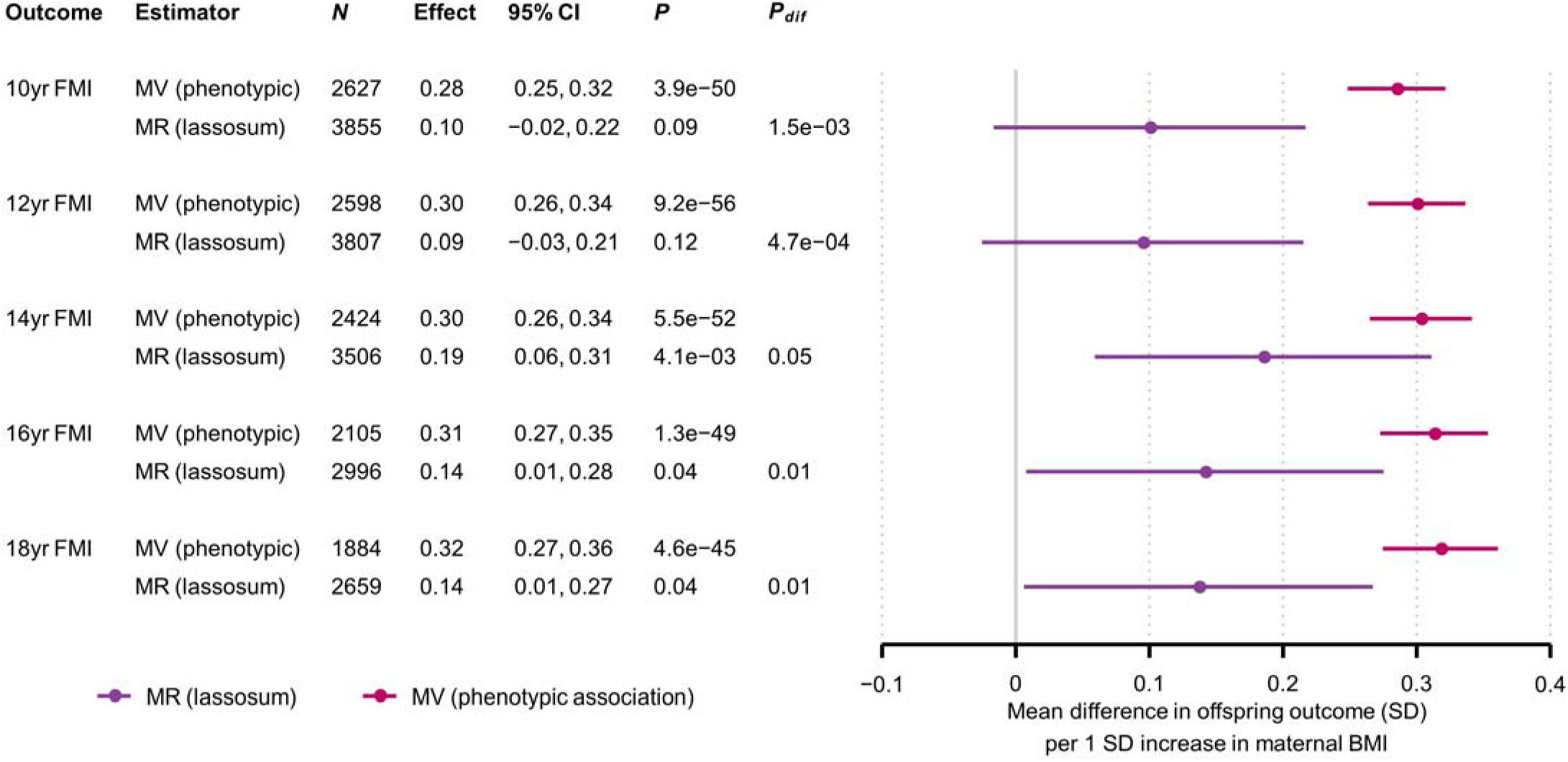
Mean difference in offspring FMI (SD) per 1SD increase in maternal BMI, from MR (lassosum) and confounder adjusted multivariable regression (MV) models Confounder adjusted multivariable regression (MV) estimates are from model three (**Methods**). ***N***: Number of participants. The number of SNPs used for the MR analyses is provided separately by cohort in **Table 1. *P***: *P*-value for the null hypothesis that the effect equals zero, ***P***_**dif**_: *P*-value for the null hypothesis that MR effect equals the MV effect.

### MR estimates for IVs with fewer SNPs

When we replaced the lassosum PRS with alternative IVs calculated from fewer SNPs, our MR estimates varied in a manner that was specific to the offspring outcome (**Figure 3, Figure 4**). For BW, including fewer SNPs in the IV did not result in large differences in the MR estimates, although the precision reduced markedly as we used fewer SNPs. For 1 and 4 year BMI, MR estimates increased as we used fewer SNPs, whereas for 10 year BMI they largely remained stable and for 15 year BMI they decreased. The patterns for adolescent FMI were similar to those for adolescent BMI. For outcomes apart from BW and 1yr BMI, including more SNPs in the IV generally resulted in stronger evidence that MR estimates differed from MV estimates (i.e. smaller *P*_dif_).

**Figure 3:**
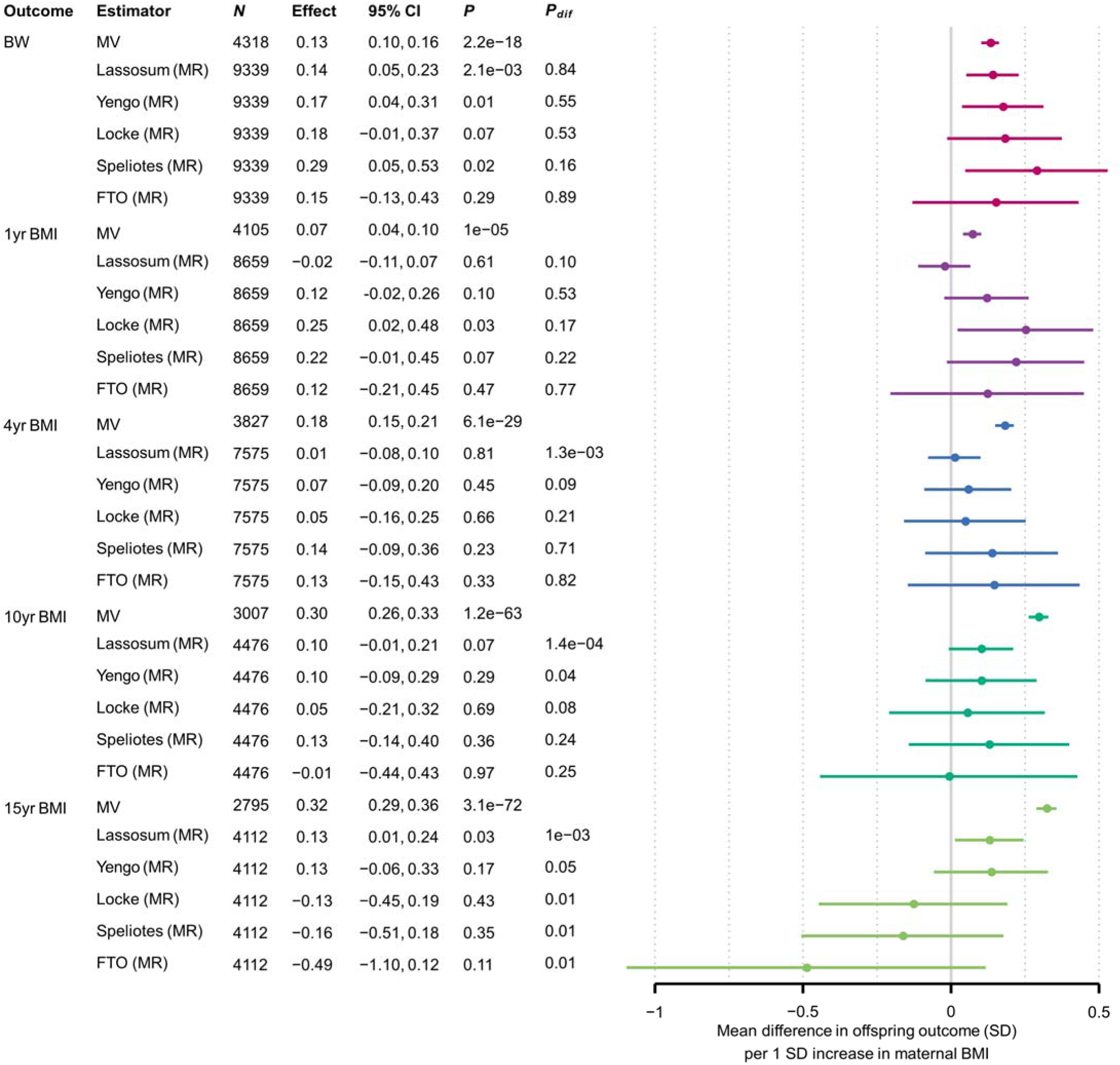
Mean difference in offspring BW and BMI (SD) per 1SD increase in maternal BMI, from MR models using different SNP sets and confounder adjusted multivariable regression (MV) models Confounder adjusted multivariable regression (MV) estimates are from model three (**Methods**). ***N***: Number of participants. The number of SNPs used for the MR analyses is provided separately by cohort in **Table 1. *P***: *P*-value for the null hypothesis that the effect equals zero, ***P***_**dif**_: *P*-value for the null hypothesis that MR effect equals the MV effect, **FTO**: rs9939609 at the *FTO* locus, **Speliotes, Locke, Yengo**: GWS SNPs from the GWAS with the indicated first author, **Lassosum**: PRS calculated by the lassosum method. Colours denote outcomes

**Figure 4:**
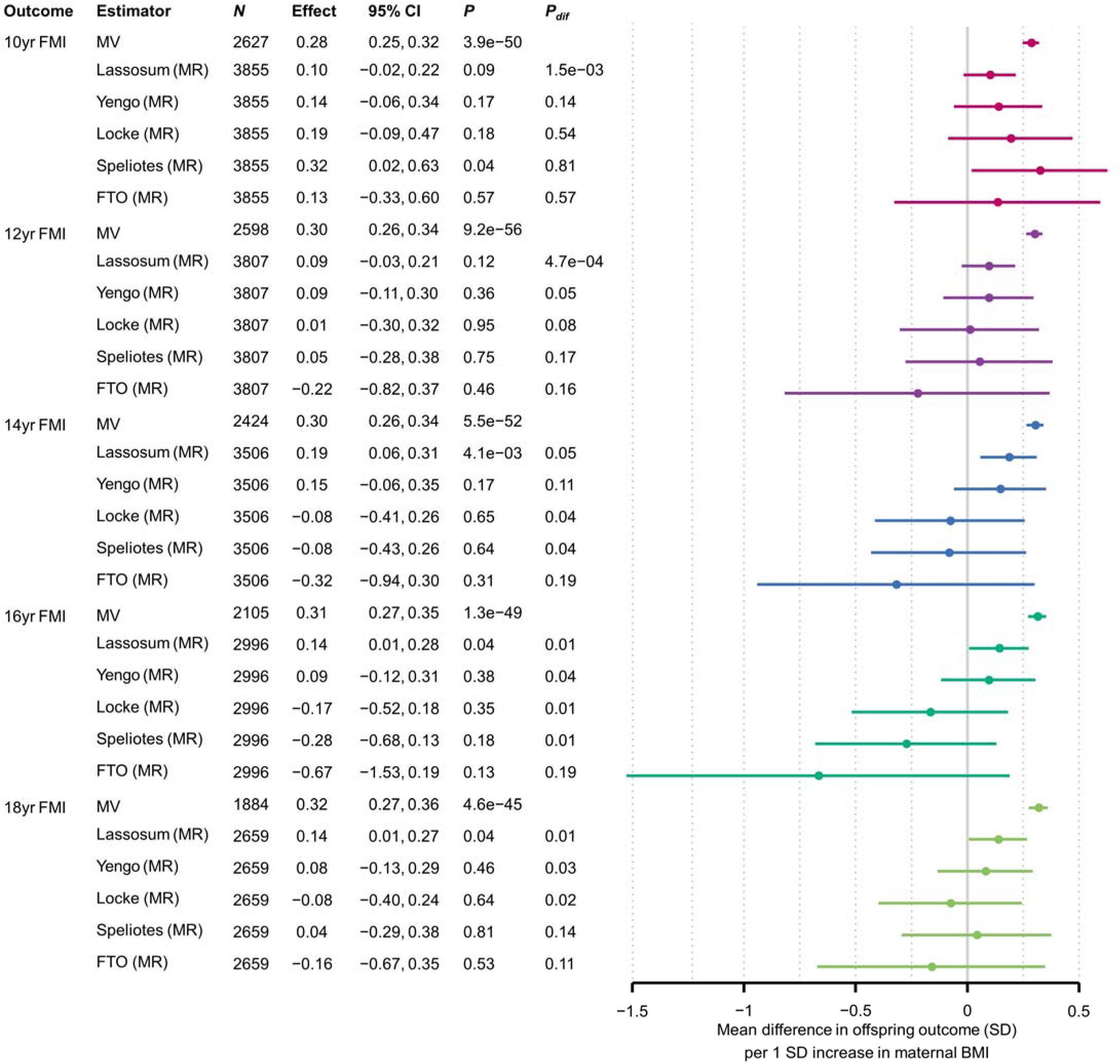
Mean difference in offspring FMI (SD) per 1SD increase in maternal BMI, from MR models using different SNP sets and confounder adjusted multivariable regression (MV) models Confounder adjusted multivariable regression (MV) estimates are from model three (**Methods**). ***N***: Number of participants. The number of SNPs used for the MR analyses is provided separately by cohort in **Table 1. *P***: *P*-value for the null hypothesis that the effect equals zero, ***P***_**dif**_: *P*-value for the null hypothesis that MR effect equals the MV effect, **FTO**: rs9939609 at the *FTO* locus, **Speliotes, Locke, Yengo**: GWS SNPs from the GWAS with the indicated first author, **Lassosum**: PRS calculated by the lassosum method. Colours denote outcomes

### MR estimates for SNPs with differing effect size distributions, between-SNP heterogeneity and MR Egger results

For the majority of outcomes (particularly in adolescence) there was moderate to strong statistical evidence that SNPs with smaller effect sizes gave larger (more positive) MR estimates (*P* = 4.0e-3, 2.6e-2, 1.8e-2 and 4.7e-4 for 15yr BMI, 14yr FMI, 16yr FMI and 18yr FMI respectively), and this was not driven by weak instrument bias (**Additional file 1: Supplementary information S32**). When using only large-effect (GWS) SNPs, in general (and in light of the 40 statistical tests carried out) there was not strong statistical evidence for between-SNP MR estimate heterogeneity (all Cochran’s *Q* test *P*-values ≥0.017), nor was there strong evidence that the MR-Egger intercept differed from zero (all MR-Egger intercept *P*-values ≥0.023) (**Additional file 1: Supplementary information S33**).

### Parental phenotypic and PRS correlations

In ALSPAC there was strong evidence for correlation between maternal and paternal BMI (Pearson’s *r*: 0.22, 95% CI: 0.16, 0.28, *P* = 7.9e-14), but no evidence for correlation between maternal non-transmitted allele and paternal lassosum BMI PRS (*r*: 0.02, 95% CI: -0.04, 0.07, *P* = 0.55). For comparison, a maternal lassosum BMI PRS that was calculated from both transmitted and non-transmitted alleles was slightly more strongly correlated with the paternal PRS (*r*: 0.04, 95% CI: -0.01, 0.10, *P* = 0.14).

## Discussion

We applied a Mendelian randomization (MR) approach using PRS calculated from maternal non-transmitted alleles, to explore the causality of associations between maternal pre-/early-pregnancy BMI and offspring birth weight (BW) and child/adolescent adiposity. For the association between maternal BMI and offspring BW, our MR and confounder adjusted multivariable regression (MV) estimates were similar. In contrast, for offspring adiposity outcomes beyond 1 year of age (including BMI and DXA-determined FMI) the MR estimates were weaker than the MV estimates. These results markedly strengthen the evidence that confounder adjusted observational associations between maternal BMI and offspring adolescent adiposity are subject to residual confounding. We found no strong evidence for a causal effect of maternal BMI on offspring adiposity beyond birth, although based on the present results we cannot rule out a small to moderate causal effect.

Our data build on two previous MR studies which investigated associations between maternal BMI and offspring child/adolescent adiposity (14, 23), and a methodological paper which presented a limited investigation of adiposity outcomes as an empirical example (24). Although the previous studies provided no strong evidence for a causal effect, they were limited by wide confidence intervals and/or potential biases (see **Strengths and Limitations** below). The present study overcame these limitations by using more powerful PRS and a maternal non-transmitted allele score approach. For the association between maternal BMI and offspring BW, our MR and MV estimates were highly concordant, in agreement with previous MR studies that supported a causal effect of greater maternal BMI on greater offspring BW (26, 27). We have previously shown that genetic confounding (i.e. confounding due to direct effects of maternal alleles inherited by the offspring) is unlikely to explain the association of maternal BMI with BW, but may potentially be important for the association with adolescent BMI (40). The present results are consistent with this, as well as with the balance of evidence from negative paternal exposure control studies (12-18) and within sibship analyses (19, 20), which suggests that familial confounding is an important explanation of the maternal BMI-offspring child/adolescent adiposity association. Studies that examined the effect of extreme maternal obesity using a pre- and post-bariatric surgery design (91-94) have small sample sizes and have not been entirely consistent, therefore do not provide strong evidence against this conclusion.

Although we found no strong evidence *for* a causal effect in late childhood/adolescence, we cannot rule out a small to moderate causal effect, due to the imprecision of our MR estimates. Indeed, the primary lassosum MR estimates were greater than zero for 15 year BMI and 14–18 year FMI. We do not interpret these as unbiased estimates for the causal effect of maternal BMI, because of the possibility of pleiotropic bias (see below). These results do suggest however that some maternal exposure(s) that are correlated with the maternal BMI PRS have a causal effect on offspring child/adolescent adiposity, although our analyses are unable to distinguish whether this is a pre or postnatal effect. Plausible mechanisms include intrauterine effects such as fetal overnutrition (1) and postnatal effects such as maternal influence on offspring eating behaviour (95), but other mechanisms have been hypothesised, including periconceptional effects (such as altered oocyte structure or function (3)). In linear regression analyses we found moderate to strong evidence for associations between the maternal non-transmitted allele BMI PRS and offspring adolescent adiposity (including BMI and DXA-determined FMI). These observed maternal genetic effects merit further investigation in other datasets, particularly as previous studies have not found evidence for parental genetic effects on BMI in childhood (96) or adulthood (97).

In ALSPAC and BiB White Europeans we observed associations between the maternal BMI PRS and potential confounders, including parental occupation, education, age and maternal smoking. These results invite careful consideration of which of the ever-increasing number of GWS associated BMI SNPs are likely (in combinations) to be the most valid instruments for MR studies, having taken account of all IV assumptions.

### Strengths and limitations

Our study has several key strengths. We studied two prospective birth cohorts with maternal and offspring genome-wide genotype data, maternal BMI measurements and offspring adiposity outcomes available, allowing us to conduct mother-offspring MR analyses. We used state-of-the-art methods to calculate a powerful PRS from around 80,000 SNPs. This yielded a substantial increase in statistical power over previous MR studies, which analysed similar ALSPAC datasets to ours, but employed either a single SNP in the *FTO* gene (14) or allele scores calculated from up to 97 SNPs (23, 24) (similar to the “Speliotes” and “Locke” IVs in the present analysis). Our primary lassosum PRS explained 3–7% of maternal BMI variance, compared to ∼1.5% for the strongest IVs used previously (power calculations are given in **Additional file 1: Supplementary information S34**).

Another strength over previous work is our use of maternal non-transmitted allele PRS, thereby avoiding the need to control for genetic inheritance by adjusting for offspring genotype. A previous methodological paper made use of this approach (24), but conducted a much more limited analysis of a far smaller subset of adiposity outcomes than that which we have explored here. Controlling for offspring genotype may be suboptimal for two distinct reasons: (i) it may introduce collider bias if paternal genotype influences the offspring outcome independently of offspring genotype (i.e. if paternal genetic effects exist) (24, 35), and (ii) if the investigator adjusts for a *weighted* allele score, this may introduce bias by inadequately blocking the genetic inheritance path (Personal communication, Wang G, Warrington N, Evans DM, 2020). Because these two biases may be in opposite directions, the net direction of any bias affecting the largest previous study (23) is uncertain. We acknowledge that our primary MR estimates may be affected by pleiotropic bias due to the large number of SNPs, many of which had small effect sizes, that we used to calculate the PRS. This possibility is also suggested by the associations that we observed between the lassosum BMI PRS and several potential confounders of the maternal BMI-offspring adiposity association. However, sensitivity analyses suggested that for most outcomes pleiotropic bias is likely to be away from zero, which would weaken the apparent evidence for an MR-MV difference (**Additional file 1: Supplementary information S32, S33**). Thus, our primary MR results are conservative, in that they may overstate the size of the causal effect (which we hypothesised to be zero). The fact that for 10 and 15 year BMI, using more SNPs yielded increased precision and stronger evidence for an MR-MV difference (**Figure 2**), despite the potential pleiotropic bias away from zero, illustrates the benefit of our approach.

We also conducted extensive sensitivity analyses to explore other potential biases in our results. When we used a linear mixed model (LMM) to adjust for population structure the results were similar to our primary estimates. We did not remove cryptic relatedness for our primary analyses, in order to maximise the sample size and because the LMM controls for bias due to cryptic relatedness (71). However, results were similar when we removed cryptic relatedness at a level corresponding to first cousins. Finally, we found no strong evidence that maternal and paternal lassosum BMI PRS were correlated, suggesting that our results are not importantly biased due to assortative mating.

We acknowledge several limitations of our study. First, although the results in BiB and ALSPAC were similar, replication in other cohorts with suitable data, and in particular with adolescent adiposity measures (which we could only examine in ALSPAC) would be valuable. A previous study meta-analysed data from ALSPAC and the Generation R cohort using 32 maternal SNPs (23), but we were unable to extend our approach to Generation R due to the unavailability of maternal genome-wide SNP data. **Additional file 1: Supplementary information S35** compares the present analysis to previous analyses of ALSPAC data. We have only studied UK participants. However, the similarity of findings between White European and South Asian BiB participants, and between BiB (a cohort with high levels of deprivation born during the obesity epidemic) and ALSPAC (more affluent than the UK average) suggest that our findings may be generalisable to other populations. Second, BMI (especially self-reported BMI) is an imperfect proxy measure for adiposity. However, it has been shown previously in ALSPAC that self-reported pre-pregnancy BMI is strongly correlated with BMI measured in early pregnancy (23), and that any misreporting does not markedly differ by mean weight (98). There is also evidence that the correlation with directly measured adiposity is strong for child and adult BMI (99, 100) and moderate for neonatal weight (101); furthermore, our results were similar for DXA derived FMI. Third, we assumed that causal relationships between exposures and outcomes were linear. Although our data provided no evidence for non-linearity, a slight plateauing of the observational association between maternal BMI and offspring child/adolescent BMI at higher maternal BMI levels was previously observed in a large meta-analysis (6). MR estimates such as ours, which assume linearity, nevertheless approximate the population-averaged causal effect (which is the average effect resulting from a unit increase in the exposure for all individuals in the population, regardless of their initial exposure level) (102). However, given the shape of the observational association (6) it is plausible that our MR estimates overstate the true causal effect for mothers with overweight/obesity. Finally, the samples used for some of our analyses (particularly for MV models) were smaller than the full samples at baseline due to missing data and loss to follow up, raising the possibility that our results are affected by selection bias. However, the distributions of maternal BMI, BW and offspring sex were similar for the samples used for our analyses and the samples at baseline, and MV results were similar when we refitted models on larger samples without excluding individuals with missing paternal BMI data. It therefore seems unlikely that selection bias would be of sufficient magnitude to alter our conclusions.

### Conclusion

We explored the causality of associations between maternal pre-/early-pregnancy BMI and offspring BW and child/adolescent adiposity (measured by BMI and DXA-determined FMI), using an MR approach with PRS calculated from maternal non-transmitted alleles. This approach yielded narrower confidence intervals compared with previous studies, and avoided sources of bias that may have affected previous work. We found no strong evidence for a causal effect of maternal BMI on offspring adiposity beyond birth, but strong evidence that confounder adjusted observational associations between maternal BMI and adolescent adiposity are affected by residual confounding. Although we cannot rule out a small or moderate causal effect on child/adolescent adiposity, the present study suggests that higher maternal pre-/early-pregnancy BMI is not a key driver of greater adiposity in the next generation. Thus, our results support interventions that target the whole population for reducing overweight and obesity, rather than a specific focus on women of reproductive age.

## Data Availability

The ALSPAC study website (http://www.bristol.ac.uk/alspac/researchers/our-data/) contains details of all the data that are available through a fully searchable data dictionary and variable search tool. Scientists are encouraged and able to use BiB data. Data requests are made to the BiB executive using the form available from the study website http://www.borninbradford.nhs.uk (please click on ‘Science and Research’ to access the form). Guidance for researchers and collaborators, the study protocol and the data collection schedule are all available via the website. All requests are carefully considered and accepted where possible. UK Biobank data are available from the UK Biobank (http://biobank.ndph.ox.ac.uk/showcase/).

## Abbreviations

ALSPAC: Avon Longitudinal Study of Parents and Children
BiB: Born in Bradford
BMI: body mass index
DXA: dual-energy X-ray absorptiometry
FMI: fat mass index
GWAS: genome wide association study
GWS: genome wide significant (*P* < 5e-8)
IV: instrumental variable
LMM: linear mixed model
MR: Mendelian randomization
MV: multivariable
NCMP: UK Government National Child Measurement Programme
PC: principal component
PRS: polygenic risk score(s)
QC: quality control
SD: standard deviation
TSLS: two-stage least squares
UKB: UK Biobank
SNP: single nucleotide polymorphism

## Declarations

### Ethics approval and consent to participate

For ALSPAC, ethical approval was obtained from the ALSPAC Ethics and Law Committee and the Local Research Ethics Committees. For BiB, ethical approval was obtained from Bradford National Health Service Ethics Committee (ref 06/Q1202/48). UK Biobank received ethical approval from the North West Multi-centre Research Ethics Committee (MREC) (ref 11/NW/0382). Informed consent was obtained from participants of all studies.

### Consent to publish

This manuscript does not include details, images, or videos relating to an individual person, therefore consent for publication is not required, beyond the informed consent provided by all study participants as described above..

### Author’s contributions

Conceptualization: TAB, MRJ, DAL

Methodology: DAL, VZ, PFO, MG, AL, SS, DME, MRJ

Software: TAB, RCR, VK, GCP

Formal analysis: TAB

Data curation: TAB, IT, AD, ACA, DM, TY, MCB

Writing- original draft preparation: TAB, MRJ, DAL, RCR, PFO

Writing- review and editing: TAB, MRJ, DAL, RCR, PFO, VZ, MG, AL, SS, IT, AD, DM, TY, VK, ACA, GCP, DME, MCB

This publication is the work of the authors and TAB, MRJ and DAL will serve as guarantors for the contents of this paper. All authors read and approved the final manuscript.

## Competing interests

DAL has received support from numerous national and international government and charity funders and from Medtronic LTD and Roche Diagnostics for research unconnected with that presented in this study. DAL is an associate editor for BMC Medicine. GCP is an employee of 23andMe Inc and may hold stock or stock options. All other authors report no conflict of interest.

## Acknowledgements

We thank Tom Palmer, Eleanor Sanderson and Stephen Burgess for helpful discussions, Mark Iles for provision of BiB ethnicity variables and Amanda Hill and David Hughes for support in delivery and management of the ALSPAC data. We gratefully acknowledge the GIANT (Genetic Investigation of ANthropometric Traits) Consortium for making GWAS summary statistics available. This publication is the work of the authors and may not reflect the views of those acknowledged here. Where authors are identified as personnel of the International Agency for Research on Cancer / World Health Organization, the authors alone are responsible for the views expressed in this article and they do not necessarily represent the decisions, policy or views of the International Agency for Research on Cancer / World Health Organization.

We are extremely grateful to all the families who took part in ALSPAC, the midwives for their help in recruiting them, and the whole ALSPAC team, which includes interviewers, computer and laboratory technicians, clerical workers, research scientists, volunteers, managers, receptionists and nurses. Born in Bradford is only possible because of the enthusiasm and commitment of the Children and Parents in BiB. We are grateful to all the participants, health professionals and researchers who have made Born in Bradford happen. We gratefully acknowledge the contribution of TPP and the TPP ResearchOne team in completing study participant matching to GP primary care records and in providing ongoing informatics support. This research has been conducted using the UK Biobank Resource under applications 10035, 13436 and 236 granting access to the corresponding UK Biobank genetic and phenotype data. We are extremely grateful to all the UK Biobank participants, investigators and team members.

## Availability of data and materials

The data that support the findings of this study are available from the ALSPAC, BiB and UK Biobank executives, but restrictions apply to the availability of these data, which were used under license for the current study, and so are not publicly available. The ALSPAC study website (http://www.bristol.ac.uk/alspac/researchers/our-data/) contains details of all the data that are available through a fully searchable data dictionary and variable search tool. Scientists are encouraged and able to use BiB data. Data requests are made to the BiB executive using the form available from the study website: http://www.borninbradford.nhs.uk (please click on ‘Science and Research’ to access the form). Guidance for researchers and collaborators, the study protocol and the data collection schedule are all available via the website. All requests are carefully considered and accepted where possible. UK Biobank data are available from the UK Biobank (http://biobank.ndph.ox.ac.uk/showcase/). GWAS summary statistics are publicly available from the GIANT consortium website: https://portals.broadinstitute.org/collaboration/giant/index.php/GIANT_consortium_data_files. (83).

## Funding

The UK Medical Research Council and Wellcome (102215/2/13/2) and the University of Bristol provide core support for ALSPAC. Genotyping of the ALSPAC maternal samples was funded by the Wellcome Trust (WT088806) and the offspring samples were genotyped by Sample Logistics and Genotyping Facilities at the Wellcome Trust Sanger Institute and LabCorp (Laboratory Corporation of America) using support from 23andMe. A comprehensive list of grants funding is available on the ALSPAC website (http://www.bristol.ac.uk/alspac/external/documents/grant-acknowledgements.pdf). BiB receives core infrastructure funding from the Wellcome Trust (WT101597MA), a joint grant from the UK Medical Research Council (MRC) and UK Economic and Social Science Research Council (ESRC) (MR/N024397/1), the British Heart Foundation (CS/16/4/32482) and the National Institute for Health Research (NIHR) under its Applied Research Collaboration Yorkshire and Humber (NIHR200166). Further support for genome-wide data is from the UK Medical Research Council (G0600705) and the National Institute of Health Research (NF-SI-0611-10196). The work presented here was supported by the US National Institute of Health (R01 DK10324), the European Research Council under the European Union’s Seventh Framework Programme (FP7/2007-2013)/ERC grant agreement (669545) and the British Heart Foundation (AA/18/7/34219). TAB is supported by the Medical Research Council (MRC) (UK) (MR/K501281/1), TAB and DME are supported by the NHMRC (Australia) (GNT1183074 and GNT1157714), DAL, TAB, MCB and RCR work in/are affiliated with a unit that is supported by the UK Medical Research Council (MC_UU_00011/1 & MC_UU_00011/6) and DAL is a British Heart Foundation Chair (CH/F/20/90003) and NIHR Senior Investigator (NF-0616-10102). DAL, MCB and TAB are supported by the British Heart Foundation Accelerator Award at the University of Bristol (AA/18/7/34219). MCB’s contribution to this work was supported by a UK MRC Skills Development Fellowship (MR/P014054/1). VK is funded by the European Union’s Horizon 2020 research and innovation program under the Marie Sklodowska-Curie grant (721567). MRJ is funded by EU-H2020 LifeCycle Action (733206) which also supports DAL’s research, EU-H2020 EDCMET (825762), EU-H2020 EUCAN Connect (824989), EU H2020-MSCA-ITN-2016 CAPICE Marie Sklodowska-Curie grant (721567) and the MRC (UK) (MRC/BBSRC MR/S03658X/1 [JPI HDHL]). RCR is a de Pass Vice Chancellor’s Research Fellow at the University of Bristol. AL and MRJ are supported by the MRC (UK) (MR/M013138/1) and the European Union Horizon 2020 programme (633595). The funders had no role in study design, data collection and analysis, decision to publish, or preparation of the manuscript.

## Additional material

File name: “Additional_file_1.pdf”

Title: Supplementary information

Description: Additional file 1: Supplementary information S1–S35. For descriptions of individual Supplementary information items, please see the contents page of Additional file 1.

## Notes

### Author Declarations

Ethical approval was obtained from the ALSPAC Ethics and Law Committee and the Local Research Ethics Committees, and Bradford National Health Service Ethics Committee

