## Supplementary information for "Exploring the causal effect of maternal pregnancy adiposity on offspring adiposity: Mendelian randomization using polygenic risk scores"

Tom A Bond[1,2,3,4,5,*], Rebecca C Richmond[4,5], Ville Karhunen[1,6,7], Gabriel Cuellar-Partida[3,8], Maria Carolina Borges[4,5], Verena Zuber[1,9], Alexessander Couto Alves[1,10], Dan Mason[11], Tiffany C Yang[11], Marc J Gunter[12], Abbas Dehghan[1,2], Ioanna Tzoulaki[1,2,13], Sylvain Sebert[6], David M Evans[3,4,14], Alex M Lewin[1,15], Paul F O'Reilly[16], Deborah A Lawlor[4,5,§], Marjo-Riitta Järvelin[1,2,6,17,18,§]

[1]Department of Epidemiology and Biostatistics, Imperial College London, London, UK.
[2]MRC-PHE Centre for Environment and Health, School of Public Health, Imperial College London, London, UK.
[3]The University of Queensland Diamantina Institute, The University of Queensland, Brisbane, Australia.
[4]MRC Integrative Epidemiology Unit at the University of Bristol, Bristol, UK.
[5]Population Health Sciences, Bristol Medical School, University of Bristol, Bristol, UK.
[6]Center for Life-course Health Research, Faculty of Medicine, University of Oulu, Oulu, Finland.
[7]Research Unit of Mathematical Sciences, University of Oulu, Oulu, Finland.
[8]23andMe, Inc., Sunnyvale, CA, USA.
[9]MRC Biostatistics Unit, School of Clinical Medicine, University of Cambridge, Cambridge, UK.
[10]School of Biosciences and Medicine, Faculty of Health and Medical Sciences, University of Surrey, Guildford, UK.
[11]Born in Bradford, Bradford Institute for Health Research, Bradford Teaching Hospitals NHS Foundation Trust, Bradford, UK.
[12]Section of Nutrition and Metabolism, IARC, Lyon, France.
[13]Department of Hygiene and Epidemiology, University of Ioannina Medical School, Ioannina, Greece
[14]Institute for Molecular Bioscience, University of Queensland, Brisbane, Australia.
[15]Department of Medical Statistics, London School of Hygiene and Tropical Medicine, London, UK.
[16]Genetics and Genomic Sciences, Icahn School of Medicine at Mount Sinai, New York, USA.
[17]Unit of Primary Care, Oulu University Hospital, Oulu, Finland.
[18]Department of Life Sciences, College of Health and Life Sciences, Brunel University London, London, UK.

§These authors contributed equally to this work

**Supplementary information S1: Description of the Avon Longitudinal Study of Parents and Children (ALSPAC) and Born in Bradford**

We analysed data from two population based prospective birth cohorts: the Avon Longitudinal Study of Parents and Children (ALSPAC) and Born in Bradford (BiB). ALSPAC enrolled pregnant women who resided in and around the city of Bristol in the South West of England and had an expected delivery date between April 1, 1991 and December 31, 1992. The enrolled cohort included 15,247 pregnancies resulting in 14,775 live born babies. Ethical approval was obtained from the ALSPAC Ethics and Law Committee and the Local Research Ethics Committees. The study website contains details of all the data that are available through a fully searchable data dictionary and variable search tool and details of the study methodology have been reported previously (1, 2). BiB enrolled women who resided in the city of Bradford in the North of England who attended an antenatal booking clinic between March 2007 and December 2010. The recruited cohort included 13,776 pregnancies resulting in 13,740 live born babies. Ethical approval was obtained from Bradford National Health Service Ethics Committee (ref 06/Q1202/48) and details of the study methodology have been reported previously (3).

**Supplementary information S2: Selection of study participants**

For the present analysis we included live-born singletons with maternal and offspring genotype data, maternal BMI data and at least one offspring adiposity measure available, and selected one offspring from any sibling groups for inclusion (chosen at random in ALSPAC or to maximise the sample size with data available in BiB). As the effects we were exploring may differ by ethnicity (4) we limited analyses to two ethnic groups: White European and South Asian. There were very few participants from other ethnic groups in either cohort, therefore these participants were excluded. ALSPAC (93% White European) contributed only to the analyses in White Europeans and we meta-analysed these results with those from models fitted separately for BiB South Asians and BiB White Europeans. Derivation of ethnicity variables is described in **Supplementary information S4**.

**Supplementary information S3: Flow charts describing sample selection**

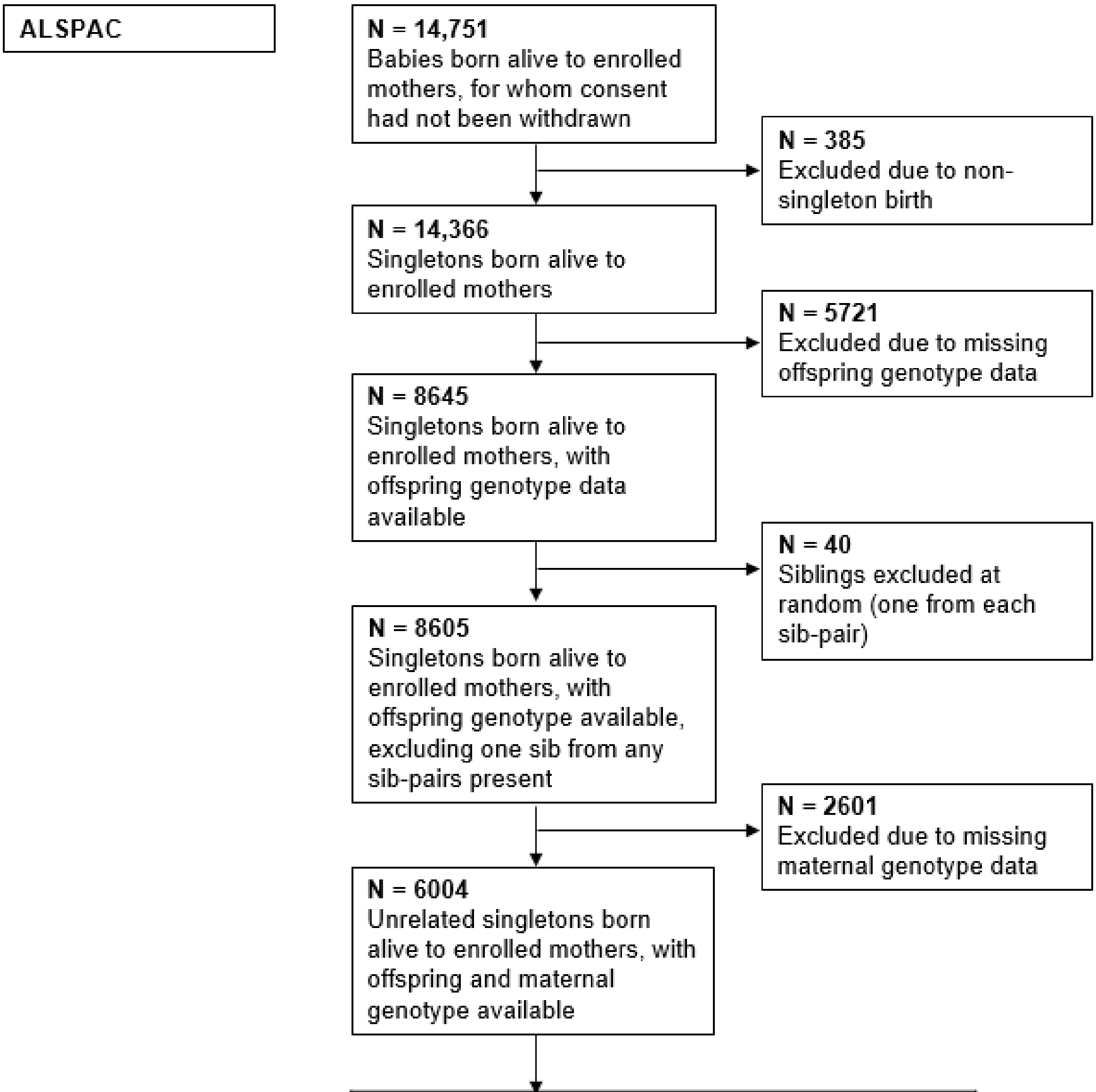

Individuals excluded due to missing phenotypic data

| Offspring phenotype | N with phenotypic data available[a] |
|---|---|
| Birth weight | 5085 |
| 1yr BMI | 4838 |
| 4yr BMI | 4670 |
| 10yr BMI | 4476 |
| 15yr BMI | 4112 |
| 10yr FMI | 3855 |
| 12yr FMI | 3807 |
| 14yr FMI | 3506 |
| 16yr FMI | 2996 |
| 18yr FMI | 2659 |

a. Phenotypic data defined as maternal BMI, maternal age, offspring phenotype and offspring age at phenotype measurement

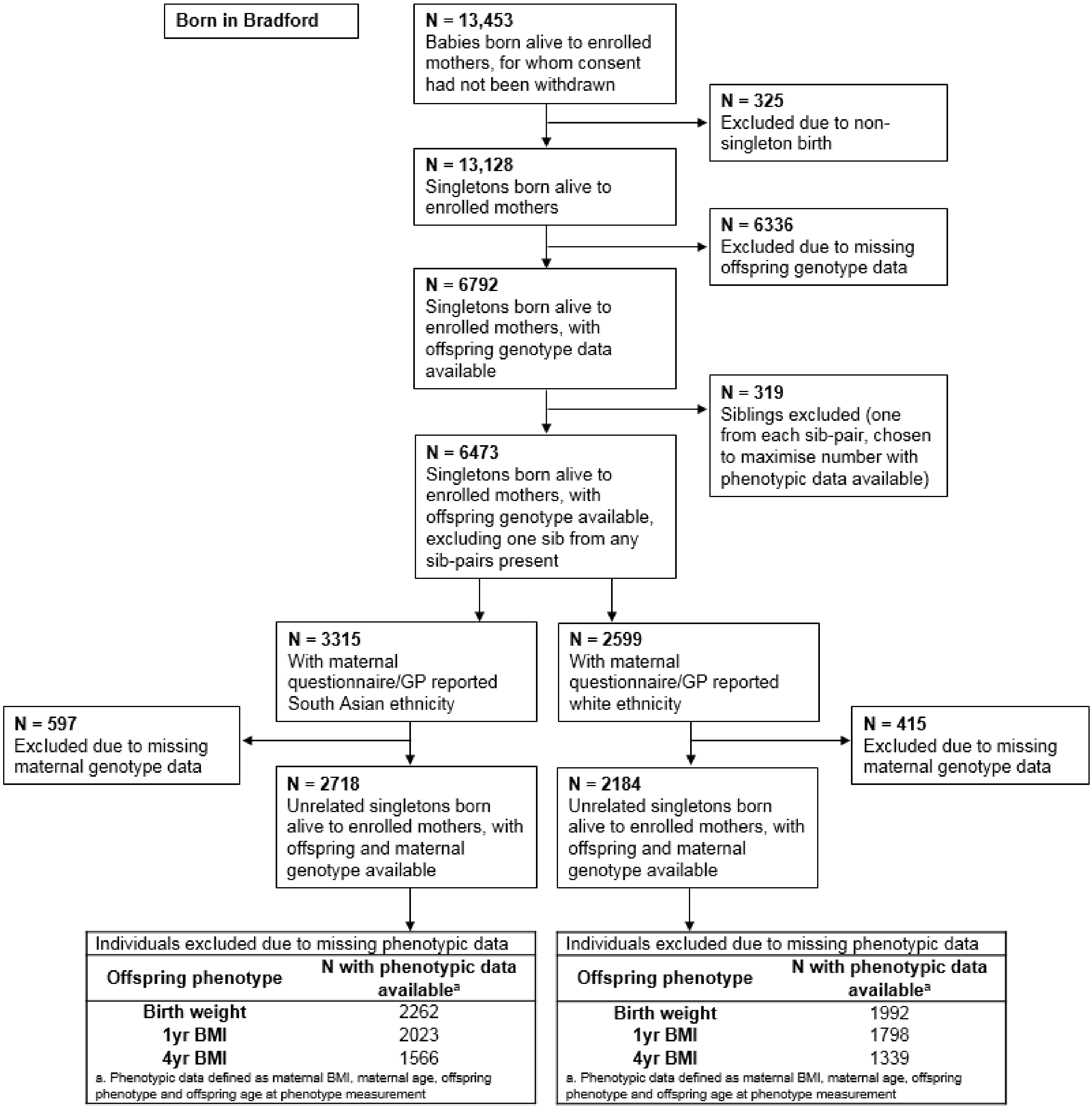
Born in Bradford
N = 13,453
Babies born alive to enrolled mothers, for whom consent had not been withdrawn
N = 325
Excluded due to non-singleton birth
N = 13,128
Singletons born alive to enrolled mothers
N = 6336
Excluded due to missing offspring genotype data
N = 6792
Singletons born alive to enrolled mothers, with offspring genotype data available
N = 319
Siblings excluded (one from each sib-pair, chosen to maximise number with phenotypic data available)
N = 6473
Singletons born alive to enrolled mothers, with offspring genotype available, excluding one sib from any sib-pairs present
N = 3315
With maternal questionnaire/GP reported South Asian ethnicity
N = 2599
With maternal questionnaire/GP reported white ethnicity
N = 597
Excluded due to missing maternal genotype data
N = 415
Excluded due to missing maternal genotype data
N = 2718
Unrelated singletons born alive to enrolled mothers, with offspring and maternal genotype available
N = 2184
Unrelated singletons born alive to enrolled mothers, with offspring and maternal genotype available
Individuals excluded due to missing phenotypic data
Offspring phenotype
N with phenotypic data available[a]
Birth weight 2262
1yr BMI 2023
4yr BMI 1566
a. Phenotypic data defined as maternal BMI, maternal age, offspring phenotype and offspring age at phenotype measurement
Individuals excluded due to missing phenotypic data
Offspring phenotype
N with phenotypic data available[a]
Birth weight 1992
1yr BMI 1798
4yr BMI 1339
a. Phenotypic data defined as maternal BMI, maternal age, offspring phenotype and offspring age at phenotype measurement

**Supplementary information S4: Derivation of ethnicity variables**

In ALSPAC, ethnicity was assessed by genetic multidimensional scaling (MDS) analysis during the quality control procedure for maternal, paternal and offspring genotype data. BiB mothers reported their ethnicity and the place of birth of their parents at the baseline interview, and where these data were missing we used information extracted from General Practice records. Categories were based on UK Office for National Statistics guidance (5). We constructed a variable with three categories: “South Asian” (composed of “Pakistani”, “Indian” and “Bangladeshi”), “White European” (composed of “White British” and “White Other”) and “Other” (composed of all other ethnicities).

**Supplementary information S5: Data sources for offspring outcomes**

Offspring birth weight (BW) was extracted from the birth record in BiB, and extracted from the birth record/notification or measured by research staff in ALSPAC. Birth length in ALSPAC was obtained similarly, and is not available for BiB participants. In ALSPAC, child and adolescent height/length and weight were obtained from clinical examination by study staff using a Harpenden Stadiometer (Holtain Limited, Dyfed, UK) and Tanita Body Fat Analyser (Model TBF 305; Tanita UK Limited, Viewsley, UK) respectively, or from child health records or maternal/offspring questionnaire responses (Supplementary information S5). Although the Tanita Body Fat Analyser is capable of measuring body fat mass by bioelectrical impedance, we had access to fat mass measured by whole body dual-energy X-ray absorptiometry (DXA; see below), therefore we only used weight measurements from the Tanita Body Fat Analyser. In BiB, childhood height and weight were obtained from a variety of sources including clinical measurement by study staff or measurement at around age 4 years as part of the UK Government National Child Measurement Programme (NCMP) (in both cases using a Leicester Height Measure [Seca] and Seca digital scales), child health records, primary care records and school nurse records (Supplementary information S5). In ALSPAC, we calculated FMI as fat mass (kg) / height $(m)^2$ using fat mass measured by whole body DXA carried out with a Lunar Prodigy DXA scanner (GE Medical Systems Lunar, Madison, WI, USA). The scans were visually inspected and realigned where necessary. Once complete, the tester examined the scan to ensure its quality, and if necessary repeated the scan.

Offspring body mass index (BMI) measurements were available from a variety of sources, as described in **Supplementary information S7**. In order to maximise the sample size available for analysis and to facilitate comparisons with previous work in which we took a similar approach (6), we used offspring BMI measurements from all available sources. We selected measurements within four age windows throughout childhood and adolescence, giving mean ages close to 1, 4, 10 and 15 years (target ages). Within windows data points were first selected to prioritise higher quality data sources (clinical exam by study staff > general practice records or UK Government National Child Measurement Programme [NCMP] records > growth records > questionnaire) and secondly to minimise the age difference from the target ages.

| Target age | Lower and upper boundaries of age window (years) | Cohorts with data available |
|---|---|---|
| 1 year | ≥0.5, <2 | ALSPAC, BiB |
| 4 years | ≥3, <7 | ALSPAC, BiB |
| 10 years | ≥8, <12 | ALSPAC |
| 15 years | ≥13, <17 | ALSPAC |

We derived fat mass index (FMI) outcomes from dual-energy X-ray absorptiometry (DXA) measurements taken at a series of clinical examinations at which the participants had approximate mean ages of 10, 12, 14, 16 and 18 years, enabling comparison with the results of a previous study that used ALSPAC data (7).

**Supplementary information S6: Source of anthropometric measurements**

| Variable | Sample | Data source |
|---|---|---|
| Maternal (pre-)pregnancy BMI | ALSPAC | Height and pre-pregnancy weight reported by the mothers during pregnancy (~90% of entire baseline sample) or 4 months postnatally (~10% of entire baseline sample) |
| | BiB (SA and WE) | Height reported by the mothers at recruitment (26–28 weeks gestation), weight measured at the first antenatal clinic assessment (median 12 weeks gestation) and abstracted from the medical records |
| Paternal BMI | ALSPAC | Height and weight reported by the fathers during their partner's pregnancy (or postnatally for a minority of fathers) |
| | BiB (SA and WE) | Height and weight reported by the fathers at the time of their partner's recruitment |
| Birth weight and length | ALSPAC | Weight measured by trained research assistants, abstracted from the birth record or abstracted from the birth notification. Length measured by trained research assistants or abstracted from the birth record |
| | BiB (SA and WE) | Weight abstracted from the birth record |
| 1 year BMI | ALSPAC | Weight and recumbent length measured clinically (14.1%), abstracted from growth records (84.9%) or from postal questionnaire (1.0%) |
| | BiB (SA) | Weight and recumbent length measured clinically (22.0%), abstracted from primary care records (33.6%) or abstracted from child health records (44.3%) |
| | BiB (WE) | Weight and recumbent length measured clinically (18.3%), abstracted from primary care records (35.7%) or abstracted from child health records (46.1%) |
| 4 year BMI | ALSPAC | Weight and height measured clinically (13.3%), abstracted from growth records (74.6%) or from postal questionnaire (12.1%) |
| | BiB (SA) | Weight and height measured clinically (16.9%), abstracted from primary care records (12.1%), abstracted from child health records (1.8%) or NCMP (69.3%) |
| | BiB (WE) | Weight and height measured clinically (15.1%), abstracted from primary care records (9.9%), abstracted from child health records (2.2%) or NCMP (72.8%) |
| 10 year BMI | ALSPAC | Weight and height measured clinically (99.3%) or from postal questionnaire (0.7%) |
| 15 year BMI | ALSPAC | Weight and height measured clinically (94.1%) or from postal questionnaire (5.9%) |

**SA**: South Asians, **WE**: White Europeans, **NCMP**: National Child Measurement Programme, **BMI**: body mass index

**Supplementary information S7: Assessment of other variables**

In ALSPAC, offspring sex data were abstracted from the birth record, as were gestational age at delivery data (which will have largely been based on the date of the mothers' last menstrual period as per contemporary UK clinical practice, with some potential modification based on first trimester ultrasound scan or clinical assessment at birth). Maternal age at delivery was calculated from maternal date of birth and offspring date of delivery. Paternal age when the mother was recruited was obtained from a questionnaire completed by the father. Parity (defined as the number of previous pregnancies resulting in a live or stillbirth), parental occupation, maternal and paternal education, maternal smoking during pregnancy and maternal education variables were derived from questionnaires completed by the mothers during pregnancy. Parental occupation was derived from the highest occupational group of the mother or father and coded in five categories: class I (professional occupations), class II (managerial and technical occupations), class III (skilled manual occupations), class IV (partly skilled occupations) and class V (unskilled occupations). Maternal smoking was coded in three categories: "never smoked during pregnancy", "smoked in early pregnancy only" and "smoked throughout pregnancy". Highest maternal and paternal educational qualifications were treated as separate variables and were coded in five categories: "no qualifications or Certificate of Secondary Education", "vocational qualifications", "General Certificate of Education (GCE) (ordinary level)", "GCE (advanced level)", and "university degree".

In BiB, data on offspring sex, maternal parity and gestational age at delivery (in completed weeks) were abstracted from the birth record. All participants who were first seen early enough in their pregnancy were invited to have a first trimester 'dating' ultrasound scan and results from this, maternal reported last menstrual period and appearance of the infant at birth were used to estimate gestational age. Maternal age at delivery, smoking, maternal and paternal education and paternal occupation data were obtained from a questionnaire completed by the mothers at recruitment (26–28 weeks gestation). Paternal occupation was coded in 12 categories: "modern professional occupations", "clerical and intermediate occupations", "senior managers or administrators", "technical and craft occupations", "semi-routine manual and service occupations", "routine manual and service occupations", "middle or junior managers", "traditional professional occupations", "self-employed", "student/in training", "long term unemployed / sick" and "does not know". Highest maternal and paternal educational qualifications were treated as separate variables and were coded in seven categories: "<5 General Certificate of Secondary Education (GCSE) equivalent", "5 GCSE equivalent", "A level equivalent", "Higher than A level", "other" (e.g. City and Guilds, RSA/OCR, BTEC), "does not know" and "foreign unknown qualification". The parents' highest educational qualifications were equivalized (based on the qualification received and the country in which it was obtained) using the UK National Agency for the Recognition and Comparison of International Qualifications and Skills (NARIC; https://www.naric.org.uk/naric/) system. "Does not know" relates to the mother responding "don't know" during interview. Foreign unknown relates to a qualification reported that does not appear in the NARIC list of qualifications. Maternal smoking was coded in two categories: "smoked during pregnancy", "did not smoke during pregnancy". Paternal age was reported by the fathers at the time of their partner's recruitment.

**Supplementary information S8: Genetic principal component calculation**

For the primary analyses we adjusted for principal components (PCs) calculated from the called (as opposed to imputed) offspring genotype data in order to control for population stratification. We first removed regions of long-range LD taken from Price *et al*. (8) (the QC steps described in **Supplementary information S8** having been applied previously). We then carried out pruning using the PLINK 1.9 (9) command *--indep-pairwise 1000 80 0.1*, and calculated PCs using the PLINK 1.9 *--pca* command. We calculated PCs separately for all samples (ALSPAC, BiB South Asians, BiB White Europeans and BiB South Asians and White Europeans combined). For the sensitivity analyses in which we removed cryptic relatedness (**Main text**) we did this prior to calculating PCs.

**Supplementary information S9: Genotyping, quality control and imputation**

ALSPAC mothers were genotyped at Centre National de Génotypage, Paris, France, using the Illumina Human 660W-quad array and genotypes were called with Illumina GenomeStudio. SNPs with call rate <95%, lack of Hardy-Weinberg equilibrium (HWE; P <1.0e-6) or minor allele frequency (MAF) <1% were excluded. Individuals with missingness >5%, indeterminate X chromosome heterozygosity, extreme autosomal heterozygosity or potential ID mismatches were excluded. Population stratification was assessed by multidimensional scaling (MDS) and compared with Hapmap phase 2 reference populations (10); all individuals with non-European ancestry were removed.

ALSPAC offspring were genotyped at the Wellcome Trust Sanger Institute, Cambridge, UK and the Laboratory Corporation of America, Burlington, NC, USA using the Illumina HumanHap550 quad chip array. SNPs with call rate <95%, lack of HWE (P <5e-7) or MAF <1% were excluded. Individuals with gender mismatches, minimal or excessive heterozygosity, missingness >3%, insufficient sample replication (IBD <0.8) or potential ID mismatches were excluded. Population stratification was assessed by MDS and compared with Hapmap phase 2 reference populations; all individuals with non-European ancestry were removed.

ALSPAC fathers were genotyped at the ALSPAC Laboratory, Bristol, UK, using the Illumina HumanCoreExome array. SNPs with call rate <95%, lack of HWE (P <1e-7), duplicate SNPs or those failing GenomeStudio quality control (QC) measures were excluded. Individuals with gender mismatches, minimal or excessive heterozygosity, missingness >5%, possible sample contamination or discordant lab assigned and genetically assigned IDs were excluded. Population stratification was assessed by MDS and compared with Hapmap phase 2 reference populations; all individuals with non-European ancestry were removed. Cryptic relatedness was removed using a relatedness filter of 0.1 in the GCTA software package (11).

BiB mothers and offspring of all ethnicities were genotyped at Bristol Bioresource Laboratories, Bristol, UK using Illumina HumanCoreExome12v1.0, HumanCoreExome12v1.1 and HumanCoreExome24v1.0 arrays and genotypes were called with Illumina GenomeStudio. Individuals with high genotype missingness and SNPs with low call rate were removed using an iterative procedure, resulting in a final sample of individuals with missingness <0.5% and SNPs with call rate >99.5%. Other typically used QC metrics such as deviation from HWE and excess heterozygosity are not appropriate here given the population structure and consanguineous union rates known to be present. For all cohorts, the PLINK software package (v1.07) was used to carry out QC measures on called genotypes.

For ALSPAC, array genotypes were harmonized, phased using SHAPEIT v2 (12) and subsequently imputed via the Michigan imputation server (13) to the Haplotype Reference Consortium (HRC) reference panel (14) (for mothers and children) or to the 1000 Genomes phase 1 version 3 reference panel (15) (for fathers). For BiB, array genotypes were harmonized and subsequently phased and imputed via the Sanger Imputation Service (14) using the "UK10K + 1000 Genomes Phase 3 reference panel" (15, 16) and the "pre-phase with EAGLE2 and impute" pipeline (17). After imputation, MAF, HWE and imputation quality score filters were applied as described in the table immediately below.

| **Sample** | **MAF threshold** [a] | **Imputation quality score threshold** | **HWE *P*-value threshold** |
|---|---|---|---|
| ALSPAC mothers and offspring | Minor allele count >5 [a] | $r^2$ >0.3 | HWE *P* >1e-6 |
| ALSPAC fathers | MAF >1% | $r^2$ >0.8 | None |
| BiB | Minor allele count >5 [a] | INFO score >0.3 | None [b] |

**a**: When calculating the genetic relatedness matrices used for the linear mixed models we applied a MAF filter of 1%
**b**: deviation from HWE and is not an appropriate quality control metric for BiB, given the population structure and consanguineous union rates known to be present. **MAF**: minor allele frequency

**Supplementary information S10: (a) Directed acyclic graph (DAG) showing the assumptions of our MR analyses, and (b) potential violations of these assumptions**

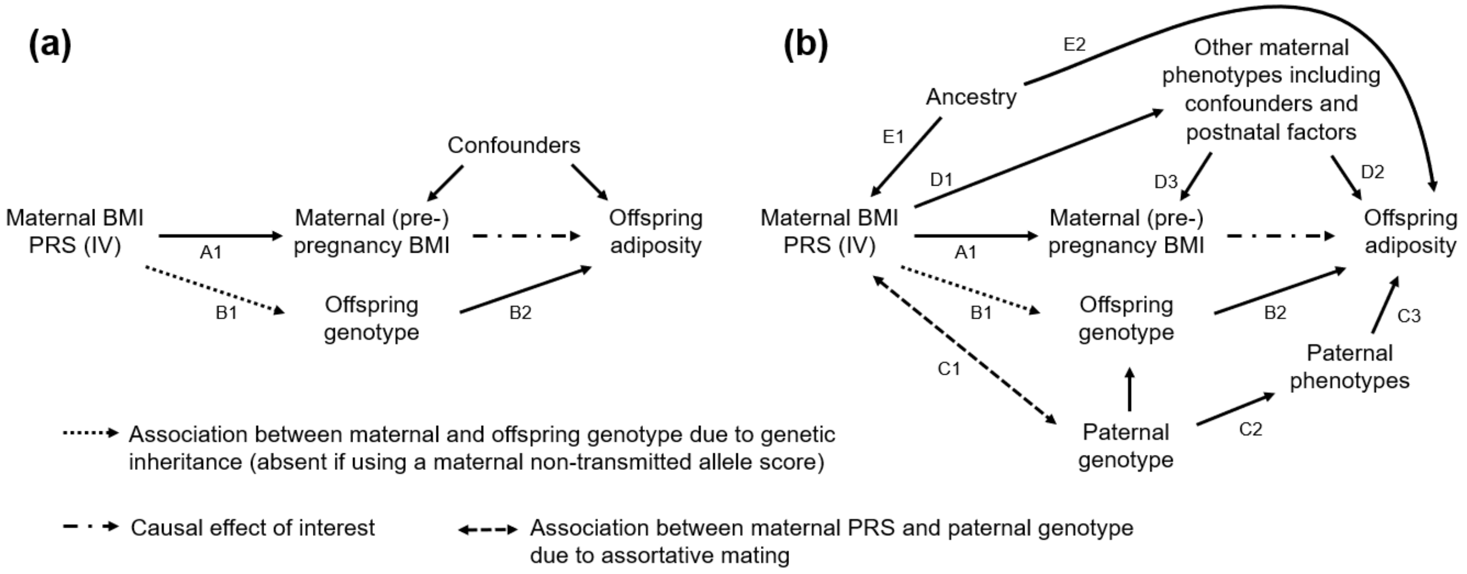

Single headed arrows show that we know or think it is plausible that the variable at the tail of the arrow causes the variable at the head. The absence of an arrow between two variables shows we do not believe there is a direct causal effect between them (18). MR makes three core assumptions (19), including:

- IV assumption 1: the instrumental variable (IV) is associated with the exposure (path A1 exists)
- IV assumption 2: the IV outcome association is not confounded (e.g. path E1-E2 does not exist)
- IV assumption 3: the IV is not associated with the outcome except via its association with the exposure (there are no other arrows from maternal BMI PRS to offspring adiposity, e.g. B1-B2, C1-C2-C3, D1-D2).

In the present study, it was necessary to ensure that IV assumption 3 was not violated by maternal alleles that are inherited by the offspring, and subsequently cause offspring adiposity (path B1-B2). We used only maternal alleles that were not inherited by the offspring to calculate the maternal BMI PRS (20). Under random mating, this maternal non-transmitted allele score should be independent of offspring genotype, therefore path B1 should be absent. A previous MR investigation of the maternal-offspring adiposity association used an alternative approach involving conditioning on the offspring's BMI PRS (a weighted allele score) (7). Such an approach may not be optimal due to (i) imperfect control for genetic inheritance (21), and (ii) collider bias (conditioning on offspring genotype will induce a spurious correlation between maternal and paternal genotype [path C1], resulting in a biased MR estimate if paths C2 and C3 exist) (22, 23). Path C1 may also exist due to assortative mating. Other potential violations of the core IV assumptions, along with the steps we have taken to explore and account for these, are described in the **Main Text (Methods)**.

In order to obtain a point estimate for the causal effect it is necessary to make further assumptions (24). In particular, it is assumed that both the IV-exposure association and the exposure-outcome causal effect are linear (25), and that either the causal effect is homogeneous (there is no effect modification by exposure-outcome confounders (24, 26)), or that for each individual the exposure is a monotonic (increasing or decreasing) function of the IV (24, 25). We conduct our analyses under the monotonicity assumption, therefore the MR effect corresponds to the local average treatment effect (LATE; i.e. the average causal effect in individuals for whom increased maternal non-transmitted allele score would cause increased maternal BMI, known as the "complier" group) (24). It is plausible that all individuals in the population are compliers, in which case the LATE is equivalent to the population average treatment effect (ATE).

**Supplementary information S11: Polygenic risk scoring methods and UK Biobank BMI GWAS**

We tested four methods for calculating the BMI PRS. All four methods

(i) involve calculating the PRS in the target sample (ALSPAC or BiB) as a weighted sum of BMI-increasing alleles at genetic variants (hereafter referred to as SNPs) across the genome

(ii) aim to maximise phenotypic prediction (measured as $R^2$, the proportion of variance of the exposure [maternal BMI] explained by the PRS) by optimising the number of SNPs included in the score and the weights given to SNPs, accounting for correlations between SNPs (linkage disequilibrium [LD])

(iii) require an independent base sample for the calculation of SNP weights (for some methods the weights are modified subsequently)

The methods that we tested were:

1. Clumping and thresholding (C+T) (27): a clumping algorithm (28) is used to select a set of SNPs that are not in strong LD in the target sample, taking account of the *P*-values for each SNP from a genome-wide association study (GWAS) of the phenotype conducted in an independent (base) sample (i.e. to avoid discarding SNPs that are strongly associated with the phenotype). SNPs are then included in the PRS based on a *P*-value threshold from the base GWAS; typically several thresholds are explored, yielding PRS calculated from different numbers of SNPs. PRS are then calculated as a weighted sum of BMI-increasing alleles, with SNPs weighted by their effects (beta coefficients) from the base GWAS
2. LDpred (29): a Bayesian model is used to estimate posterior SNP weights from the base GWAS SNP effects, assuming a point-normal mixture prior and accounting for LD information from a reference panel. A model parameter specifying the fraction of causal SNPs (*p*) must be chosen; typically several values are explored with the aim of maximising phenotypic prediction. The PRS is then calculated similarly to above as a weighted sum of BMI-increasing alleles
3. lassosum (30): penalised regression is used to carry out shrinkage and selection on the base GWAS SNP effects, accounting for LD information from an external reference panel. Two regularisation parameters ($\lambda$ and *s*) must be chosen; typically several values of each are explored with the aim of maximising phenotypic prediction. The PRS is then calculated similarly to above as a weighted sum of BMI-increasing alleles
4. the BOLT-LMM linear predictor (31): SNP weights are calculated by fitting all SNPs as random effects in order to account for LD, using individual participant data from the base sample. BOLT-LMM is a Bayesian model and assumes a mixture of Gaussians prior on SNP effects (if a single Gaussian prior was specified instead the model would be equivalent to best linear unbiased prediction [BLUP] (31)). The PRS is then calculated similarly to above as a weighted sum of BMI-increasing alleles.

Methods 1–3 require GWAS summary statistics from the largest possible sample (the base dataset) that is independent from the target datasets (ALSPAC and BiB) (32, 33). It is important to avoid overlap of individuals between the base and target datasets because this can cause overfitting and lead to severely inflated prediction $R^2$ for the target phenotype (34). We therefore conducted a GWAS in the UK Biobank (UKB), which is a prospective cohort of 502,628 volunteers recruited across the UK at age 40–69 years through United Kingdom National Health Service registers. Details of UKB study design and genotype data are described in full elsewhere (35, 36). Participants attended dedicated assessment centres across the UK between 2006 and 2010, during which weight and height were measured by trained study personnel. We carried out a GWAS on UKB individuals with imputed genotype and BMI data (field ID f.21001.0.0) available and self-reported white ethnicity (field ID f.21000.0.0). Because ALSPAC and BiB parents could also be participants in UKB, we used two samples, excluding participants attending the Bristol assessment centre (remaining $N$ = 416,824) or the Leeds assessment centre (remaining $N$ = 416,352) in order to minimise overlap with ALSPAC and BiB respectively (field ID f.54.0.0). We treated the BMI phenotype similarly to previous GWAS studies (37, 38): we regressed BMI on age (field ID f.21003.0.0), age squared, batch (field ID f.22000.0.0) and assessment centre (field ID f.54.0.0) separately for each sex (field ID f.31.0.0). We then used inverse-normal transformed residuals from these regression models as the phenotype for the GWAS analyses. We ran GWAS using a linear mixed model implemented in the BOLT-LMM software package to control for population structure (31) and tested for association of the phenotype with ~45 million autosomal SNPs with MAF >0.01% and imputation INFO score >0.3, using imputed genotype probabilities and assuming an infinitesimal model. BOLT-LMM requires a set of hard-called (i.e. integer valued) genotypes with which to build the LMM; following a similar analysis by the BOLT-LMM authors (31) we used 672,345 genotyped SNPs with missingness <10% and MAF >0.1%. The BOLT-LMM authors also recommend including genetic PCs as fixed effects in the LMM in order to speed up model convergence (31), therefore we included 20 PCs which we calculated from the same set of SNPs using the FastPCA algorithm (39) (as implemented in PLINK 2.0 (9) --*pca approx*) as fixed effects. We then meta-analysed the summary statistics from our two UKB GWAS with the largest available published BMI GWAS from the GIANT consortium (37) using the METAL software package version 2011-03-25 (40), having first removed ambiguous SNPs (A/T and G/C SNPs). These meta-analyses yielded two sets of GWAS summary statistics ([i] GIANT + UKB excluding Bristol, and [ii] GIANT + UKB excluding Leeds, to be used for analyses of ALSPAC and BiB respectively) giving a total base sample size of up to 756,048 individuals and summary statistics for >2 million SNPs.

Clumping and thresholding

We first applied the C+T method, using the PRSice2 software package version 2.1.3beta (27). For the target datasets (ALSPAC and BiB) we used the subset of genotyped autosomal SNPs (having first applied quality control [QC] steps as described in **Supplementary information S9**) that were in common with the base dataset. For each target dataset we used the appropriate meta-analysed base GWAS dataset as described above. We applied the default PRSice2 clumping parameters: --*clump-kb 250*, --*clump-r2 0.1* and --*clump-p 1*, resulting in a set of SNPs that were near-independent in the target dataset. We then tested *P*-value thresholds between zero and 1 (at increments of 0.01) to find the threshold resulting in the highest PRS $R^2$ for maternal BMI, in linear regression models with 20 PCs calculated from maternal genotype fitted as covariates. The best *P*-value thresholds and PRS $R^2$ for maternal BMI for all target samples are shown in the table immediately below ($R^2$ is for PRS calculated from maternal genotype, as opposed to the maternal non-transmitted allele PRS used for the MR analyses).

| **Target sample** | **Best *P*-value threshold** | ***N* SNPs included** | **PRS $R^2$ (maternal BMI)** |
|---|---|---|---|
| ALSPAC | 0.03 | 16,369 | 9.9% |
| BiB (all ethnicities) | 0.06 | 15,179 | 6.8% |
| BiB (Pakistanis) | 0.08 | 17,240 | 6.0% |
| BiB (White British) | 0.30 | 27,654 | 7.8% |

LDpred

We next applied the LDpred method, using the LDpred software package version 0.9.9 (29). We used target and base datasets as described for the C+T method, and ran LDpred with the LD radius parameter set to *M*/3000 (where *M* is the number of SNPs) as recommended by the authors. We used the target datasets as the LD reference, and explored a range of values for the parameter *p* (the fraction of SNPs that have a non-zero effect on the phenotype) (1, 0.3, 0.1, 0.03, 0.01, 0.003, 0.001), as well as the infinitesimal model (which assumes a Gaussian prior on SNP effects). We subsequently used the *--score* function in the PLINK software package version 1.90 to calculate PRS in the target samples. The best values of *p* and the corresponding PRS $R^2$ for maternal BMI for all target samples, in linear regression models with 20 PCs calculated from maternal genotype fitted as covariates are shown in the table immediately below ($R^2$ is for PRS calculated from maternal genotype, as opposed to the maternal non-transmitted allele PRS used for the MR analyses).

| **Target sample** | **Best *p* parameter value** | ***N* SNPs included*** | **PRS $R^2$ (maternal BMI)** |
|---|---|---|---|
| ALSPAC | 1.0 | 375,261 | 12.1% |
| BiB (all ethnicities) | Infinitesimal model | 221,265 | 8.1% |
| BiB (Pakistanis) | Infinitesimal model | 220,315 | 7.6% |
| BiB (White British) | Infinitesimal model | 220,040 | 8.6% |

*For the LDPred PRS all the SNPs were included, but many of the included SNPs had weights close to zero

Lassosum

We applied the lassosum method using the lassosum R package version 0.4.3 (30). We used target and base datasets as described for the C+T method (with the exception that for ALSPAC we ran lassosum on the set of SNPs that were common to the base dataset, the set of genotyped SNPs in the mothers and offspring and the set of imputed SNPs in the fathers [see **Supplementary information S9**]). We ran lassosum using the European 1000 Genomes populations (EUR) LD region file as defined in Berisa *et al* (41), and used the target dataset as the LD reference panel. Results were similar for BiB when we instead used the Asian (ASN) LD region file. We explored the default grid of parameter values for *λ* (20 values ranging from 0.001 to 0.1) and *s* (0.2, 0.5, 0.9, 1.0). The best values of *λ* and *s* and the corresponding PRS $R^2$ for maternal BMI for all target samples, in linear regression models with 20 PCs calculated from maternal genotype fitted as covariates are shown in the table immediately below ($R^2$ is for PRS calculated from maternal genotype, as opposed to the maternal non-transmitted allele PRS used for the MR analyses).

| **Target sample** | **Best *λ* and *s* parameter values** | | ***N* SNPs included** | **PRS $R^2$ (maternal BMI)** |
|---|---|---|---|---|
| | ***λ*** | ***s*** | | |
| ALSPAC | 0.00127 | 0.2 | 81,113 | 13.1% |
| BiB (all ethnicities) | 0.00127 | 0.5 | 79,824 | 8.9% |
| BiB (Pakistanis) | 0.00162 | 0.5 | 64,828 | 8.7% |
| BiB (White British) | 0.00100 | 0.2 | 75,639 | 8.6% |

BOLT-LMM linear predictor

We applied the BOLT-LMM linear predictor method using the BOLT-LMM software package version 2.3 (31). We used the *--predBetasFile* command to calculate SNP effects (beta coefficients) for BMI for the set of genotyped SNPs that we used to build the LMM (as described above), in the same UKB samples as for our UKB GWAS (i.e. excluding either Bristol or Leeds assessment centre participants), and fitted 20 PCs calculated from the same set of SNPs as fixed effects. We treated the BMI phenotype as detailed above for our UKB GWAS. We then used PRSice2 to calculate PRS for individuals in the target datasets as sums of BMI increasing alleles weighted by their BOLT-LMM betas, for the SNPs with BOLT-LMM betas available and imputed genotype data available in the target datasets. PRSice2 automatically detects and accounts for strand flips, and we did not use clumping. PRS $R^2$ for maternal BMI for all target samples, in linear regression models with 20 PCs calculated from maternal genotype fitted as covariates are shown in the table immediately below ($R^2$ is for PRS calculated from maternal genotype, as opposed to the maternal non-transmitted allele PRS used for the MR analyses).

| **Target sample** | ***N* SNPs included*** | **PRS $R^2$ (maternal BMI)** |
|---|---|---|
| ALSPAC | 272,172 | 11.9% |
| BiB (all ethnicities) | 441,512 | 7.1% |
| BiB (Pakistanis) | 441,512 | 4.0% |
| BiB (White British) | 441,512 | 8.1% |

*For the BOLT-LMM linear predictor PRS all the SNPs were included, but many of the included SNPs had weights close to zero

Of the four methods, lassosum achieved the highest PRS $R^2$ for maternal BMI in all target samples. We therefore calculated lassosum PRS for all ALSPAC and BiB mothers with non-transmitted allele data available. To avoid overfitting one would ideally use an independent validation dataset to optimise the values of $\lambda$ and $s$, before applying these optimised values to calculate PRS in the target datasets. Overfitting does not appear to be a problem for our PRS however, because (i) for ALSPAC and BiB the best values for $\lambda$ and $s$ were similar, and (ii) PRS $R^2$ was similar for $s$ = 0.2 and $s$ = 0.5 at values of $\lambda$ close to the best $\lambda$, as shown in the plots immediately below.

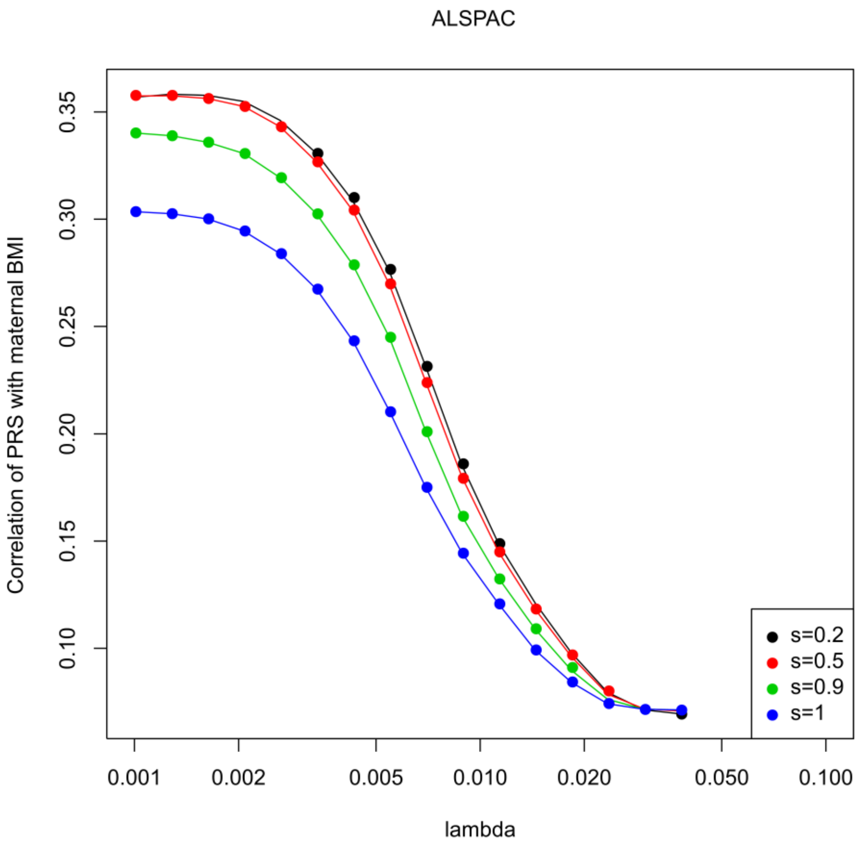

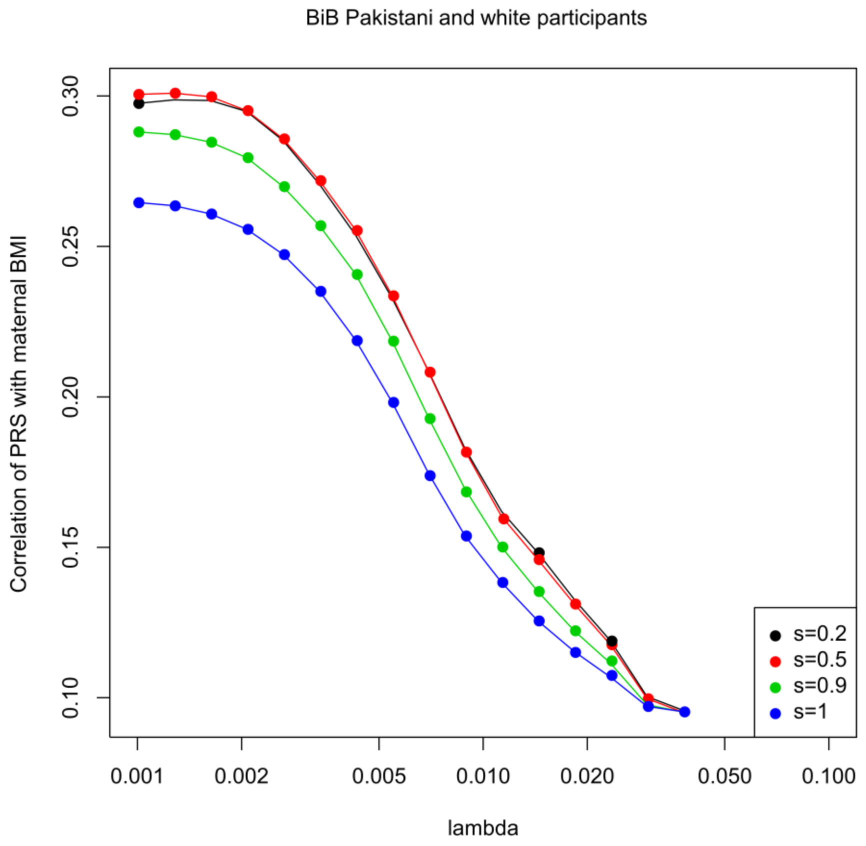
BiB Pakistani and white participants
Correlation of PRS with maternal BMI
0.10
0.15
0.20
0.25
0.30
0.001
0.002
0.005
0.010
0.020
0.050
0.100
lambda
s=0.2
s=0.5
s=0.9
s=1

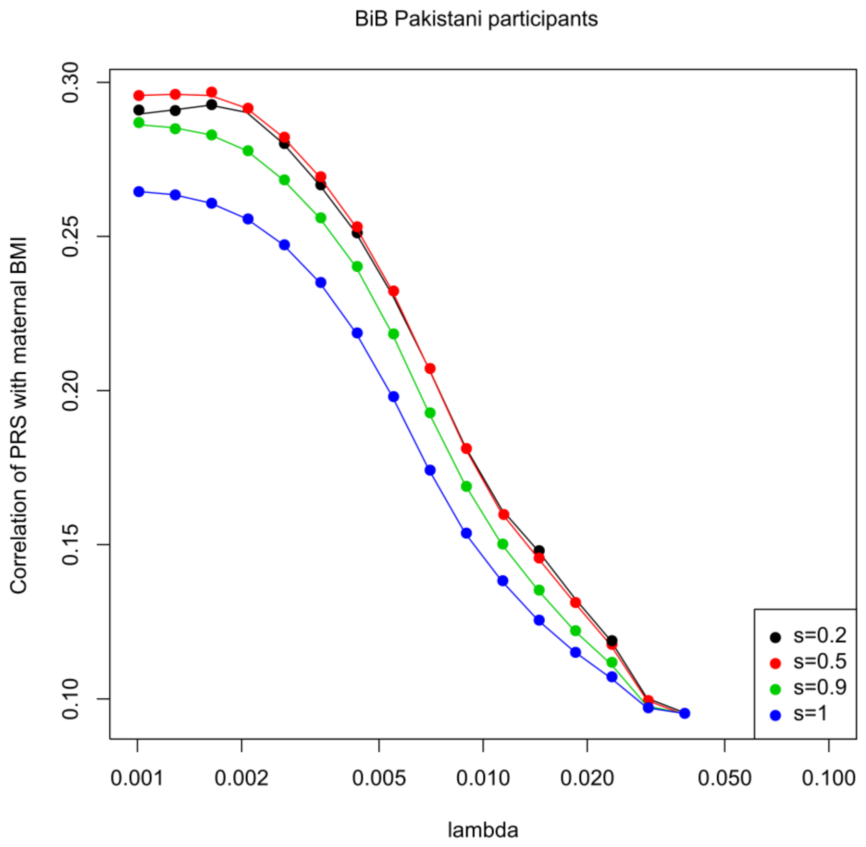
BiB Pakistani participants
Correlation of PRS with maternal BMI
0.10
0.15
0.20
0.25
0.30
0.001
0.002
0.005
0.010
0.020
0.050
0.100
lambda
s=0.2
s=0.5
s=0.9
s=1

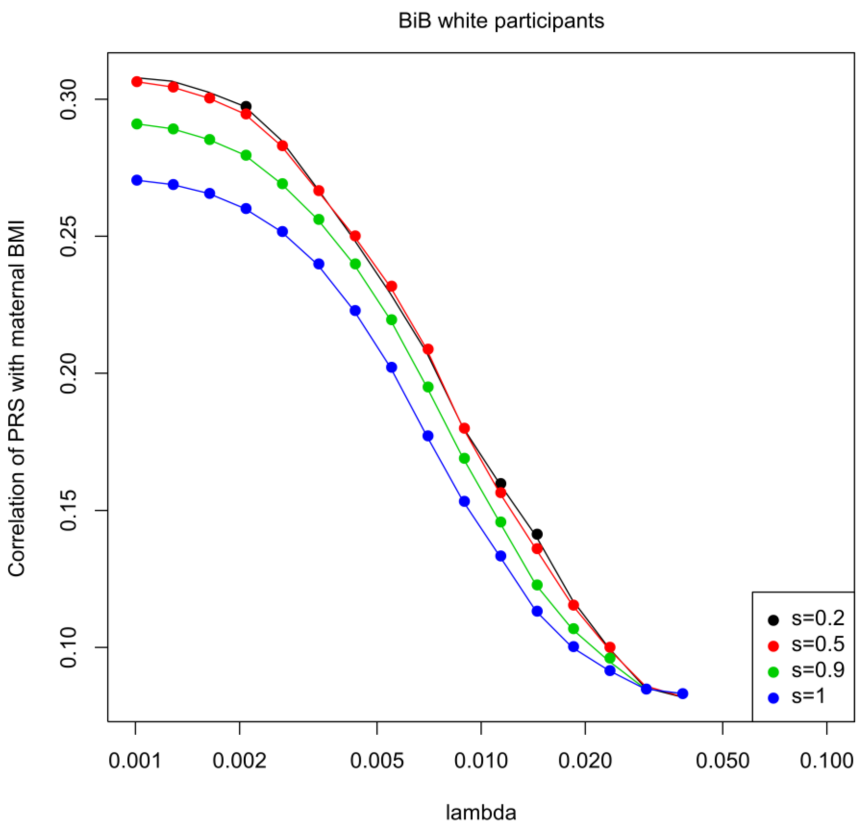

**Supplementary information S12: Selection of SNPs for alternative IVs**

We repeated MR analyses with IVs calculated from fewer SNPs than the lassosum PRS, in order to explore whether MR estimates changed as a function of the number of SNPs included in the IV. A priori we would expect the risk of pleiotropic bias to increase as the number of SNPs included in the IV increases, and that the magnitude of pleiotropic bias might change as the effect size distribution of the included SNPs changes (for example the lassosum PRS includes many SNPs with small effects). We used four additional IVs:

(i) the SNP rs9939609 at the *FTO* locus (this SNP was the first to be identified as associated with BMI (42), and *FTO* is the locus for which there is currently the strongest evidence for association with BMI; rs9939609 was also used as an IV in a previous MR study investigating the effect of maternal BMI on offspring adiposity (43))

(ii) a PRS calculated from 32 SNPs associated with BMI at genome wide significance (GWS) in a 2010 GWAS by Speliotes *et al*. (44) (*N* up to 249,796 individuals); this PRS was also used as an IV in a previous MR study investigating the effect of maternal BMI on offspring adiposity (7)

(iii) a PRS calculated from 94 SNPs associated with BMI at GWS in a 2015 GWAS by Locke *et al*. (37) (*N* up to 339,224 individuals); this PRS was also used as an IV in a previous MR study investigating the effect of maternal BMI on offspring adiposity (7)

(iv) a PRS calculated from 656 SNPs associated with BMI at GWS in a 2018 GWAS by Yengo *et al*. (38) (*N* up to 795,640 individuals)

For the Locke *et al*. GWAS we excluded three SNPs for which there was only strong evidence for association with BMI in men (given that in our MR analyses the exposure is maternal BMI), and for the Yengo *et al*. GWAS we included 656 SNPs identified as primary GWS associations by the authors (based on *P*-value for association <1e-8 in single marker regression [this more conservative threshold is appropriate when using SNPs imputed to the Haplotype Reference Consortium (HRC) or 1000 genomes imputation reference panels (45)]). The lists of GWS SNPs from the three GWAS that we included in our analyses are given in the table immediately below.

| GWAS | SNPs taken forward into further analyses | |
|---|---|---|
| | ***N* SNPs** | **rsIDs** |
| Speliotes *et al.* | 32 | rs2815752, rs1514175, rs1555543, rs543874, rs2867125, rs713586, rs887912, rs2890652, rs13078807, rs9816226, rs10938397, rs13107325, rs2112347, rs4836133, rs206936, rs987237, rs10968576, rs4929949, rs10767664, rs3817334, rs7138803, rs4771122, rs11847697, rs10150332, rs2241423, rs12444979, rs7359397, rs1558902, rs571312, rs29941, rs2287019, rs3810291 |
| Locke *et al.* | 94 | rs11583200, rs977747, rs657452, rs3101336, rs12566985, rs12401738, rs11165643, rs543874, rs2820292, rs17024393, rs13021737, rs10182181, rs11126666, rs1016287, rs11688816, rs2121279, rs1460676, rs1528435, rs17203016, rs7599312, rs6804842, rs2365389, rs3849570, rs13078960, rs16851483, rs1516725, rs10938397, rs17001654, rs13107325, rs11727676, rs2112347, rs7715256, rs205262, rs2033529, rs2207139, rs9400239, rs9374842, rs13201877, rs13191362, rs1167827, rs2245368, rs9641123, rs6465468, rs17405819, rs2033732, rs4740619, rs10968576, rs6477694, rs1928295, rs10733682, rs7899106, rs17094222, rs11191560, rs7903146, rs4256980, rs11030104, rs2176598, rs3817334, rs12286929, rs7138803, rs11057405, rs12016871, rs12429545, rs9540493, rs1441264, rs10132280, rs12885454, rs11847697, rs7141420, rs3736485, rs16951275, rs7164727, rs758747, rs12446632, rs2650492, rs3888190, rs4787491, rs9925964, rs2080454, rs1558902, rs9914578, rs1000940, rs12940622, rs1808579, rs7239883, rs7243357, rs6567160, rs17724992, rs29941, rs2075650, rs2287019, rs3810291, rs6091540, rs2836754 |
| Yengo *et al.* | 656 | rs1000096, rs1000940, rs1003081, rs1006353, rs10066835, rs10083803, rs10110727, rs10118701, rs10132280, rs10146527, rs10182181, rs1020548, rs10211055, rs10269783, rs1035010, rs1037587, rs1038088, rs10408013, rs10438964, rs1045411, rs10456637, rs10460960, rs1048932, rs10497807, rs10499276, rs10511093, rs1057452, rs10733051, rs10744146, rs10745785, rs10754210, rs1075901, rs10772055, rs10773049, rs10779751, rs10797115, rs10818810, rs10818938, rs10830452, rs10832778, rs10838122, rs10840606, rs10842240, rs10867256, rs10876418, rs10878946, rs10883553, rs10883759, rs10886017, rs10887578, rs10915840, rs10920678, rs10923724, rs10929925, rs10930641, rs10935143, rs10938397, rs10953620, rs10962549, rs10971712, rs10989568, rs11066301, rs1106908, rs11074446, rs11075489, rs11075986, rs11079849, rs11115176, rs11118308, rs11128760, rs11150911, rs11208662, rs11250076, rs11251352, rs11496125, rs11577179, rs11583122, rs11600990, rs11611246, rs11611496, rs11629783, rs11635675, rs11642001, rs11659764, rs11668301, rs11672660, rs11736228, rs11753081, rs11773362, rs1178060, rs11781222, rs11781699, rs11783247, rs11790280, rs11792069, rs11855853, rs11866815, rs11889536, rs11915371, rs11945861, rs11971041, rs12033257, rs12035149, rs12035349, rs12041258, rs12042908, rs12042959, rs12044597, rs12098284, rs12101393, rs12147845, rs1218822, rs12189178, rs12215331, rs1229057, rs12364470, rs12411886, rs12420725, rs12422552, rs12429545, rs12439798, rs12446632, rs12448257, rs12448738, rs12479233, rs12488237, rs12514473, rs12564992, rs12574668, rs12575252, rs12577642, rs12587412, rs12593036, rs12595158, rs12595749, rs12602912, rs12609744, rs12631248, rs12655756, rs12675063, rs12680842, rs12694021, rs12705987, rs12731372, rs12776880, rs1285245, rs12885454, rs12888545, rs12922346, rs12939549, rs1296328, rs12964689, rs13021737, rs13041173, rs13047416, rs13107325, rs13110266, rs13174863, rs13191362, rs13201877, rs1320903, rs13209872, rs13209968, rs13227433, rs13227658, rs1323068, rs13240600, rs1324110, rs13247665, rs13263601, rs13290794, rs13292976, rs13321566, rs13329567, rs1333423, rs13417156, rs13432055, rs1345148, rs1358808, rs1362910, rs1365466, rs1371108, rs1375561, rs1394, rs1405348, rs1412235, rs1431659, rs1436343, rs1437929, rs1451533, rs1452075, rs1454687, rs1455137, rs1477887, rs1512065, rs1522569, rs1544459, rs1549293, rs1552717, rs1554193, rs156151, rs159032, rs1608445, rs1625427, rs1658820, rs1668633, rs16828086, rs16851483, rs16906845, rs16907751, rs16932761, rs16966801, rs1700137, rs17014375, rs17035438, rs17056301, rs17094222, rs17105272, rs17203016, rs17207196, rs17238110, rs1730859, rs17327461, rs17391694, rs17446091, rs17522122, rs17531363, rs17538472, rs17551974, rs17608150, rs17636031, rs17681451, rs17681708, rs17724992, rs17757975, rs17806379, rs17820822, rs1782507, rs1787267, rs1789165, rs1814170, rs1819844, rs1852006, rs1853639, rs1862451, rs1865341, rs1866510, rs1884389, rs1884897, rs1899689, rs1899951, rs1903579, rs1909586, rs1927790, rs1928295, rs1937684, rs1941697, rs1942866, rs1951455, rs1956151, rs1973993, rs1982441, rs1983864, rs2000746, rs2010281, rs2012927, rs2030342, rs2033529, rs2075205, rs2075650, rs208015, rs2080454, rs2100814, rs2122042, rs2124499, rs2143253, rs2154297, rs215632, rs2163188, rs2170382, rs2174307, rs217671, rs2185027, rs2190788, rs2192158, rs2228213, rs2229616, rs223051, rs2230590, rs2275426, rs2282231, rs2283093, rs2293605, rs2304130, rs2304607, rs2307022, rs2307111, rs2356865, rs2357760, rs2365389, rs2367112, rs2425241, rs2425857, rs2429150, rs2479958, rs248139, rs2481665, rs2491864, rs2504674, rs2516739, rs2543132, rs25832, rs2590942, rs2605603, rs2611742, rs2619976, rs262956, rs2631681, rs2634047, rs2635727, rs2680648, rs2710323, rs2721965, rs2731222, rs2733287, rs273504, rs2768950, rs2781668, rs2814992, rs2820311, rs28350, rs2836961, rs2838006, rs284227, rs2850969, rs2866816, rs2875762, rs2890652, rs29938, rs310618, rs3134353, rs3134438, rs326889, rs329124, rs329651, rs331949, rs340025, rs355777, rs368863, rs3731544, rs3736485, rs3739733, rs3751813, rs3753549, rs3781099, rs3800229, rs3800649, rs3803286, rs3807566, rs3808434, rs3810291, rs3814883, rs3822683, rs3826705, rs3829849, rs3849570, rs3850422, rs3851083, rs3887080, rs3902840, rs3902951, rs3915844, rs39654, rs40067, rs40245, rs4077093, rs4148155, rs419261, rs4273371, rs4278019, rs427943, rs4284600, rs4303732, rs4307239, rs4366093, rs4372296, rs4372836, rs450231, rs4516268, rs4518345, rs453520, rs4624596, rs4639527, rs4653017, rs4670626, rs4671328, rs4671358, rs4673553, rs4718966, rs4722398, rs4722672, rs4725984, rs4737183, rs4740383, rs4744275, rs4759073, rs4759228, rs4764949, rs4766710, rs4783241, rs4796243, rs4814512, rs4820408, rs4841659, rs4858193, rs4864201, rs487060, rs4889782, rs4906908, rs4936175, rs4936671, rs4969387, rs4981693, rs498240, rs4985155, rs4986044, rs5396, rs543874, rs555267, rs559267, rs577525, rs580438, rs587230, rs6011457, rs6019482, rs6121381, rs6138482, rs6142096, rs621042, rs6235, rs6265, rs629443, rs6442101, rs6443750, rs6448587, rs6449531, rs6461115, rs6463489, rs6474945, rs6477694, rs651548, rs6545714, rs6548221, rs6564360, rs6569648, rs6577584, rs6587552, rs6595205, rs6606686, rs663129, rs6690764, rs6700816, rs6700838, rs6710871, rs6711584, rs6713781, rs6720868, rs6738445, rs6764533, rs676749, rs6767619, rs6786582, rs6804181, rs6804842, rs6827083, rs6843738, rs6877851, rs6888159, rs6900723, rs6921533, rs6938239, rs6963840, rs6968554, rs7006629, rs7024334, rs7042372, rs7083450, rs7084454, rs7102454, rs711347, rs7117238, rs7120873, rs7123876, rs7124442, rs7124681, rs7131262, rs7133378, rs7138803, rs7144011, rs7147503, rs7164727, rs7181498, rs7181610, rs7187776, rs7195386, rs719802, rs7200919, rs7206608, rs7209235, rs7211567, rs7217226, rs7239114, rs7243357, rs7334078, rs733594, rs7377083, rs740157, rs7444298, rs7531118, rs7531656, rs7535528, rs7550711, rs7556169, rs7557796, rs7560871, rs7561278, rs7564679, rs756717, rs757318, rs7598402, rs7599312, rs7600699, rs7601895, rs7607351, rs7607369, rs761423, rs7615297, rs762147, rs7640424, rs7674623, rs7683836, rs7694732, rs769674, rs7710595, rs7711753, rs7715256, rs7716275, rs7730004, rs7730898, rs7748777, rs775731, rs7760082, rs7784465, rs7805441, rs7827182, rs7844647, rs785278, rs7865157, rs7869771, rs7871866, rs7874154, rs7893571, rs7899106, rs7903146, rs7919, rs7941030, rs7948120, rs7968230, rs7973955, rs7975187, rs799449, rs8016771, rs8016859, rs8033510, rs8036040, rs8047395, rs8070454, rs8071182, rs8075273, rs8081039, rs8089514, rs8092503, rs8094523, rs8095404, rs8097544, rs8102137, rs8123881, rs823074, rs825680, rs827092, rs845084, rs847747, rs849135, rs853679, rs868554, rs879620, rs886444, rs889398, rs892261, rs895330, rs900144, rs901630, rs902695, rs903959, rs905938, rs9077, rs925018, rs925421, rs9291467, rs9294260, rs929641, rs9299, rs930295, rs9332817, rs934515, rs9349239, rs9362662, rs9367368, rs9394312, rs9426003, rs9463175, rs946526, rs9475173, rs9507983, rs9527706, rs9530843, rs9538141, rs954018, rs9540493, rs9571687, rs9595908, rs9603697, rs9615905, rs9630985, rs9675376, rs967605, rs9688431, rs9714342, rs972283, rs972540, rs977540, rs9787495, rs980329, rs9806058, rs9809534, rs9814633, rs9816226, rs9838283, rs987237, rs9905991, rs9922708, rs9927848, rs9931967, rs9951893, rs998584, rs998732 |

We checked whether rs9939609 and each of the SNPs listed in the table above were available in the imputed genotype data for each cohort (ALSPAC or BiB) after application of the filters described in **Supplementary information S9**. Where SNPs were unavailable we used the Single Nucleotide Polymorphisms Annotator (SNiPA) proxy search tool (46) to identify a suitable proxy SNP that was in LD with the index SNP ($r^2$ >0.8) in the appropriate 1000 Genomes Project phase 3 v5 reference panel ("European" for ALSPAC and BiB White Europeans and "South Asian" for BiB South Asians) and was also available in our UKB + GIANT meta-analysed GWAS summary statistics. We then applied clumping to each set of SNPs in order to ensure that each SNP was an independent instrument, using the PLINK command *--clump-kb 10000 --clump-r2 0.001 --clump-p1 1 --clump-p2 1* and the samples of ALSPAC, BiB South Asians or BiB White Europeans as LD references. The final number of SNPs available for calculation of each IV is given in the table immediately below.

| **Cohort** | ***N* snps** | **GWS IV** | | |
|---|---|---|---|---|
| | | **Speliotes *et al.*** | **Locke *et al.*** | **Yengo *et al.*** |
| ALSPAC | Before SNiPA proxy search | 32 | 94 | 656 |
| | After SNiPA proxy search | 31 | 91 | 616 |
| | After clumping | 31 | 87 | 497 |
| BiB South Asians | Before SNiPA proxy search | 32 | 94 | 656 |
| | After SNiPA proxy search | 30 | 91 | 633 |
| | After clumping | 29 | 82 | 446 |
| BiB White Europeans | Before SNiPA proxy search | 32 | 94 | 656 |
| | After SNiPA proxy search | 31 | 92 | 633 |
| | After clumping | 31 | 86 | 453 |

We calculated each GWS PRS as a weighted sum of BMI increasing maternal non-transmitted alleles at the relevant SNPs using the SNP effects from our UKB + GIANT GWAS meta-analysis as weights. The SNPs used for PRS calculation in each sample are listed in the tables immediately below.

| GWAS | SNPs used to calculate GWS IVs in ALSPAC | |
|---|---|---|
| | ***N* SNPs** | **rsIDs** |
| Speliotes *et al.* | 31 | rs543874, rs2815752, rs1514175, rs1555543, rs2030323, rs3817334, rs4929949, rs7138803, rs4771122, rs11847697, rs10150332, rs2241423, rs12444979, rs7359397, rs1421085, rs571312, rs29941, rs2287019, rs3810291, rs2890652, rs713586, rs887912, rs2867125, rs7647305, rs13078807, rs13107325, rs10938397, rs2112347, rs206936, rs987237, rs10968576 |
| Locke *et al.* | 87 | rs17024393, rs543874, rs2820292, rs977747, rs657452, rs3101336, rs12566985, rs12401738, rs11165643, rs17113297, rs11191560, rs7903146, rs7899106, rs12286929, rs11030104, rs2176598, rs3817334, rs4929928, rs11057405, rs7138803, rs12429545, rs9540493, rs1441264, rs10132280, rs12885454, rs11847697, rs7141420, rs3736485, rs16951275, rs7164727, rs12446632, rs3888190, rs9925964, rs758747, rs2080454, rs1421085, rs4061660, rs1000940, rs12940622, rs1808579, rs7239883, rs7243357, rs6567160, rs17724992, rs29941, rs2075650, rs2287019, rs3810291, rs2121279, rs1460676, rs1528435, rs17203016, rs7599312, rs10182181, rs1016287, rs11688816, rs13021737, rs6091540, rs2836754, rs16851483, rs1516725, rs6804842, rs2365389, rs3849570, rs13078960, rs13107325, rs10938397, rs17001561, rs7715256, rs2112347, rs9400239, rs9374842, rs3904531, rs13191362, rs205262, rs2033529, rs2207139, rs1167827, rs10464483, rs6465468, rs17405819, rs2033732, rs6477694, rs1928295, rs10733682, rs4740619, rs10968576 |
| Yengo *et al.* | 497 | rs1730859, rs11185092, rs7550711, rs10779751, rs12731372, rs10923724, rs6587552, rs905938, rs10733051, rs12044597, rs12564992, rs543874, rs10920678, rs12041258, rs10754210, rs2820311, rs9077, rs823074, rs17014375, rs11118308, rs10915840, rs967605, rs12042959, rs3753549, rs7535528, rs2787120, rs11583122, rs2282231, rs2275426, rs6700838, rs7531656, rs2481665, rs6577584, rs2590942, rs12042908, rs17391694, rs2154297, rs284227, rs7556169, rs6690764, rs1973993, rs4372296, rs17113297, rs10883553, rs7083450, rs12411886, rs7903146, rs10886017, rs845084, rs7893571, rs11593937, rs7084454, rs11251352, rs3781099, rs3851083, rs10762499, rs6479905, rs12098284, rs7899106, rs10749537, rs2631681, rs577525, rs719802, rs1048932, rs12420725, rs1037587, rs1003081, rs583564, rs7941030, rs1625427, rs4936175, rs900144, rs2051772, rs10840606, rs6265, rs1782507, rs223051, rs10838122, rs7928523, rs7124681, rs11600990, rs7102454, rs1789165, rs7123876, rs7117238, rs7113874, rs10830452, rs2605603, rs4764949, rs11611496, rs17696736, rs4766710, rs7973955, rs3887080, rs7133378, rs7968230, rs10744146, rs621042, rs2429150, rs4569092, rs10876418, rs2733287, rs7138803, rs4077093, rs4759073, rs7975187, rs1819844, rs10878946, rs11115176, rs2731222, rs11611246, rs10745785, rs651548, rs2479958, rs1218822, rs9507983, rs1045411, rs9595908, rs9603697, rs12429545, rs9538141, rs9540493, rs9571687, rs629443, rs1668633, rs9530843, rs1927790, rs7334078, rs12147845, rs7147503, rs3803286, rs10132280, rs12885454, rs8003383, rs17522122, rs11847892, rs1956151, rs12587412, rs217671, rs17105272, rs7144011, rs12888545, rs1951455, rs3850422, rs4906908, rs4284600, rs1036949, rs8036040, rs12439798, rs3736485, rs340025, rs8033510, rs12595158, rs11635675, rs17200970, rs13329567, rs7164727, rs11855853, rs12595749, rs12593036, rs12101386, rs7181498, rs4985155, rs12446632, rs11074446, rs2516739, rs9927848, rs7195386, rs7187776, rs3814883, rs12448257, rs11866815, rs2080454, rs9922708, rs11076022, rs12448738, rs11075489, rs10083803, rs7200919, rs889398, rs11642001, rs6564360, rs10492861, rs977540, rs1075901, rs4516268, rs4986044, rs7217226, rs1038088, rs7211567, rs1106908, rs4796243, rs8070454, rs208015, rs1000940, rs8075273, rs12602912, rs2619976, rs7209235, rs12939549, rs9905991, rs17681708, rs8097544, rs12964689, rs1941697, rs1365466, rs555267, rs954018, rs10438964, rs8092503, rs7243357, rs663129, rs9951893, rs2012927, rs12604935, rs11150911, rs673429, rs1608445, rs12609744, rs273504, rs757318, rs998732, rs2304130, rs8102137, rs2866816, rs11668301, rs10408013, rs29938, rs7254272, rs3826705, rs2075650, rs11672660, rs3810291, rs4303732, rs1451533, rs902695, rs17551974, rs6710871, rs7560871, rs453520, rs2119753, rs9646758, rs6738445, rs10930641, rs9630985, rs4850808, rs7564679, rs12694021, rs17203016, rs4673553, rs7599312, rs7607369, rs11889536, rs10211055, rs6720868, rs10182181, rs4372836, rs6548221, rs17327461, rs4670626, rs4639527, rs17035438, rs7561278, rs930295, rs3198123, rs13432055, rs4671328, rs6545714, rs1559625, rs10929925, rs13417156, rs13021737, rs7607351, rs934515, rs1371108, rs7557796, rs1884389, rs8123881, rs4814512, rs6138482, rs613309, rs6142096, rs2143253, rs2425857, rs6019482, rs17806379, rs1512065, rs559267, rs6010786, rs1884897, rs762147, rs9983113, rs2836961, rs2838006, rs427943, rs4820408, rs9615905, rs1436343, rs7640424, rs17681451, rs2868975, rs4624596, rs1899951, rs1909586, rs1320903, rs580438, rs10935143, rs6786582, rs16851483, rs171390, rs6805114, rs827092, rs11128760, rs5396, rs39654, rs262956, rs7647305, rs6764533, rs4858193, rs6804842, rs9814633, rs10460960, rs2230590, rs12488237, rs2365389, rs1452075, rs11915371, rs775731, rs3849570, rs2122042, rs4857329, rs3915844, rs13107325, rs6843738, rs326889, rs7694732, rs4864201, rs1296328, rs769671, rs331949, rs1455137, rs7663212, rs6827083, rs13110266, rs17538472, rs1522569, rs7683836, rs1477887, rs1323068, rs9291467, rs6448587, rs1345148, rs1000096, rs10938397, rs784944, rs2192158, rs925421, rs11945861, rs7674623, rs4148155, rs13103126, rs7710595, rs40067, rs4895141, rs7711753, rs329124, rs13174863, rs2190788, rs7715256, rs17056301, rs248139, rs7730898, rs4518345, rs12189178, rs6449531, rs2367112, rs25832, rs2307111, rs368863, rs12514473, rs7444298, rs2304607, rs2611742, rs156153, rs3800229, rs2357760, rs2228213, rs1159974, rs6569648, rs9367368, rs2781668, rs3904531, rs2185027, rs10499276, rs487060, rs13191362, rs11753081, rs7760082, rs6900723, rs853679, rs498240, rs2814992, rs17757975, rs2033529, rs9349239, rs998584, rs987237, rs9475173, rs1020548, rs6921533, rs9688431, rs9294260, rs1853639, rs9362662, rs9463175, rs901630, rs11496125, rs10953620, rs13227658, rs1899689, rs2283093, rs972283, rs7811342, rs11773362, rs4725984, rs6968554, rs6461115, rs2282888, rs4307239, rs11971041, rs849135, rs4722398, rs215632, rs1229057, rs799449, rs10269783, rs3807566, rs6463489, rs7784465, rs4718966, rs1167827, rs740157, rs1852006, rs6963840, rs13247665, rs13240600, rs3134358, rs11783247, rs2721965, rs11781699, rs12675099, rs16906845, rs13263601, rs7357604, rs4366093, rs11781222, rs17446091, rs1982441, rs1362910, rs7844647, rs1658820, rs4737183, rs16932761, rs1431659, rs2170382, rs1405348, rs16907751, rs733594, rs1700137, rs12680842, rs450231, rs10118701, rs10989568, rs7024334, rs6477694, rs17820822, rs1928295, rs7865157, rs10818810, rs10818938, rs13292976, rs4734, rs4740383, rs11790280, rs6474945, rs10962549, rs1412234, rs10971712, rs13290794, rs7042372, rs1928538, rs10867256, rs1865341, rs7852822, rs10797115, rs7869771, rs10992876 |

| GWAS | SNPs used to calculate GWS IVs in BiB South Asians | |
|---|---|---|
| | ***N*** **SNPs** | **rsIDs** |
| Speliotes *et al.* | 29 | rs543874, rs2815752, rs1555543, rs2030323, rs3817334, rs4929949, rs7138803, rs4771122, rs11847697, rs10150332, rs2241423, rs12444979, rs7359397, rs1421085, rs571312, rs29941, rs2287019, rs3810291, rs2890652, rs713586, rs887912, rs2867125, rs13078807, rs13107325, rs10938397, rs2112347, rs206936, rs987237, rs10968576 |
| Locke *et al.* | 82 | rs17024393, rs543874, rs2820292, rs977747, rs657452, rs3101336, rs12401738, rs11165643, rs17094222, rs7903146, rs7899106, rs12286929, rs11030104, rs2176598, rs3817334, rs10840100, rs11057405, rs7138803, rs12429545, rs9540493, rs1441264, rs10132280, rs12885454, rs11847697, rs7141420, rs3736485, rs16951275, rs7164727, rs12446632, rs3888190, rs9925964, rs758747, rs2080454, rs1421085, rs4061660, rs1000940, rs12940622, rs1808579, rs7239883, rs7243357, rs6567160, rs17724992, rs29941, rs2287019, rs3810291, rs2121279, rs1460676, rs1528435, rs17203016, rs7599312, rs10182181, rs1016287, rs11688816, rs13021737, rs6091540, rs2836754, rs16851483, rs1516725, rs6804842, rs2365389, rs13078960, rs13107325, rs11727676, rs10938397, rs7715256, rs2112347, rs9400239, rs9374842, rs13201877, rs13191362, rs205262, rs2207139, rs1167827, rs2245368, rs10464483, rs6465468, rs17405819, rs2033732, rs6477694, rs10733682, rs4740619, rs10968576 |
| Yengo *et al.* | 446 | rs1730859, rs11185092, rs17531363, rs7550711, rs12033257, rs10779751, rs12731372, rs10923724, rs6587552, rs905938, rs10733051, rs12044597, rs761423, rs12564992, rs543874, rs10920678, rs10754210, rs2820311, rs9077, rs823074, rs17014375, rs11118308, rs10915840, rs967605, rs3753549, rs7535528, rs2787120, rs4653017, rs11583122, rs2282231, rs2275426, rs7531656, rs2481665, rs6577584, rs7531118, rs12042908, rs17391694, rs2154297, rs7556169, rs6690764, rs1973993, rs2030342, rs17094222, rs10883553, rs9787495, rs7903146, rs10886017, rs845084, rs17636031, rs11593937, rs7084454, rs11251352, rs3781099, rs3851083, rs10762499, rs6479905, rs12098284, rs7899106, rs2631681, rs577525, rs719802, rs1048932, rs12420725, rs1037587, rs583564, rs3134438, rs1625427, rs4936175, rs329651, rs12364470, rs1557765, rs10840606, rs6265, rs7948120, rs1782507, rs223051, rs10838122, rs7928523, rs7124681, rs11600990, rs7102454, rs587230, rs7117238, rs7113874, rs10830452, rs2605603, rs4764949, rs11611496, rs17608150, rs11066301, rs4766710, rs7973955, rs3887080, rs7133378, rs10773049, rs7968230, rs10744146, rs621042, rs4569092, rs10876418, rs7138803, rs4077093, rs4759073, rs1819844, rs10878946, rs11115176, rs2731222, rs11611246, rs2479958, rs1218822, rs9507983, rs9595908, rs9603697, rs12429545, rs9538141, rs892261, rs9540493, rs9571687, rs629443, rs1668633, rs9530843, rs1927790, rs7334078, rs12147845, rs7147503, rs3803286, rs10132280, rs12885454, rs8003383, rs7144747, rs1956151, rs12587412, rs217671, rs3902951, rs17105272, rs7144011, rs12888545, rs1951455, rs4284600, rs1036949, rs8036040, rs3736485, rs340025, rs17238110, rs11635675, rs17200970, rs13329567, rs7164727, rs11855853, rs12593036, rs12101386, rs7181498, rs4985155, rs12446632, rs9931967, rs2516739, rs1862451, rs7187776, rs1549293, rs12448257, rs11866815, rs879620, rs2080454, rs9922708, rs11076022, rs12448738, rs11075489, rs2307022, rs889398, rs756717, rs862227, rs977540, rs1075901, rs4516268, rs4986044, rs1038088, rs7211567, rs1106908, rs4796243, rs16966801, rs208015, rs11079849, rs1000940, rs8075273, rs12602912, rs2619976, rs7209235, rs8081039, rs1696757, rs12939549, rs9905991, rs17681708, rs8097544, rs12964689, rs1941697, rs555267, rs954018, rs7239114, rs8092503, rs7243357, rs663129, rs2012927, rs11150911, rs673429, rs12609744, rs273504, rs757318, rs8102137, rs11668301, rs3826705, rs2075650, rs11672660, rs3810291, rs6711584, rs1451533, rs902695, rs2890652, rs7560871, rs453520, rs2119753, rs9646758, rs6738445, rs10930641, rs9630985, rs12472359, rs17203016, rs4673553, rs7599312, rs7607369, rs10211055, rs6720868, rs10182181, rs4372836, rs17327461, rs4670626, rs4639527, rs17035438, rs930295, rs6757907, rs13432055, rs4671328, rs13417156, rs13021737, rs7607351, rs934515, rs1371108, rs7557796, rs1884389, rs8123881, rs4814512, rs6138482, rs654750, rs6142096, rs2425241, rs6019482, rs17806379, rs559267, rs6010784, rs310618, rs1884897, rs2836767, rs2836961, rs2838006, rs427943, rs4820408, rs9615905, rs1436343, rs13321566, rs7640424, rs17681451, rs9817840, rs4624596, rs1899951, rs1909586, rs1320903, rs10935143, rs6786582, rs16851483, rs171390, rs6805114, rs827092, rs5396, rs39654, rs6443750, rs262956, rs6764533, rs4858193, rs6804842, rs2230590, rs2680648, rs12488237, rs2365389, rs1452075, rs11915371, rs775731, rs10511093, rs2122042, rs4857329, rs3915844, rs2850969, rs13107325, rs326889, rs7694732, rs4864201, rs1296328, rs331949, rs7663212, rs6827083, rs13110266, rs1522569, rs7683836, rs1477887, rs1323068, rs9291467, rs6448587, rs1000096, rs10938397, rs784944, rs2192158, rs11945861, rs7674623, rs4148155, rs1461741, rs7710595, rs40067, rs4895141, rs7711753, rs329124, rs7716275, rs2190788, rs10066835, rs7715256, rs17056301, rs7730898, rs4518345, rs7730004, rs12189178, rs2367112, rs25832, rs2307111, rs368863, rs12514473, rs7444298, rs2304607, rs7713317, rs2611742, rs156153, rs3800229, rs2357760, rs2228213, rs6569648, rs9367368, rs2781668, rs13201877, rs2185027, rs10499276, rs487060, rs13191362, rs11753081, rs7760082, rs6900723, rs498240, rs419261, rs2814992, rs7748777, rs998584, rs987237, rs9475173, rs1020548, rs6921533, rs9688431, rs9294260, rs1853639, rs9362662, rs901630, rs11496125, rs13227658, rs12705986, rs1899689, rs972283, rs3800649, rs7811342, rs11773362, rs4725984, rs6968554, rs6461115, rs2282888, rs4307239, rs849135, rs4722398, rs215632, rs799449, rs10269783, rs3807566, rs7784465, rs1035010, rs17207196, rs1852006, rs6963840, rs13247665, rs13240600, rs2721965, rs11781699, rs12675099, rs13263601, rs12549680, rs4366093, rs11781222, rs17446091, rs1982441, rs1362910, rs7844647, rs1658820, rs4737183, rs16932761, rs1431659, rs2170382, rs1405348, rs16907751, rs733594, rs2634044, rs1700137, rs1394, rs12680842, rs10118701, rs10989568, rs1928295, rs10818810, rs3829849, rs3902840, rs4734, rs4740383, rs11790280, rs6474945, rs10962549, rs7874154, rs10968576, rs10971712, rs13290794, rs7042372, rs1928538, rs10867256, rs1865341, rs3739733, rs10797115, rs4744275 |

| GWAS | SNPs used to calculate GWS IVs in BiB White Europeans | |
|---|---|---|
| | ***N* SNPs** | **rsIDs** |
| Speliotes *et al.* | 31 | rs543874, rs2815752, rs1514175, rs1555543, rs2030323, rs3817334, rs4929949, rs7138803, rs4771122, rs11847697, rs10150332, rs2241423, rs12444979, rs7359397, rs1421085, rs571312, rs29941, rs2287019, rs3810291, rs2890652, rs713586, rs887912, rs2867125, rs7647305, rs13078807, rs13107325, rs10938397, rs2112347, rs206936, rs987237, rs10968576 |
| Locke *et al.* | 86 | rs17024393, rs543874, rs2820292, rs977747, rs657452, rs3101336, rs12566985, rs12401738, rs11165643, rs17094222, rs11191560, rs7903146, rs7899106, rs12286929, rs11030104, rs2176598, rs3817334, rs4929928, rs11057405, rs7138803, rs12429545, rs9540493, rs1441264, rs10132280, rs12885454, rs7141420, rs3736485, rs16951275, rs7164727, rs12446632, rs3888190, rs9925964, rs758747, rs2080454, rs1421085, rs4061660, rs1000940, rs12940622, rs1808579, rs7239883, rs7243357, rs6567160, rs17724992, rs29941, rs2075650, rs2287019, rs3810291, rs2121279, rs1460676, rs1528435, rs17203016, rs7599312, rs10182181, rs1016287, rs11688816, rs13021737, rs6091540, rs2836754, rs16851483, rs1516725, rs6804842, rs2365389, rs13078960, rs13107325, rs11727676, rs10938397, rs17001561, rs7715256, rs2112347, rs9400239, rs9374842, rs13201877, rs13191362, rs205262, rs2207139, rs1167827, rs2245368, rs10464483, rs6465468, rs17405819, rs2033732, rs6477694, rs1928295, rs10733682, rs4740619, rs10968576 |
| Yengo *et al.* | 453 | rs1730859, rs7550711, rs12033257, rs10779751, rs12731372, rs6587552, rs905938, rs10733051, rs12044597, rs761423, rs12564992, rs543874, rs10920678, rs12041258, rs10754210, rs2820311, rs823074, rs17014375, rs11118308, rs10915840, rs967605, rs3753549, rs7535528, rs4653017, rs9426003, rs11583122, rs2282231, rs946526, rs6700838, rs7531656, rs2481665, rs6577584, rs7531118, rs12042908, rs17391694, rs2154297, rs7556169, rs6690764, rs1973993, rs17094222, rs10883553, rs10883759, rs12411886, rs7903146, rs10886017, rs845084, rs17636031, rs7893571, rs7084454, rs11251352, rs3781099, rs3851083, rs10762499, rs6479905, rs12098284, rs7899106, rs10749537, rs577525, rs719802, rs1048932, rs1037587, rs583564, rs7941030, rs3134438, rs1625427, rs4936175, rs900144, rs329651, rs12364470, rs2051772, rs10840606, rs6265, rs1782507, rs223051, rs10838122, rs7928523, rs7124681, rs11600990, rs7102454, rs1789165, rs7123876, rs7113874, rs10830452, rs2605603, rs4764949, rs11611496, rs4630352, rs11066301, rs7973955, rs7133378, rs7968230, rs10744146, rs621042, rs2429150, rs4569092, rs10876418, rs2733287, rs7138803, rs4077093, rs4759073, rs7975187, rs1819844, rs10878946, rs11115176, rs2731222, rs11611246, rs651548, rs2479958, rs1218822, rs9507983, rs1045411, rs9595908, rs12429545, rs9538141, rs9540493, rs9571687, rs9530843, rs1927790, rs3803286, rs10132280, rs12885454, rs17522122, rs1956151, rs12587412, rs217671, rs3902951, rs17105272, rs10146527, rs7144011, rs12888545, rs1951455, rs3850422, rs4906908, rs4284600, rs1036949, rs12439798, rs3736485, rs340025, rs17238110, rs13329567, rs7164727, rs11855853, rs12595749, rs12101386, rs7181498, rs4985155, rs12446632, rs2516739, rs9927848, rs7195386, rs7187776, rs1549293, rs12448257, rs11866815, rs879620, rs2080454, rs9922708, rs11076022, rs12448738, rs11075489, rs10083803, rs889398, rs756717, rs862227, rs6564360, rs977540, rs1075901, rs4516268, rs4986044, rs7217226, rs1038088, rs1106908, rs4796243, rs8070454, rs16966801, rs9299, rs11079849, rs1000940, rs8071182, rs8075273, rs12602912, rs2619976, rs8081039, rs12939549, rs8097544, rs12964689, rs1941697, rs1365466, rs555267, rs954018, rs7239114, rs8092503, rs7243357, rs663129, rs9951893, rs2012927, rs11150911, rs673429, rs1608445, rs12609744, rs757318, rs8102137, rs11668301, rs29938, rs7254272, rs2075650, rs11672660, rs3810291, rs4303732, rs1451533, rs902695, rs17551974, rs2890652, rs6710871, rs7560871, rs453520, rs2119753, rs9646758, rs6738445, rs10930641, rs9630985, rs4850808, rs7564679, rs12694021, rs972540, rs4673553, rs7599312, rs11889536, rs10211055, rs10182181, rs6548221, rs17327461, rs4670626, rs17035438, rs7561278, rs930295, rs4671328, rs6545714, rs980329, rs10929925, rs13417156, rs13021737, rs7607351, rs934515, rs1371108, rs7557796, rs1884389, rs8123881, rs4814512, rs613309, rs733320, rs6142096, rs2143253, rs2425857, rs6019482, rs17806379, rs1512065, rs559267, rs6010786, rs310618, rs1884897, rs762147, rs2838006, rs427943, rs4820408, rs9615905, rs1436343, rs7640424, rs17681451, rs4624596, rs1899951, rs1909586, rs1320903, rs580438, rs10935143, rs171390, rs6441080, rs6805114, rs827092, rs39654, rs6443750, rs262956, rs7647305, rs6764533, rs4858193, rs6804842, rs10460960, rs2230590, rs2680648, rs12488237, rs2365389, rs1452075, rs11915371, rs775731, rs3849570, rs2122042, rs4857329, rs3915844, rs13107325, rs6843738, rs326889, rs7694732, rs4864201, rs1296328, rs331949, rs7663212, rs6827083, rs13110266, rs17538472, rs1522569, rs7683836, rs1477887, rs1323068, rs9291467, rs6448587, rs1345148, rs1000096, rs1866510, rs10938397, rs784944, rs2192158, rs925421, rs11945861, rs7674623, rs4148155, rs13103126, rs7710595, rs40067, rs4895141, rs7711753, rs329124, rs13174863, rs2190788, rs10066835, rs7715256, rs17056301, rs248139, rs7730898, rs4518345, rs7730004, rs12189178, rs6449531, rs2367112, rs25832, rs2307111, rs368863, rs12514473, rs7444298, rs2304607, rs159032, rs2611742, rs156153, rs3800229, rs2357760, rs2228213, rs1159974, rs6569648, rs9367368, rs2781668, rs13201877, rs2185027, rs487060, rs13191362, rs7760082, rs6900723, rs419261, rs2814992, rs17757975, rs847747, rs7748777, rs998584, rs987237, rs9475173, rs6921533, rs9688431, rs9294260, rs9362662, rs9463175, rs901630, rs11496125, rs10953620, rs12705986, rs1899689, rs2283093, rs3800649, rs7811342, rs11773362, rs4725984, rs6968554, rs6461115, rs4307239, rs11971041, rs849135, rs4722398, rs215632, rs1229057, rs10269783, rs3807566, rs7784465, rs1035010, rs4718966, rs17207196, rs740157, rs1544459, rs7805441, rs13247665, rs13240600, rs3134358, rs11783247, rs2721965, rs11781699, rs12675099, rs16906845, rs13263601, rs7357604, rs435540, rs4366093, rs11781222, rs17446091, rs1982441, rs1362910, rs7844647, rs1658820, rs4737183, rs16932761, rs1431659, rs2170382, rs1405348, rs16907751, rs733594, rs1700137, rs12680842, rs450231, rs10118701, rs6477694, rs17820822, rs1928295, rs7865157, rs13292976, rs4734, rs4740383, rs11790280, rs6474945, rs10962549, rs1412234, rs10971712, rs13290794, rs1928538, rs10867256, rs1865341, rs3739733, rs10797115, rs7869771 |

**Supplementary information S13: Bootstrapping methods**

We tested for a difference between the MR and MV estimates using a *z*-test, for which we calculated the *z*-statistic using the formula:

$$z = \delta / \sqrt{\mathrm{var}(\delta)},$$

where $\delta$ denotes the difference between the MR and MV estimate. We calculated the variance of $\delta$ as

$$\mathrm{var}(\delta) = \mathrm{var}(MR\ estimate) + \mathrm{var}(OLS\ estimate) - 2\mathrm{cov}(MR\ estimate, OLS\ estimate).$$

For the analyses involving only one sample we estimated the variance of the MR and MV estimates, and their covariance, using nonparametric bootstrapping with 1000 resamples. For the meta-analyses we calculated meta-analysed estimates of the MR effect $\hat{\beta}_{MR}$ using the ratio estimator: $\hat{\beta}_{MR} = \hat{\beta}_{ZY}/\hat{\beta}_{ZX}$, where $\hat{\beta}_{ZY}$ is the meta-analysed estimate of the coefficient from regression of the outcome on the instrumental variable (IV) and $\hat{\beta}_{ZX}$ is the meta-analysed estimate of the coefficient from regression of the exposure on the IV. We estimated the variance of the pooled MR effect using a second order Taylor series approximation (47), having first estimated $\mathrm{cov}(\hat{\beta}_{ZY}, \hat{\beta}_{ZX})$ using nonparametric bootstrapping with 1000 resamples (which we also used to estimate the covariance of the MR and MV estimates). We then calculated $\mathrm{var}(\delta)$ and the *z*-statistic as above.

We compared the *z*-statistics to a standard normal distribution in order to calculate the *P*-values for the difference between MR and MV estimates ($P_{\text{difference}}$), and calculated 95% confidence intervals for the MR estimates as 1.96 times their standard error.

**Supplementary information S14: Meta-analysis heterogeneity statistics**

| Samples meta-analysed | Outcome | IV | IV-outcome association | | IV-exposure association | | Exposure-outcome association | |
|---|---|---|---|---|---|---|---|---|
| | | | $I^2$ (%) | $P_{het}$ | $I^2$ (%) | $P_{het}$ | $I^2$ (%) | $P_{het}$ |
| ALSPAC + BiB (South Asian) + BiB (White European) | BW | FTO | 0.0 | 0.552 | 0.0 | 0.534 | 53.0 | 0.119 |
| | | Speliotes | 58.9 | 0.088 | 24.1 | 0.268 | 53.0 | 0.119 |
| | | Locke | 56.4 | 0.101 | 0.0 | 0.837 | 53.0 | 0.119 |
| | | Yengo | 0.0 | 0.411 | 0.0 | 0.555 | 53.0 | 0.119 |
| | | Lassosum | 0.0 | 0.796 | 79.2 | 0.008 | 53.0 | 0.119 |
| | 1yr BMI | FTO | 75.6 | 0.016 | 6.6 | 0.343 | 32.1 | 0.229 |
| | | Speliotes | 0.0 | 0.414 | 0.0 | 0.379 | 32.1 | 0.229 |
| | | Locke | 0.0 | 0.572 | 0.0 | 0.803 | 32.1 | 0.229 |
| | | Yengo | 0.0 | 0.987 | 0.0 | 0.668 | 32.1 | 0.229 |
| | | Lassosum | 2.3 | 0.359 | 69.8 | 0.036 | 32.1 | 0.229 |
| | 4yr BMI | FTO | 67.7 | 0.045 | 17.8 | 0.296 | 0.0 | 0.582 |
| | | Speliotes | 0.0 | 0.606 | 39.7 | 0.190 | 0.0 | 0.582 |
| | | Locke | 29.1 | 0.244 | 0.0 | 0.915 | 0.0 | 0.582 |
| | | Yengo | 0.0 | 0.462 | 0.0 | 0.593 | 0.0 | 0.582 |
| | | Lassosum | 0.0 | 0.755 | 10.9 | 0.326 | 0.0 | 0.582 |
| BiB (South Asian) + BiB (White European) | BW | FTO | 0.0 | 0.362 | 0.0 | 0.651 | 36.1 | 0.211 |
| | | Speliotes | 79.5 | 0.027 | 26.5 | 0.243 | 36.1 | 0.211 |
| | | Locke | 77.8 | 0.034 | 0.0 | 0.782 | 36.1 | 0.211 |
| | | Yengo | 5.6 | 0.303 | 0.0 | 0.871 | 36.1 | 0.211 |
| | | Lassosum | 0.0 | 0.518 | 53.0 | 0.145 | 36.1 | 0.211 |
| | 1yr BMI | FTO | 87.6 | 0.004 | 0.0 | 0.514 | 0.0 | 0.786 |
| | | Speliotes | 43.4 | 0.184 | 6.9 | 0.300 | 0.0 | 0.786 |
| | | Locke | 0.0 | 0.558 | 0.0 | 0.658 | 0.0 | 0.786 |
| | | Yengo | 0.0 | 0.996 | 0.0 | 0.884 | 0.0 | 0.786 |
| | | Lassosum | 41.2 | 0.192 | 0.0 | 0.397 | 0.0 | 0.786 |
| | 4yr BMI | FTO | 83.6 | 0.014 | 0.0 | 0.585 | 0.0 | 0.326 |
| | | Speliotes | 0.0 | 0.506 | 0.0 | 0.526 | 0.0 | 0.326 |
| | | Locke | 8.0 | 0.297 | 0.0 | 0.980 | 0.0 | 0.326 |
| | | Yengo | 0.0 | 0.866 | 0.0 | 0.730 | 0.0 | 0.326 |
| | | Lassosum | 0.0 | 0.460 | 0.0 | 0.444 | 0.0 | 0.326 |

$P_{het}$: *P*-value from Cochran's *Q* test for heterogeneity of effect size

**Supplementary information S15: Linear mixed models**

We repeated MR analyses using a linear mixed model (LMM) to control for population stratification and cryptic relatedness. We fitted the models for the numerator and denominator of the MR ratio estimator (**Supplementary information S13**) separately, using the *--reml-est-fix* command in the GCTA software package (version 1.91.7beta) (11). The numerator model is specified by the equation:

$$\mathbf{y} = \mathbf{Xb} + \mathbf{g} + \mathbf{e}, \qquad [1]$$

where $\mathbf{y}$ is a vector of offspring phenotypes, $\mathbf{b}$ is a vector of fixed effects which include the maternal non-transmitted allele PRS and 20 genetic PCs, $\mathbf{X}$ is a design matrix, $\mathbf{g}$ is a vector of additive genetic values (the sum of the additive effects of all SNPs) modelled as a random effect, with $\mathbf{g} \sim N(\mathbf{0}, \mathbf{I}\sigma_g^2)$, $\mathbf{I}$ is an identity matrix, $\sigma_g^2$ is the additive genetic variance, $\mathbf{e}$ is a vector of residuals with $\mathbf{e} \sim N(\mathbf{0}, \mathbf{I}\sigma_e^2)$ and $\sigma_e^2$ is the residual variance. The variance-covariance matrix is $\mathrm{var}(\mathbf{y}) = \mathbf{A}\sigma_g^2 + \mathbf{I}\sigma_e^2$, where $\mathbf{A}$ is a genetic relatedness matrix (GRM) calculated with the *--make-grm* command in GCTA; the genomic relationship between individuals j and k is calculated as $A_{jk} = \frac{1}{N}\sum_i \frac{(x_{ij}-2p_i)(x_{ik}-2p_i)}{2p_i(1-p_i)}$, where *N* is the number of SNPs, $x_{ij}$ is the number of copies of the reference allele for the $i^{th}$ SNP and the $j^{th}$ or $k^{th}$ individual and $p_i$ is the frequency of the reference allele. We fitted the model for the denominator of the ratio estimator similarly, substituting maternal BMI for offspring phenotype ($\mathbf{y}$ in **Equation 1**). We calculated the GRM using imputed offspring SNPs with MAF >1%, imputation quality score >0.3 and (for ALSPAC only) HWE *P*-value >1e-6. The linear mixed model approach has been widely used in GWAS to control for population stratification and cryptic relatedness (48).

**Supplementary information S16: Descriptive statistics for the samples at baseline and the samples used for MV estimates**

| Cohort | Sample | Phenotype | *N* | Females (%) | BW (SD) [g] | Maternal BMI (SD) [$kg/m^2$] |
|---|---|---|---|---|---|---|
| ALSPAC | Live-born singletons at baseline | | 11134 | 48.80 | 3425 (537) | 22.9 (3.8) |
| | Sample for MV model 3, for the indicated phenotype | BW | 3265 | 51.03 | 3456 (510) | 22.9 (3.7) |
| | | 1yr BMI | 3145 | 51.16 | 3456 (509) | 22.9 (3.7) |
| | | 4yr BMI | 3060 | 50.78 | 3457 (509) | 22.9 (3.7) |
| | | 10yr BMI | 3007 | 51.31 | 3456 (505) | 22.9 (3.7) |
| | | 15yr BMI | 2795 | 51.66 | 3456 (507) | 22.9 (3.7) |
| | | 10yr FMI | 2627 | 51.43 | 3448 (509) | 22.9 (3.7) |
| | | 12yr FMI | 2598 | 51.58 | 3460 (508) | 22.9 (3.6) |
| | | 14yr FMI | 2424 | 51.82 | 3458 (508) | 22.8 (3.6) |
| | | 16yr FMI | 2105 | 52.64 | 3452 (503) | 22.8 (3.6) |
| | | 18yr FMI | 1884 | 55.04 | 3451 (507) | 22.7 (3.6) |
| BiB (South Asians) | Live-born singletons at baseline | | 5281 | 48.80 | 3082 (520) | 25.5 (5.4) |
| | Sample for MV model 3, for the indicated phenotype | BW | 449 | 46.55 | 3084 (465) | 25.1 (5.0) |
| | | 1yr BMI | 401 | 45.89 | 3084 (456) | 25.1 (5.1) |
| | | 4yr BMI | 325 | 47.38 | 3057 (463) | 25.3 (5.0) |
| BiB (White Europeans) | Live-born singletons at baseline | | 4338 | 48.09 | 3321 (552) | 26.6 (6.0) |
| | Sample for MV model 3, for the indicated phenotype | BW | 604 | 48.84 | 3379 (487) | 26.8 (6.0) |
| | | 1yr BMI | 559 | 49.55 | 3383 (486) | 26.9 (6.0) |
| | | 4yr BMI | 442 | 50.23 | 3381 (497) | 27.0 (6.0) |

**SD**: standard deviations

### Supplementary information S17: Confounder adjusted MV estimates for the association between maternal BMI and offspring outcomes, *retaining individuals with missing paternal BMI data*

| Cohort | Outcome | *N* | Model 1 | | | Model 2 | | | $P_{sex\ int.}$ |
|---|---|---|---|---|---|---|---|---|---|
| | | | β | 95% CI | *P* | β | 95% CI | *P* | |
| ALSPAC | BW | 4249 | 0.12 | 0.09, 0.15 | 7e-16 | 0.12 | 0.09, 0.15 | 2.1e-15 | 0.83 |
| | 1yr BMI | 4064 | 0.07 | 0.04, 0.10 | 3.7e-06 | 0.07 | 0.04, 0.11 | 3.7e-06 | 0.57 |
| | 4yr BMI | 3950 | 0.19 | 0.16, 0.22 | 3.6e-32 | 0.19 | 0.16, 0.23 | 7.5e-34 | 0.10 |
| | 10yr BMI | 3845 | 0.32 | 0.29, 0.35 | 3.7e-96 | 0.33 | 0.30, 0.36 | 2.3e-96 | 0.73 |
| | 15yr BMI | 3545 | 0.35 | 0.32, 0.38 | 7.6e-106 | 0.35 | 0.32, 0.38 | 6e-104 | 0.02 |
| | 10yr FMI | 3340 | 0.31 | 0.28, 0.34 | 2.7e-74 | 0.31 | 0.28, 0.34 | 1.9e-73 | 0.63 |
| | 12yr FMI | 3299 | 0.33 | 0.29, 0.36 | 4.8e-82 | 0.32 | 0.29, 0.36 | 1.6e-79 | 0.44 |
| | 14yr FMI | 3061 | 0.34 | 0.31, 0.38 | 1.1e-81 | 0.34 | 0.30, 0.37 | 1.6e-78 | 0.01 |
| | 16yr FMI | 2632 | 0.34 | 0.31, 0.38 | 2.6e-75 | 0.34 | 0.30, 0.38 | 1.1e-71 | 0.24 |
| | 18yr FMI | 2356 | 0.34 | 0.30, 0.38 | 8e-64 | 0.33 | 0.29, 0.37 | 6.2e-60 | 0.31 |
| BiB (SA) | BW | 2088 | 0.16 | 0.12, 0.20 | 4.9e-13 | 0.15 | 0.10, 0.19 | 5.9e-11 | 0.40 |
| | 1yr BMI | 1864 | 0.12 | 0.07, 0.16 | 3.9e-07 | 0.12 | 0.07, 0.16 | 1e-06 | 0.90 |
| | 4yr BMI | 1463 | 0.24 | 0.19, 0.29 | 1.7e-20 | 0.24 | 0.19, 0.29 | 1.3e-19 | 0.92 |
| BiB (WE) | BW | 1791 | 0.17 | 0.13, 0.22 | 1.5e-13 | 0.17 | 0.13, 0.22 | 5.5e-14 | 0.75 |
| | 1yr BMI | 1632 | 0.14 | 0.09, 0.19 | 1.4e-08 | 0.14 | 0.09, 0.19 | 2e-08 | 0.28 |
| | 4yr BMI | 1250 | 0.21 | 0.16, 0.27 | 1e-15 | 0.22 | 0.17, 0.28 | 3.6e-16 | 0.68 |

**SA**: South Asians, **WE**: White Europeans, **Model 1**: controlled for maternal age, offspring age and sex in the standardised exposure and outcome, **Model 2**: additionally adjusted for potential confounders including parity, maternal smoking during pregnancy, maternal and paternal education and parental occupation, **Model 3**: additionally adjusted for paternal BMI, $P_{sex\ int.}$: *P*-value for exposure * sex interaction (covariates as per model 2), **β**: coefficient from linear regression of outcome (age- [except for BW] and sex-standardised *z*-score) on maternal BMI (age-standardised *z*-score)

### Supplementary information S18: Characteristics of the mothers and offspring in ALSPAC and BiB (absolute values)

| | ALSPAC | | | | BiB (WE) | | | | BiB (SA) | | | |
|---|---|---|---|---|---|---|---|---|---|---|---|---|
| | Mean | SD | *N* | Female offspring (%) | Mean | SD | *N* | Female offspring (%) | Mean | SD | *N* | Female offspring (%) |
| Birth weight (kg) | 3.45 | 0.52 | 5085 | 50.5 | 3.36 | 0.53 | 1992 | 47.9 | 3.12 | 0.49 | 2262 | 47.9 |
| 1yr BMI (kg/m$^2$) | 17.5 | 1.5 | 4838 | 50.6 | 17.4 | 1.6 | 1798 | 47.8 | 16.7 | 1.6 | 2023 | 48.1 |
| 4yr BMI (kg/m$^2$) | 16.1 | 1.5 | 4670 | 50.2 | 16.4 | 1.5 | 1339 | 48.6 | 15.9 | 1.8 | 1566 | 48.5 |
| 10yr BMI (kg/m$^2$) | 17.7 | 2.8 | 4476 | 51.3 | | | | | | | | |
| 15yr BMI (kg/m$^2$) | 21.0 | 3.5 | 4112 | 51.7 | | | | | | | | |

**SA**: South Asians, **WE**: White Europeans, **SD**: standard deviation

### Supplementary information S19: IV *F*-statistics

| Outcome | IV | ALSPAC | | BiB (South Asians) | | BiB (White Europeans) | |
|---|---|---|---|---|---|---|---|
| | | ***F*-statistic** | ***N*** | ***F*-statistic** | ***N*** | ***F*-statistic** | ***N*** |
| BW | FTO | 19.2 | 5085 | 18.0 | 2262 | 11.4 | 1992 |
| | Speliotes | 43.8 | 5085 | 39.6 | 2262 | 18.7 | 1992 |
| | Locke | 53.5 | 5085 | 30.7 | 2262 | 23.8 | 1992 |
| | Yengo | 124.6 | 5085 | 37.9 | 2262 | 37.2 | 1992 |
| | Lassosum | 376.8 | 5085 | 81.5 | 2262 | 110.8 | 1992 |
| 1yr BMI | FTO | 15.5 | 4838 | 18.3 | 2023 | 10.0 | 1798 |
| | Speliotes | 38.7 | 4838 | 31.5 | 2023 | 15.3 | 1798 |
| | Locke | 44.9 | 4838 | 25.8 | 2023 | 18.2 | 1798 |
| | Yengo | 116.6 | 4838 | 37.8 | 2023 | 32.2 | 1798 |
| | Lassosum | 350.6 | 4838 | 80.6 | 2023 | 95.5 | 1798 |
| 4yr BMI | FTO | 18.2 | 4670 | 12.0 | 1566 | 15.1 | 1339 |
| | Speliotes | 44.6 | 4670 | 34.3 | 1566 | 19.8 | 1339 |
| | Locke | 54.4 | 4670 | 20.8 | 1566 | 17.9 | 1339 |
| | Yengo | 115.0 | 4670 | 25.1 | 1566 | 25.5 | 1339 |
| | Lassosum | 344.5 | 4670 | 78.2 | 1566 | 80.9 | 1339 |
| 10yr BMI | FTO | 20.5 | 4476 | | | | |
| | Speliotes | 49.1 | 4476 | | | | |
| | Locke | 54.3 | 4476 | | | | |
| | Yengo | 104.8 | 4476 | | | | |
| | Lassosum | 329.3 | 4476 | | | | |
| 15yr BMI | FTO | 16.5 | 4112 | | | | |
| | Speliotes | 37.6 | 4112 | | | | |
| | Locke | 41.8 | 4112 | | | | |
| | Yengo | 96.8 | 4112 | | | | |
| | Lassosum | 279.3 | 4112 | | | | |
| 10yr FMI | FTO | 16.8 | 3855 | | | | |
| | Speliotes | 37.7 | 3855 | | | | |
| | Locke | 45.6 | 3855 | | | | |
| | Yengo | 92.8 | 3855 | | | | |
| | Lassosum | 285.1 | 3855 | | | | |
| 12yr FMI | FTO | 13.3 | 3807 | | | | |
| | Speliotes | 35.3 | 3807 | | | | |
| | Locke | 40.3 | 3807 | | | | |
| | Yengo | 93.0 | 3807 | | | | |
| | Lassosum | 275.2 | 3807 | | | | |
| 14yr FMI | FTO | 13.4 | 3506 | | | | |
| | Speliotes | 34.9 | 3506 | | | | |
| | Locke | 37.1 | 3506 | | | | |
| | Yengo | 87.2 | 3506 | | | | |
| | Lassosum | 240.4 | 3506 | | | | |
| 16yr FMI | FTO | 10.0 | 2996 | | | | |
| | Speliotes | 29.9 | 2996 | | | | |
| | Locke | 36.1 | 2996 | | | | |
| | Yengo | 83.5 | 2996 | | | | |
| | Lassosum | 212.2 | 2996 | | | | |
| 18yr FMI | FTO | 16.6 | 2659 | | | | |
| | Speliotes | 33.4 | 2659 | | | | |
| | Locke | 39.5 | 2659 | | | | |
| | Yengo | 81.8 | 2659 | | | | |
| | Lassosum | 223.7 | 2659 | | | | |

**Supplementary information S20: Associations between instrumental variables, exposures and outcome risk factors, ALSPAC**

| Independent variable | | | Genetic IV for BMI (z-score; dependent variable) | | | | | | | | | | | | | | | | | |
|---|---|---|---|---|---|---|---|---|---|---|---|---|---|---|---|---|---|---|---|---|
| | | | **FTO** | | | **Speliotes** | | | **Locke** | | | **Yengo** | | | **Lassosum** | | | **Maternal BMI (age standardised *z*-score)** | | |
| | | ***N*** | **Beta** | **95% CI** | ***P*** | **Beta** | **95% CI** | ***P*** | **Beta** | **95% CI** | ***P*** | **Beta** | **95% CI** | ***P*** | **Beta** | **95% CI** | ***P*** | **Beta** | **95% CI** | ***P*** |
| **Parental occupation** | I | 745 | Ref | Ref | Ref | Ref | Ref | Ref | Ref | Ref | Ref | Ref | Ref | Ref | Ref | Ref | Ref | Ref | Ref | Ref |
| | II | 2137 | -0.02 | -0.10, 0.07 | 6.72e-01 | 0.03 | -0.05, 0.12 | 4.62e-01 | 0.02 | -0.07, 0.10 | 7.19e-01 | 0.07 | -0.02, 0.15 | 1.19e-01 | 0.19 | 0.11, 0.27 | 8.81e-06 | 0.20 | 0.12, 0.28 | 1.84e-06 |
| | III (non-manual) | 1207 | 0.02 | -0.07, 0.11 | 6.24e-01 | 0.01 | -0.09, 0.10 | 9.01e-01 | 0.01 | -0.08, 0.10 | 8.76e-01 | 0.04 | -0.05, 0.13 | 3.60e-01 | 0.19 | 0.09, 0.28 | 6.64e-05 | 0.32 | 0.23, 0.41 | 1.43e-12 |
| | III (manual) | 516 | 0.08 | -0.03, 0.20 | 1.44e-01 | 0.07 | -0.04, 0.19 | 1.97e-01 | 0.08 | -0.03, 0.19 | 1.50e-01 | 0.16 | 0.05, 0.27 | 5.41e-03 | 0.25 | 0.13, 0.36 | 1.87e-05 | 0.33 | 0.22, 0.44 | 6.01e-09 |
| | IV | 175 | 0.11 | -0.06, 0.27 | 1.98e-01 | 0.10 | -0.07, 0.26 | 2.42e-01 | 0.11 | -0.06, 0.27 | 2.11e-01 | 0.20 | 0.03, 0.36 | 1.96e-02 | 0.41 | 0.25, 0.58 | 9.22e-07 | 0.43 | 0.27, 0.59 | 2.04e-07 |
| | V | 27 | 0.04 | -0.34, 0.43 | 8.28e-01 | 0.17 | -0.22, 0.55 | 3.95e-01 | 0.25 | -0.13, 0.64 | 1.95e-01 | 0.30 | -0.08, 0.69 | 1.25e-01 | 0.73 | 0.35, 1.11 | 1.98e-04 | 0.51 | 0.14, 0.89 | 7.73e-03 |
| **Maternal education** | Degree | 795 | -0.01 | -0.12, 0.10 | 8.46e-01 | -0.02 | -0.13, 0.09 | 7.16e-01 | -0.05 | -0.16, 0.06 | 3.86e-01 | -0.17 | -0.28, -0.05 | 3.69e-03 | -0.28 | -0.39, -0.17 | 1.19e-06 | -0.42 | -0.53, -0.32 | 3.47e-14 |
| | A-level | 1316 | -0.05 | -0.15, 0.05 | 3.31e-01 | 0.06 | -0.04, 0.16 | 2.52e-01 | 0.04 | -0.06, 0.15 | 4.29e-01 | -0.07 | -0.17, 0.04 | 1.95e-01 | -0.16 | -0.27, -0.06 | 2.04e-03 | -0.24 | -0.34, -0.14 | 4.55e-06 |
| | O-level | 1793 | -0.03 | -0.13, 0.07 | 5.68e-01 | 0.06 | -0.04, 0.16 | 2.19e-01 | 0.05 | -0.05, 0.15 | 3.08e-01 | -0.02 | -0.12, 0.08 | 6.76e-01 | -0.06 | -0.16, 0.04 | 2.26e-01 | -0.14 | -0.24, -0.04 | 4.75e-03 |
| | Vocational | 421 | -0.05 | -0.18, 0.08 | 4.86e-01 | -0.03 | -0.16, 0.11 | 7.07e-01 | -0.04 | -0.17, 0.09 | 5.31e-01 | -0.16 | -0.29, -0.03 | 1.71e-02 | -0.15 | -0.28, -0.02 | 2.66e-02 | -0.06 | -0.19, 0.07 | 3.52e-01 |
| | CSE/none | 501 | Ref | Ref | Ref | Ref | Ref | Ref | Ref | Ref | Ref | Ref | Ref | Ref | Ref | Ref | Ref | Ref | Ref | Ref |
| **Paternal education** | Degree | 1040 | 0.02 | -0.08, 0.12 | 6.95e-01 | -0.08 | -0.18, 0.02 | 1.11e-01 | -0.10 | -0.19, -0.00 | 4.97e-02 | -0.13 | -0.23, -0.03 | 8.43e-03 | -0.28 | -0.38, -0.18 | 1.73e-08 | -0.37 | -0.46, -0.28 | 1.45e-14 |
| | A-level | 1359 | -0.04 | -0.13, 0.05 | 3.78e-01 | -0.05 | -0.14, 0.04 | 2.92e-01 | -0.05 | -0.15, 0.04 | 2.46e-01 | -0.08 | -0.17, 0.01 | 8.44e-02 | -0.14 | -0.23, -0.05 | 3.32e-03 | -0.13 | -0.22, -0.05 | 3.18e-03 |
| | O-level | 1092 | 0.08 | -0.01, 0.18 | 9.00e-02 | 0.02 | -0.08, 0.11 | 7.35e-01 | 0.01 | -0.08, 0.11 | 7.89e-01 | -0.01 | -0.11, 0.09 | 8.50e-01 | -0.11 | -0.21, -0.02 | 2.24e-02 | -0.13 | -0.22, -0.03 | 8.43e-03 |
| | Vocational | 396 | 0.02 | -0.10, 0.15 | 7.03e-01 | -0.00 | -0.13, 0.12 | 9.64e-01 | 0.00 | -0.12, 0.13 | 9.85e-01 | 0.05 | -0.08, 0.17 | 4.39e-01 | -0.14 | -0.26, -0.01 | 2.93e-02 | -0.02 | -0.14, 0.10 | 7.31e-01 |
| | CSE/none | 685 | Ref | Ref | Ref | Ref | Ref | Ref | Ref | Ref | Ref | Ref | Ref | Ref | Ref | Ref | Ref | Ref | Ref | Ref |
| **Maternal smoking in pregnancy** | Never | 3554 | Ref | Ref | Ref | Ref | Ref | Ref | Ref | Ref | Ref | Ref | Ref | Ref | Ref | Ref | Ref | Ref | Ref | Ref |
| | Early pregnancy | 494 | -0.02 | -0.11, 0.07 | 6.76e-01 | 0.05 | -0.05, 0.14 | 3.38e-01 | 0.05 | -0.05, 0.14 | 3.26e-01 | 0.09 | -0.00, 0.18 | 6.11e-02 | 0.19 | 0.09, 0.28 | 9.55e-05 | 0.07 | -0.03, 0.16 | 1.64e-01 |
| | Throughout pregnancy | 843 | 0.01 | -0.07, 0.08 | 8.69e-01 | -0.04 | -0.11, 0.04 | 3.60e-01 | -0.04 | -0.12, 0.03 | 2.77e-01 | 0.03 | -0.05, 0.10 | 5.12e-01 | 0.15 | 0.07, 0.22 | 1.06e-04 | -0.04 | -0.11, 0.04 | 3.26e-01 |
| **Parity** | | 5042 | 0.02 | -0.01, 0.05 | 1.19e-01 | 0.03 | -0.00, 0.06 | 9.73e-02 | 0.03 | 0.00, 0.06 | 3.23e-02 | 0.03 | -0.00, 0.06 | 5.69e-02 | 0.03 | -0.00, 0.06 | 9.88e-02 | 0.06 | 0.03, 0.09 | 4.30e-05 |
| **Maternal age (years)** | | 5157 | -0.00 | -0.01, 0.00 | 7.14e-01 | 0.00 | -0.01, 0.01 | 9.78e-01 | -0.00 | -0.01, 0.01 | 9.62e-01 | -0.00 | -0.01, 0.00 | 1.63e-01 | -0.02 | -0.02, -0.01 | 4.65e-09 | -0.00 | -0.01, 0.01 | 8.00e-01 |
| **Paternal age (years)** | | 3593 | 0.00 | -0.01, 0.01 | 9.34e-01 | 0.00 | -0.01, 0.01 | 7.56e-01 | -0.00 | -0.01, 0.01 | 9.65e-01 | -0.00 | -0.01, 0.00 | 6.42e-01 | -0.01 | -0.02, -0.01 | 2.26e-04 | -0.00 | -0.01, 0.00 | 5.39e-01 |
| **Paternal BMI (kg/m$^2$)** | | 3766 | -0.01 | -0.04, 0.02 | 4.88e-01 | 0.01 | -0.02, 0.05 | 3.92e-01 | 0.01 | -0.02, 0.04 | 5.71e-01 | 0.01 | -0.02, 0.04 | 5.75e-01 | 0.03 | 0.00, 0.06 | 4.85e-02 | 0.05 | 0.04, 0.06 | 3.59e-23 |

Abbreviations are as for **Figure 1**

### Supplementary information S21: Associations between instrumental variables, exposures and outcome risk factors, BiB South Asians

| Independent variable | | | Genetic IV for BMI (z-score; dependent variable) | | | | | | | | | | | | | | | | | |
|---|---|---|---|---|---|---|---|---|---|---|---|---|---|---|---|---|---|---|---|---|
| | | | FTO | | | Speliotes | | | Locke | | | Yengo | | | Lassosum | | | Maternal BMI (age standardised *z*-score) | | |
| | | ***N*** | **Beta** | **95% CI** | ***P*** | **Beta** | **95% CI** | ***P*** | **Beta** | **95% CI** | ***P*** | **Beta** | **95% CI** | ***P*** | **Beta** | **95% CI** | ***P*** | **Beta** | **95% CI** | ***P*** |
| **Parental occupation** | Modern professional | 140 | Ref | Ref | Ref | Ref | Ref | Ref | Ref | Ref | Ref | Ref | Ref | Ref | Ref | Ref | Ref | Ref | Ref | Ref |
| | Clerical and intermediate | 152 | -0.18 | -0.41, 0.05 | 1.31e-01 | -0.07 | -0.30, 0.15 | 5.20e-01 | -0.10 | -0.33, 0.13 | 4.07e-01 | -0.14 | -0.37, 0.09 | 2.32e-01 | -0.05 | -0.29, 0.18 | 6.50e-01 | -0.18 | -0.41, 0.05 | 1.18e-01 |
| | Sr. managers/administrators | 75 | -0.36 | -0.64, -0.08 | 1.27e-02 | -0.11 | -0.39, 0.16 | 4.22e-01 | -0.11 | -0.39, 0.17 | 4.28e-01 | 0.01 | -0.27, 0.29 | 9.29e-01 | -0.10 | -0.39, 0.18 | 4.80e-01 | -0.17 | -0.45, 0.11 | 2.27e-01 |
| | Technical and craft | 113 | -0.09 | -0.34, 0.16 | 4.88e-01 | -0.10 | -0.35, 0.15 | 4.29e-01 | -0.03 | -0.27, 0.22 | 8.32e-01 | -0.01 | -0.26, 0.24 | 9.42e-01 | -0.10 | -0.35, 0.15 | 4.44e-01 | 0.09 | -0.16, 0.34 | 4.71e-01 |
| | Semi-routine manual/service | 408 | -0.15 | -0.34, 0.04 | 1.23e-01 | -0.09 | -0.29, 0.10 | 3.32e-01 | -0.01 | -0.20, 0.18 | 9.25e-01 | 0.07 | -0.12, 0.26 | 4.54e-01 | 0.00 | -0.19, 0.20 | 9.66e-01 | 0.11 | -0.08, 0.30 | 2.74e-01 |
| | Routine manual and service | 460 | -0.16 | -0.36, 0.03 | 8.98e-02 | -0.01 | -0.20, 0.18 | 8.96e-01 | 0.02 | -0.17, 0.20 | 8.75e-01 | -0.00 | -0.19, 0.19 | 9.92e-01 | -0.02 | -0.21, 0.18 | 8.65e-01 | 0.12 | -0.07, 0.30 | 2.21e-01 |
| | Middle or junior managers | 122 | -0.19 | -0.43, 0.05 | 1.29e-01 | 0.09 | -0.16, 0.33 | 4.87e-01 | 0.02 | -0.22, 0.26 | 8.79e-01 | -0.10 | -0.34, 0.14 | 4.16e-01 | 0.07 | -0.17, 0.32 | 5.52e-01 | -0.15 | -0.39, 0.09 | 2.26e-01 |
| | Traditional professional | 101 | -0.26 | -0.52, -0.00 | 4.62e-02 | -0.03 | -0.28, 0.23 | 8.37e-01 | -0.07 | -0.33, 0.18 | 5.88e-01 | -0.03 | -0.28, 0.23 | 8.23e-01 | -0.13 | -0.39, 0.13 | 3.20e-01 | -0.30 | -0.55, -0.05 | 2.10e-02 |
| | Self-employed | 323 | -0.25 | -0.45, -0.05 | 1.46e-02 | 0.04 | -0.16, 0.24 | 6.91e-01 | 0.06 | -0.14, 0.26 | 5.57e-01 | 0.02 | -0.18, 0.22 | 8.27e-01 | -0.08 | -0.28, 0.12 | 4.47e-01 | -0.01 | -0.21, 0.18 | 8.93e-01 |
| | Student/in training | 128 | -0.11 | -0.35, 0.13 | 3.61e-01 | -0.04 | -0.28, 0.20 | 7.56e-01 | 0.03 | -0.21, 0.27 | 8.36e-01 | -0.18 | -0.42, 0.06 | 1.43e-01 | -0.07 | -0.32, 0.17 | 5.47e-01 | -0.02 | -0.26, 0.22 | 8.64e-01 |
| | Unemployed/sick leave | 118 | -0.14 | -0.39, 0.11 | 2.65e-01 | 0.05 | -0.20, 0.29 | 7.08e-01 | 0.14 | -0.10, 0.39 | 2.58e-01 | 0.19 | -0.06, 0.43 | 1.30e-01 | 0.01 | -0.24, 0.26 | 9.37e-01 | 0.03 | -0.21, 0.27 | 7.95e-01 |
| | Unknown | 24 | -0.39 | -0.83, 0.04 | 7.70e-02 | -0.20 | -0.63, 0.24 | 3.74e-01 | -0.16 | -0.59, 0.27 | 4.75e-01 | 0.10 | -0.33, 0.53 | 6.61e-01 | 0.01 | -0.43, 0.45 | 9.64e-01 | -0.07 | -0.50, 0.36 | 7.38e-01 |
| **Maternal education** | Higher than A-level | 626 | -0.03 | -0.15, 0.09 | 6.37e-01 | 0.05 | -0.07, 0.17 | 4.20e-01 | 0.04 | -0.07, 0.16 | 4.65e-01 | -0.14 | -0.26, -0.03 | 1.53e-02 | -0.09 | -0.21, 0.03 | 1.35e-01 | -0.30 | -0.41, -0.19 | 2.20e-07 |
| | A-level equivalent | 305 | -0.06 | -0.20, 0.08 | 3.90e-01 | -0.02 | -0.16, 0.12 | 7.63e-01 | -0.06 | -0.20, 0.08 | 4.08e-01 | -0.14 | -0.28, 0.00 | 5.50e-02 | 0.02 | -0.12, 0.16 | 7.96e-01 | -0.09 | -0.23, 0.05 | 1.92e-01 |
| | 5 GCSE equivalent | 665 | -0.02 | -0.13, 0.10 | 7.55e-01 | 0.03 | -0.08, 0.14 | 5.82e-01 | 0.00 | -0.11, 0.11 | 9.81e-01 | -0.03 | -0.14, 0.08 | 6.06e-01 | -0.05 | -0.17, 0.06 | 3.48e-01 | -0.03 | -0.15, 0.08 | 5.42e-01 |
| | <5 GCSE equivalent | 562 | Ref | Ref | Ref | Ref | Ref | Ref | Ref | Ref | Ref | Ref | Ref | Ref | Ref | Ref | Ref | Ref | Ref | Ref |
| **Paternal education** | Higher than A-level) | 671 | 0.05 | -0.08, 0.19 | 4.29e-01 | 0.07 | -0.07, 0.20 | 3.26e-01 | 0.09 | -0.05, 0.22 | 1.93e-01 | -0.02 | -0.15, 0.11 | 7.69e-01 | -0.02 | -0.15, 0.12 | 8.11e-01 | -0.24 | -0.37, -0.11 | 3.16e-04 |
| | A-level equivalent | 214 | 0.04 | -0.13, 0.22 | 6.19e-01 | 0.03 | -0.14, 0.21 | 7.21e-01 | 0.04 | -0.13, 0.22 | 6.39e-01 | -0.05 | -0.22, 0.12 | 5.79e-01 | 0.06 | -0.11, 0.24 | 4.96e-01 | -0.23 | -0.40, -0.06 | 8.19e-03 |
| | 5 GCSE equivalent | 542 | 0.06 | -0.08, 0.20 | 3.72e-01 | 0.00 | -0.14, 0.14 | 9.73e-01 | 0.05 | -0.09, 0.19 | 4.71e-01 | -0.03 | -0.17, 0.11 | 6.72e-01 | 0.08 | -0.06, 0.23 | 2.35e-01 | 0.01 | -0.13, 0.14 | 9.28e-01 |
| | <5 GCSE equivalent | 331 | Ref | Ref | Ref | Ref | Ref | Ref | Ref | Ref | Ref | Ref | Ref | Ref | Ref | Ref | Ref | Ref | Ref | Ref |
| **Maternal smoking in pregnancy** | No | 2178 | Ref | Ref | Ref | Ref | Ref | Ref | Ref | Ref | Ref | Ref | Ref | Ref | Ref | Ref | Ref | Ref | Ref | Ref |
| | Yes | 84 | -0.15 | -0.37, 0.07 | 1.74e-01 | -0.18 | -0.39, 0.04 | 1.10e-01 | -0.23 | -0.44, -0.01 | 4.09e-02 | 0.04 | -0.17, 0.26 | 6.88e-01 | 0.02 | -0.20, 0.24 | 8.62e-01 | 0.05 | -0.16, 0.27 | 6.27e-01 |
| **Parity** | | 2215 | -0.00 | -0.03, 0.03 | 9.07e-01 | 0.00 | -0.03, 0.03 | 9.23e-01 | -0.00 | -0.03, 0.03 | 8.77e-01 | 0.01 | -0.02, 0.04 | 5.08e-01 | -0.01 | -0.05, 0.02 | 3.45e-01 | 0.11 | 0.08, 0.14 | 4.08e-13 |
| **Maternal age (years)** | | 2267 | 0.00 | -0.01, 0.01 | 6.92e-01 | -0.00 | -0.01, 0.01 | 8.77e-01 | -0.00 | -0.01, 0.01 | 8.88e-01 | -0.00 | -0.01, 0.01 | 5.33e-01 | -0.01 | -0.02, 0.00 | 1.01e-01 | -0.00 | -0.01, 0.00 | 4.16e-01 |
| **Paternal age (years)** | | 583 | 0.01 | -0.00, 0.02 | 1.80e-01 | -0.01 | -0.02, 0.00 | 2.32e-01 | 0.00 | -0.01, 0.02 | 5.56e-01 | -0.00 | -0.02, 0.01 | 6.13e-01 | -0.01 | -0.02, 0.01 | 2.60e-01 | 0.00 | -0.01, 0.02 | 6.35e-01 |
| **Paternal BMI ($kg/m^2$)** | | 475 | 0.02 | -0.07, 0.11 | 6.42e-01 | 0.01 | -0.08, 0.09 | 8.93e-01 | -0.05 | -0.14, 0.04 | 2.95e-01 | -0.00 | -0.10, 0.09 | 9.60e-01 | -0.02 | -0.12, 0.07 | 6.22e-01 | 0.02 | 0.01, 0.04 | 9.07e-03 |

Abbreviations are as for **Figure 1**

### Supplementary information S22: Associations between instrumental variables, exposures and outcome risk factors, BiB White Europeans

| Independent variable | | | Genetic IV for BMI (z-score; dependent variable) | | | | | | | | | | | | | | | | | |
|---|---|---|---|---|---|---|---|---|---|---|---|---|---|---|---|---|---|---|---|---|
| | | | FTO | | | Speliotes | | | Locke | | | Yengo | | | Lassosum | | | Maternal BMI (age standardised *z*-score) | | |
| | | ***N*** | **Beta** | **95% CI** | ***P*** | **Beta** | **95% CI** | ***P*** | **Beta** | **95% CI** | ***P*** | **Beta** | **95% CI** | ***P*** | **Beta** | **95% CI** | ***P*** | **Beta** | **95% CI** | ***P*** |
| **Parental occupation** | Modern professional | 164 | Ref | Ref | Ref | Ref | Ref | Ref | Ref | Ref | Ref | Ref | Ref | Ref | Ref | Ref | Ref | Ref | Ref | Ref |
| | Clerical and intermediate | 124 | -0.07 | -0.31, 0.16 | 5.55e-01 | -0.01 | -0.25, 0.22 | 9.07e-01 | -0.07 | -0.30, 0.17 | 5.85e-01 | -0.03 | -0.27, 0.20 | 7.84e-01 | 0.06 | -0.17, 0.30 | 5.98e-01 | 0.38 | 0.15, 0.61 | 1.34e-03 |
| | Sr. managers/administrators | 81 | -0.08 | -0.34, 0.19 | 5.69e-01 | -0.04 | -0.30, 0.22 | 7.65e-01 | 0.01 | -0.26, 0.28 | 9.41e-01 | -0.11 | -0.38, 0.15 | 4.07e-01 | -0.11 | -0.37, 0.16 | 4.34e-01 | 0.11 | -0.16, 0.37 | 4.29e-01 |
| | Technical and craft | 342 | -0.16 | -0.34, 0.03 | 1.01e-01 | -0.02 | -0.21, 0.16 | 8.09e-01 | -0.07 | -0.25, 0.12 | 4.75e-01 | -0.01 | -0.20, 0.17 | 8.96e-01 | 0.07 | -0.11, 0.26 | 4.30e-01 | 0.18 | -0.00, 0.36 | 5.47e-02 |
| | Semi-routine manual/service | 233 | -0.18 | -0.38, 0.02 | 7.74e-02 | -0.04 | -0.24, 0.16 | 6.68e-01 | -0.07 | -0.27, 0.13 | 5.15e-01 | 0.05 | -0.15, 0.25 | 6.32e-01 | 0.05 | -0.15, 0.25 | 6.27e-01 | 0.33 | 0.13, 0.53 | 1.09e-03 |
| | Routine manual and service | 314 | -0.06 | -0.25, 0.13 | 5.66e-01 | -0.13 | -0.32, 0.06 | 1.73e-01 | -0.09 | -0.28, 0.09 | 3.28e-01 | -0.07 | -0.26, 0.12 | 4.87e-01 | 0.15 | -0.04, 0.34 | 1.25e-01 | 0.24 | 0.06, 0.43 | 1.03e-02 |
| | Middle or junior managers | 139 | -0.13 | -0.36, 0.09 | 2.45e-01 | -0.10 | -0.33, 0.12 | 3.70e-01 | -0.13 | -0.35, 0.10 | 2.68e-01 | -0.01 | -0.24, 0.21 | 9.12e-01 | -0.05 | -0.27, 0.18 | 6.74e-01 | 0.10 | -0.12, 0.32 | 3.70e-01 |
| | Traditional professional | 61 | -0.18 | -0.48, 0.11 | 2.29e-01 | 0.06 | -0.23, 0.36 | 6.73e-01 | 0.13 | -0.16, 0.42 | 3.81e-01 | -0.25 | -0.55, 0.04 | 9.33e-02 | -0.13 | -0.42, 0.16 | 3.79e-01 | -0.02 | -0.31, 0.27 | 9.11e-01 |
| | Self-employed | 154 | -0.12 | -0.34, 0.10 | 2.87e-01 | -0.04 | -0.26, 0.18 | 7.01e-01 | -0.16 | -0.38, 0.05 | 1.41e-01 | -0.03 | -0.25, 0.19 | 7.72e-01 | 0.05 | -0.17, 0.27 | 6.55e-01 | 0.05 | -0.17, 0.26 | 6.76e-01 |
| | Student/in training | 76 | -0.08 | -0.35, 0.19 | 5.59e-01 | -0.04 | -0.31, 0.23 | 7.91e-01 | 0.08 | -0.19, 0.35 | 5.79e-01 | -0.08 | -0.35, 0.20 | 5.79e-01 | 0.14 | -0.13, 0.41 | 3.07e-01 | 0.07 | -0.19, 0.34 | 5.91e-01 |
| | Unemployed/sick leave | 129 | -0.29 | -0.52, -0.05 | 1.61e-02 | -0.20 | -0.43, 0.03 | 8.92e-02 | -0.10 | -0.33, 0.13 | 3.89e-01 | -0.05 | -0.28, 0.18 | 6.56e-01 | 0.18 | -0.05, 0.41 | 1.33e-01 | 0.29 | 0.06, 0.52 | 1.31e-02 |
| | Unknown | 29 | 0.18 | -0.23, 0.59 | 3.87e-01 | 0.02 | -0.38, 0.43 | 9.07e-01 | 0.02 | -0.39, 0.43 | 9.18e-01 | 0.04 | -0.37, 0.45 | 8.34e-01 | 0.05 | -0.36, 0.45 | 8.21e-01 | 0.45 | 0.06, 0.84 | 2.30e-02 |
| **Maternal education** | Higher than A-level | 396 | 0.05 | -0.10, 0.19 | 5.25e-01 | 0.10 | -0.04, 0.24 | 1.66e-01 | 0.04 | -0.10, 0.18 | 5.97e-01 | -0.00 | -0.15, 0.14 | 9.48e-01 | -0.14 | -0.28, 0.00 | 5.05e-02 | -0.17 | -0.31, -0.03 | 1.56e-02 |
| | A-level equivalent | 369 | 0.02 | -0.12, 0.17 | 7.70e-01 | -0.01 | -0.15, 0.13 | 8.64e-01 | -0.02 | -0.17, 0.12 | 7.48e-01 | -0.02 | -0.16, 0.13 | 8.19e-01 | -0.19 | -0.34, -0.05 | 8.39e-03 | 0.05 | -0.09, 0.19 | 4.82e-01 |
| | 5 GCSE equivalent | 660 | -0.02 | -0.15, 0.11 | 7.38e-01 | 0.04 | -0.08, 0.17 | 5.19e-01 | 0.03 | -0.09, 0.16 | 6.09e-01 | 0.06 | -0.07, 0.19 | 3.49e-01 | -0.02 | -0.14, 0.11 | 7.81e-01 | 0.12 | -0.01, 0.24 | 6.58e-02 |
| | <5 GCSE equivalent | 381 | Ref | Ref | Ref | Ref | Ref | Ref | Ref | Ref | Ref | Ref | Ref | Ref | Ref | Ref | Ref | Ref | Ref | Ref |
| **Paternal education** | Higher than A-level) | 309 | 0.04 | -0.12, 0.20 | 6.20e-01 | -0.00 | -0.16, 0.15 | 9.66e-01 | 0.00 | -0.15, 0.16 | 9.71e-01 | 0.04 | -0.12, 0.19 | 6.18e-01 | -0.16 | -0.31, 0.00 | 5.04e-02 | -0.09 | -0.25, 0.06 | 2.31e-01 |
| | A-level equivalent | 241 | 0.10 | -0.07, 0.27 | 2.47e-01 | 0.10 | -0.07, 0.26 | 2.51e-01 | -0.03 | -0.19, 0.14 | 7.29e-01 | 0.06 | -0.11, 0.22 | 5.11e-01 | -0.18 | -0.35, -0.02 | 2.95e-02 | -0.05 | -0.21, 0.11 | 5.58e-01 |
| | 5 GCSE equivalent | 518 | -0.05 | -0.19, 0.08 | 4.43e-01 | -0.13 | -0.27, 0.01 | 6.04e-02 | -0.13 | -0.26, 0.01 | 7.26e-02 | -0.01 | -0.15, 0.13 | 9.02e-01 | -0.15 | -0.29, -0.01 | 3.29e-02 | 0.05 | -0.09, 0.18 | 5.08e-01 |
| | <5 GCSE equivalent | 333 | Ref | Ref | Ref | Ref | Ref | Ref | Ref | Ref | Ref | Ref | Ref | Ref | Ref | Ref | Ref | Ref | Ref | Ref |
| **Maternal smoking in pregnancy** | No | 1324 | Ref | Ref | Ref | Ref | Ref | Ref | Ref | Ref | Ref | Ref | Ref | Ref | Ref | Ref | Ref | Ref | Ref | Ref |
| | Yes | 675 | 0.00 | -0.09, 0.09 | 9.96e-01 | -0.06 | -0.16, 0.03 | 1.68e-01 | -0.01 | -0.11, 0.08 | 7.92e-01 | 0.02 | -0.07, 0.11 | 6.74e-01 | 0.15 | 0.06, 0.24 | 1.33e-03 | 0.02 | -0.07, 0.12 | 6.03e-01 |
| **Parity** | | 1951 | -0.00 | -0.04, 0.04 | 9.74e-01 | 0.00 | -0.04, 0.05 | 8.83e-01 | -0.00 | -0.05, 0.04 | 9.14e-01 | 0.01 | -0.03, 0.05 | 7.01e-01 | 0.04 | -0.00, 0.09 | 5.26e-02 | 0.11 | 0.06, 0.15 | 1.11e-06 |
| **Maternal age (years)** | | 2000 | 0.00 | -0.01, 0.01 | 7.57e-01 | 0.00 | -0.01, 0.01 | 8.52e-01 | -0.00 | -0.01, 0.00 | 3.41e-01 | -0.00 | -0.01, 0.00 | 2.33e-01 | -0.01 | -0.02, -0.00 | 5.81e-03 | 0.00 | -0.01, 0.01 | 9.80e-01 |
| **Paternal age (years)** | | 788 | -0.00 | -0.01, 0.01 | 5.41e-01 | -0.00 | -0.01, 0.01 | 5.48e-01 | -0.00 | -0.01, 0.01 | 5.52e-01 | -0.00 | -0.01, 0.01 | 9.47e-01 | 0.00 | -0.01, 0.01 | 7.29e-01 | 0.01 | -0.00, 0.02 | 3.08e-01 |
| **Paternal BMI ($kg/m^2$)** | | 639 | 0.04 | -0.04, 0.11 | 3.60e-01 | 0.02 | -0.06, 0.09 | 6.68e-01 | 0.04 | -0.04, 0.12 | 3.22e-01 | 0.01 | -0.07, 0.09 | 8.13e-01 | 0.04 | -0.04, 0.12 | 3.29e-01 | 0.05 | 0.03, 0.06 | 1.11e-08 |

Abbreviations are as for **Figure 1**

### Supplementary information S23: Associations between instrumental variables, exposures and outcome risk factors, BiB South Asians and White Europeans

| Independent variable | | | Genetic IV for BMI (z-score; dependent variable) | | | | | | | | | | | | | | | | | |
|---|---|---|---|---|---|---|---|---|---|---|---|---|---|---|---|---|---|---|---|---|
| | | | FTO | | | Speliotes | | | Locke | | | Yengo | | | Lassosum | | | Maternal BMI (age standardised *z*-score) | | |
| | | ***N*** | **Beta** | **95% CI** | ***P*** | **Beta** | **95% CI** | ***P*** | **Beta** | **95% CI** | ***P*** | **Beta** | **95% CI** | ***P*** | **Beta** | **95% CI** | ***P*** | **Beta** | **95% CI** | ***P*** |
| **Parental occupation** | Modern professional | 304 | Ref | Ref | Ref | Ref | Ref | Ref | Ref | Ref | Ref | Ref | Ref | Ref | Ref | Ref | Ref | Ref | Ref | Ref |
| | Clerical and intermediate | 276 | -0.13 | -0.29, 0.04 | 1.25e-01 | -0.05 | -0.21, 0.11 | 5.48e-01 | -0.08 | -0.24, 0.08 | 3.32e-01 | -0.09 | -0.25, 0.07 | 2.66e-01 | 0.01 | -0.16, 0.17 | 9.42e-01 | 0.09 | -0.08, 0.25 | 2.98e-01 |
| | Sr. managers/administrators | 156 | -0.20 | -0.40, -0.01 | 3.81e-02 | -0.09 | -0.28, 0.10 | 3.62e-01 | -0.06 | -0.25, 0.13 | 5.53e-01 | -0.07 | -0.26, 0.12 | 4.75e-01 | -0.12 | -0.31, 0.08 | 2.39e-01 | -0.02 | -0.21, 0.17 | 8.08e-01 |
| | Technical and craft | 455 | -0.15 | -0.29, -0.00 | 4.52e-02 | -0.04 | -0.18, 0.10 | 5.92e-01 | -0.05 | -0.19, 0.10 | 5.24e-01 | -0.01 | -0.15, 0.13 | 8.92e-01 | 0.00 | -0.14, 0.15 | 9.77e-01 | 0.12 | -0.02, 0.26 | 1.04e-01 |
| | Semi-routine manual/service | 641 | -0.15 | -0.29, -0.01 | 3.32e-02 | -0.08 | -0.22, 0.06 | 2.44e-01 | -0.04 | -0.17, 0.10 | 5.93e-01 | 0.06 | -0.07, 0.20 | 3.72e-01 | 0.04 | -0.10, 0.18 | 5.73e-01 | 0.22 | 0.08, 0.35 | 1.49e-03 |
| | Routine manual and service | 774 | -0.11 | -0.24, 0.03 | 1.12e-01 | -0.06 | -0.19, 0.07 | 3.67e-01 | -0.03 | -0.16, 0.10 | 6.36e-01 | -0.03 | -0.17, 0.10 | 6.28e-01 | 0.06 | -0.08, 0.19 | 3.92e-01 | 0.19 | 0.06, 0.32 | 4.02e-03 |
| | Middle or junior managers | 261 | -0.16 | -0.33, 0.00 | 5.22e-02 | -0.02 | -0.18, 0.15 | 8.37e-01 | -0.06 | -0.22, 0.10 | 4.73e-01 | -0.07 | -0.23, 0.10 | 4.20e-01 | 0.01 | -0.16, 0.18 | 9.00e-01 | -0.01 | -0.18, 0.15 | 8.68e-01 |
| | Traditional professional | 162 | -0.22 | -0.41, -0.02 | 2.71e-02 | 0.00 | -0.19, 0.19 | 9.89e-01 | -0.01 | -0.20, 0.18 | 9.44e-01 | -0.14 | -0.33, 0.05 | 1.47e-01 | -0.12 | -0.31, 0.07 | 2.28e-01 | -0.16 | -0.35, 0.03 | 9.06e-02 |
| | Self-employed | 477 | -0.19 | -0.33, -0.04 | 1.06e-02 | 0.00 | -0.14, 0.14 | 9.85e-01 | -0.03 | -0.17, 0.12 | 7.18e-01 | -0.01 | -0.15, 0.13 | 9.08e-01 | -0.03 | -0.18, 0.11 | 6.51e-01 | 0.05 | -0.10, 0.19 | 5.35e-01 |
| | Student/in training | 204 | -0.08 | -0.26, 0.09 | 3.57e-01 | -0.04 | -0.22, 0.13 | 6.43e-01 | 0.04 | -0.14, 0.22 | 6.63e-01 | -0.15 | -0.33, 0.02 | 9.22e-02 | 0.00 | -0.17, 0.18 | 9.56e-01 | 0.04 | -0.13, 0.22 | 6.18e-01 |
| | Unemployed/sick leave | 247 | -0.21 | -0.38, -0.04 | 1.31e-02 | -0.08 | -0.25, 0.08 | 3.23e-01 | 0.01 | -0.15, 0.18 | 8.71e-01 | 0.05 | -0.12, 0.22 | 5.60e-01 | 0.09 | -0.08, 0.26 | 2.80e-01 | 0.17 | 0.00, 0.34 | 4.62e-02 |
| | Unknown | 53 | -0.08 | -0.38, 0.21 | 5.95e-01 | -0.09 | -0.38, 0.20 | 5.41e-01 | -0.07 | -0.36, 0.22 | 6.39e-01 | 0.04 | -0.26, 0.33 | 8.01e-01 | -0.00 | -0.30, 0.29 | 9.77e-01 | 0.21 | -0.08, 0.50 | 1.49e-01 |
| **Maternal education** | Higher than A-level | 1022 | 0.01 | -0.08, 0.10 | 8.64e-01 | 0.07 | -0.02, 0.16 | 1.25e-01 | 0.05 | -0.04, 0.13 | 3.23e-01 | -0.10 | -0.19, -0.01 | 3.31e-02 | -0.11 | -0.20, -0.02 | 2.23e-02 | -0.25 | -0.34, -0.16 | 3.42e-08 |
| | A-level equivalent | 674 | -0.02 | -0.11, 0.08 | 7.65e-01 | -0.02 | -0.12, 0.08 | 7.01e-01 | -0.04 | -0.14, 0.06 | 4.54e-01 | -0.08 | -0.18, 0.02 | 1.08e-01 | -0.09 | -0.19, 0.01 | 8.81e-02 | -0.03 | -0.13, 0.07 | 5.43e-01 |
| | 5 GCSE equivalent | 1325 | -0.02 | -0.10, 0.07 | 6.82e-01 | 0.04 | -0.05, 0.12 | 4.00e-01 | 0.02 | -0.07, 0.10 | 6.75e-01 | 0.00 | -0.08, 0.09 | 9.07e-01 | -0.03 | -0.12, 0.05 | 4.77e-01 | 0.03 | -0.05, 0.11 | 4.72e-01 |
| | <5 GCSE equivalent | 943 | Ref | Ref | Ref | Ref | Ref | Ref | Ref | Ref | Ref | Ref | Ref | Ref | Ref | Ref | Ref | Ref | Ref | Ref |
| **Paternal education** | Higher than A-level) | 980 | 0.04 | -0.06, 0.14 | 4.17e-01 | 0.05 | -0.05, 0.15 | 3.32e-01 | 0.05 | -0.05, 0.15 | 2.99e-01 | 0.01 | -0.09, 0.11 | 8.47e-01 | -0.08 | -0.18, 0.02 | 1.35e-01 | -0.17 | -0.27, -0.07 | 5.52e-04 |
| | A-level equivalent | 455 | 0.07 | -0.05, 0.20 | 2.23e-01 | 0.06 | -0.05, 0.18 | 2.85e-01 | 0.01 | -0.11, 0.13 | 8.75e-01 | 0.01 | -0.11, 0.13 | 8.71e-01 | -0.06 | -0.18, 0.06 | 3.30e-01 | -0.14 | -0.25, -0.02 | 2.29e-02 |
| | 5 GCSE equivalent | 1060 | 0.00 | -0.09, 0.10 | 9.37e-01 | -0.06 | -0.16, 0.04 | 2.13e-01 | -0.04 | -0.13, 0.06 | 4.76e-01 | -0.02 | -0.11, 0.08 | 7.38e-01 | -0.03 | -0.13, 0.07 | 5.45e-01 | 0.03 | -0.07, 0.12 | 5.79e-01 |
| | <5 GCSE equivalent | 664 | Ref | Ref | Ref | Ref | Ref | Ref | Ref | Ref | Ref | Ref | Ref | Ref | Ref | Ref | Ref | Ref | Ref | Ref |
| **Maternal smoking in pregnancy** | No | 3502 | Ref | Ref | Ref | Ref | Ref | Ref | Ref | Ref | Ref | Ref | Ref | Ref | Ref | Ref | Ref | Ref | Ref | Ref |
| | Yes | 759 | -0.02 | -0.10, 0.06 | 5.87e-01 | -0.08 | -0.16, -0.00 | 3.90e-02 | -0.05 | -0.13, 0.03 | 2.00e-01 | 0.01 | -0.07, 0.09 | 7.39e-01 | 0.12 | 0.04, 0.20 | 2.82e-03 | 0.02 | -0.06, 0.10 | 6.13e-01 |
| **Parity** | | 4166 | -0.00 | -0.03, 0.02 | 8.74e-01 | 0.00 | -0.02, 0.03 | 8.33e-01 | -0.00 | -0.03, 0.02 | 8.62e-01 | 0.01 | -0.02, 0.03 | 4.92e-01 | -0.00 | -0.02, 0.02 | 9.40e-01 | 0.10 | 0.08, 0.13 | 2.15e-17 |
| **Maternal age (years)** | | 4267 | 0.00 | -0.00, 0.01 | 5.46e-01 | 0.00 | -0.01, 0.01 | 9.21e-01 | -0.00 | -0.01, 0.00 | 4.44e-01 | -0.00 | -0.01, 0.00 | 1.64e-01 | -0.01 | -0.02, -0.00 | 5.54e-04 | -0.00 | -0.01, 0.00 | 6.25e-01 |
| **Paternal age (years)** | | 1371 | 0.00 | -0.01, 0.01 | 7.64e-01 | -0.00 | -0.01, 0.00 | 3.16e-01 | -0.00 | -0.01, 0.01 | 9,02e-01 | -0.00 | -0.01, 0.01 | 8.00e-01 | -0.00 | -0.01, 0.01 | 6.83e-01 | 0.00 | -0.00, 0.01 | 3.26e-01 |
| **Paternal BMI ($kg/m^2$)** | | 1114 | 0.02 | -0.03, 0.08 | 4.20e-01 | 0.01 | -0.04, 0.07 | 6.51e-01 | 0.00 | -0.06, 0.06 | 8.88e-01 | -0.00 | -0.06, 0.06 | 9.66e-01 | 0.01 | -0.05, 0.07 | 7.52e-01 | 0.04 | 0.02, 0.05 | 1.02e-09 |

Abbreviations are as for **Figure 1**

### Supplementary information S24: Confounder adjusted MV estimates for the association between maternal BMI and offspring outcomes

| Cohort | Outcome | *N* | Model 1 | | | Model 2 | | | Model 3 | | | $P_{sex\ int.}$ |
|---|---|---|---|---|---|---|---|---|---|---|---|---|
| | | | **β** | **95% CI** | ***P*** | **β** | **95% CI** | ***P*** | **β** | **95% CI** | ***P*** | |
| ALSPAC | BW | 3265 | 0.12 | 0.09, 0.15 | 8.1e-12 | 0.12 | 0.09, 0.16 | 1e-12 | 0.12 | 0.08, 0.15 | 1.8e-11 | 0.79 |
| | 1yr BMI | 3145 | 0.07 | 0.04, 0.11 | 7.4e-05 | 0.07 | 0.03, 0.11 | 1.1e-04 | 0.06 | 0.02, 0.09 | 2.1e-03 | 0.81 |
| | 4yr BMI | 3060 | 0.20 | 0.16, 0.23 | 4.2e-28 | 0.20 | 0.17, 0.24 | 4.7e-29 | 0.18 | 0.15, 0.22 | 7.1e-24 | 0.13 |
| | 10yr BMI | 3007 | 0.32 | 0.29, 0.36 | 7.1e-77 | 0.33 | 0.29, 0.36 | 4.5e-76 | 0.30 | 0.26, 0.33 | 1.2e-63 | 0.99 |
| | 15yr BMI | 2795 | 0.36 | 0.32, 0.39 | 4.6e-88 | 0.36 | 0.32, 0.39 | 1.8e-85 | 0.32 | 0.29, 0.36 | 3.1e-72 | 0.17 |
| | 10yr FMI | 2627 | 0.31 | 0.28, 0.35 | 2.8e-61 | 0.31 | 0.27, 0.35 | 3.5e-59 | 0.28 | 0.25, 0.32 | 3.9e-50 | 0.41 |
| | 12yr FMI | 2598 | 0.33 | 0.30, 0.37 | 1.8e-68 | 0.33 | 0.29, 0.36 | 2.1e-65 | 0.30 | 0.26, 0.34 | 9.2e-56 | 0.75 |
| | 14yr FMI | 2424 | 0.33 | 0.30, 0.37 | 3.9e-64 | 0.33 | 0.29, 0.37 | 5.8e-61 | 0.30 | 0.26, 0.34 | 5.5e-52 | 0.06 |
| | 16yr FMI | 2105 | 0.35 | 0.31, 0.39 | 4.1e-62 | 0.34 | 0.30, 0.38 | 1.2e-58 | 0.31 | 0.27, 0.35 | 1.3e-49 | 0.46 |
| | 18yr FMI | 1884 | 0.35 | 0.31, 0.39 | 4.2e-55 | 0.34 | 0.30, 0.38 | 2e-51 | 0.32 | 0.27, 0.36 | 4.6e-45 | 0.31 |
| BiB (SA) | BW | 449 | 0.16 | 0.06, 0.25 | 1e-03 | 0.13 | 0.03, 0.22 | 0.01 | 0.13 | 0.03, 0.22 | 0.01 | 0.24 |
| | 1yr BMI | 401 | 0.16 | 0.06, 0.26 | 2.2e-03 | 0.14 | 0.04, 0.25 | 0.01 | 0.13 | 0.03, 0.24 | 0.01 | 0.40 |
| | 4yr BMI | 325 | 0.26 | 0.14, 0.37 | 2.1e-05 | 0.25 | 0.13, 0.38 | 6.5e-05 | 0.22 | 0.10, 0.34 | 4e-04 | 0.32 |
| BiB (WE) | BW | 604 | 0.20 | 0.13, 0.27 | 4.4e-08 | 0.21 | 0.14, 0.29 | 1.4e-08 | 0.20 | 0.13, 0.28 | 1.3e-07 | 0.22 |
| | 1yr BMI | 559 | 0.11 | 0.03, 0.19 | 0.01 | 0.12 | 0.04, 0.21 | 3.5e-03 | 0.11 | 0.03, 0.20 | 0.01 | 0.70 |
| | 4yr BMI | 442 | 0.15 | 0.06, 0.23 | 6.6e-04 | 0.17 | 0.08, 0.26 | 1.4e-04 | 0.14 | 0.05, 0.23 | 2e-03 | 0.36 |

**SA**: South Asians, **WE**: White Europeans, **Model 1**: controlled for maternal age, offspring age and sex in the standardised exposure and outcome, **Model 2**: additionally adjusted for potential confounders including parity, maternal smoking during pregnancy, maternal and paternal education and parental occupation, **Model 3**: additionally adjusted for paternal BMI, $P_{sex\ int.}$: *P*-value for exposure * sex interaction (covariates as per model 3), **β**: coefficient from linear regression of outcome (age- [except for BW] and sex-standardised *z*-score) on maternal BMI (age-standardised *z*-score)

**Supplementary information S25: Confounder adjusted MV estimates for the association between maternal BMI and offspring outcomes, *additionally adjusted for gestational age at birth***

| Cohort | Outcome | *N* | Model 1 | | | Model 2 | | | Model 3 | | | $P_{sex\ int.}$ |
|---|---|---|---|---|---|---|---|---|---|---|---|---|
| | | | β | 95% CI | *P* | β | 95% CI | *P* | β | 95% CI | *P* | |
| ALSPAC | BW | 3265 | 0.11 | 0.08, 0.14 | 4.1e-14 | 0.12 | 0.09, 0.15 | 3e-15 | 0.11 | 0.08, 0.14 | 1.6e-13 | 0.47 |
| | 1yr BMI | 3145 | 0.07 | 0.04, 0.11 | 7.4e-05 | 0.07 | 0.03, 0.10 | 1.2e-04 | 0.06 | 0.02, 0.09 | 2.1e-03 | 0.98 |
| | 4yr BMI | 3060 | 0.20 | 0.16, 0.23 | 4.7e-28 | 0.20 | 0.17, 0.24 | 5.2e-29 | 0.18 | 0.15, 0.22 | 8.1e-24 | 0.14 |
| | 10yr BMI | 3007 | 0.32 | 0.29, 0.36 | 1.1e-76 | 0.33 | 0.29, 0.36 | 7.6e-76 | 0.29 | 0.26, 0.33 | 2.4e-63 | 0.70 |
| | 15yr BMI | 2795 | 0.36 | 0.32, 0.39 | 5.5e-88 | 0.36 | 0.32, 0.39 | 2.2e-85 | 0.32 | 0.29, 0.36 | 4.1e-72 | 0.42 |
| | 10yr FMI | 2627 | 0.31 | 0.28, 0.35 | 4.6e-61 | 0.31 | 0.27, 0.35 | 5.9e-59 | 0.28 | 0.25, 0.32 | 7.7e-50 | 0.97 |
| | 12yr FMI | 2598 | 0.33 | 0.30, 0.37 | 2e-68 | 0.33 | 0.29, 0.36 | 2.4e-65 | 0.30 | 0.26, 0.34 | 1.2e-55 | 0.78 |
| | 14yr FMI | 2424 | 0.33 | 0.30, 0.37 | 4.2e-64 | 0.33 | 0.29, 0.37 | 6.2e-61 | 0.30 | 0.26, 0.34 | 6.4e-52 | 0.78 |
| | 16yr FMI | 2105 | 0.35 | 0.31, 0.39 | 4.3e-62 | 0.34 | 0.30, 0.38 | 1.2e-58 | 0.31 | 0.27, 0.35 | 1.4e-49 | 0.30 |
| | 18yr FMI | 1884 | 0.35 | 0.31, 0.39 | 5e-55 | 0.34 | 0.30, 0.38 | 2.2e-51 | 0.32 | 0.27, 0.36 | 5.5e-45 | 0.33 |
| BiB (SA) | BW | 449 | 0.15 | 0.06, 0.23 | 5.3e-04 | 0.12 | 0.04, 0.21 | 0.01 | 0.12 | 0.03, 0.20 | 0.01 | 0.65 |
| | 1yr BMI | 401 | 0.16 | 0.06, 0.26 | 2.3e-03 | 0.14 | 0.04, 0.25 | 0.01 | 0.13 | 0.03, 0.24 | 0.01 | 0.41 |
| | 4yr BMI | 325 | 0.26 | 0.14, 0.38 | 2.2e-05 | 0.25 | 0.13, 0.38 | 6.7e-05 | 0.22 | 0.10, 0.34 | 4e-04 | 0.59 |
| BiB (WE) | BW | 604 | 0.19 | 0.12, 0.25 | 1e-08 | 0.19 | 0.13, 0.26 | 9.3e-09 | 0.19 | 0.12, 0.25 | 6.2e-08 | 0.04 |
| | 1yr BMI | 559 | 0.11 | 0.03, 0.19 | 0.01 | 0.12 | 0.04, 0.21 | 4.1e-03 | 0.11 | 0.03, 0.20 | 0.01 | 0.84 |
| | 4yr BMI | 442 | 0.15 | 0.06, 0.23 | 6.4e-04 | 0.17 | 0.08, 0.26 | 1.5e-04 | 0.14 | 0.05, 0.23 | 2.2e-03 | 0.34 |

**SA**: South Asians, **WE**: White Europeans, **Model 1**: controlled for maternal age, offspring age and sex in the standardised exposure and outcome and gestational age at birth, **Model 2**: additionally adjusted for potential confounders including parity, maternal smoking during pregnancy, maternal and paternal education and parental occupation, **Model 3**: additionally adjusted for paternal BMI, $P_{sex\ int.}$: *P*-value for exposure * sex interaction (covariates as per model 3), **β**: coefficient from linear regression of outcome (age- [except for BW] and sex-standardised *z*-score) on maternal BMI (age-standardised *z*-score)

**Supplementary information S26: Confounder adjusted MV estimates for the association between maternal BMI and offspring outcomes, *additionally adjusted for 20 genetic principal components***

| Cohort | Outcome | *N* | Model 1 | | | Model 2 | | | Model 3 | | | $P_{sex\ int.}$ |
|---|---|---|---|---|---|---|---|---|---|---|---|---|
| | | | β | 95% CI | *P* | β | 95% CI | *P* | β | 95% CI | *P* | |
| ALSPAC | BW | 3265 | 0.12 | 0.08, 0.15 | 1.3e-11 | 0.12 | 0.09, 0.16 | 2.2e-12 | 0.12 | 0.08, 0.15 | 3.5e-11 | 0.93 |
| | 1yr BMI | 3145 | 0.07 | 0.03, 0.10 | 1.1e-04 | 0.07 | 0.03, 0.10 | 1.5e-04 | 0.06 | 0.02, 0.09 | 2.5e-03 | 0.05 |
| | 4yr BMI | 3060 | 0.20 | 0.16, 0.23 | 5.9e-28 | 0.20 | 0.17, 0.24 | 6.6e-29 | 0.18 | 0.15, 0.22 | 7.4e-24 | 0.62 |
| | 10yr BMI | 3007 | 0.32 | 0.29, 0.36 | 3.2e-76 | 0.33 | 0.29, 0.36 | 2.2e-75 | 0.30 | 0.26, 0.33 | 2.6e-63 | 0.98 |
| | 15yr BMI | 2795 | 0.35 | 0.32, 0.39 | 8e-86 | 0.35 | 0.32, 0.39 | 2.7e-83 | 0.32 | 0.28, 0.35 | 1.5e-70 | 0.97 |
| | 10yr FMI | 2627 | 0.31 | 0.28, 0.35 | 3.1e-60 | 0.31 | 0.27, 0.35 | 3.6e-58 | 0.28 | 0.25, 0.32 | 2e-49 | 0.94 |
| | 12yr FMI | 2598 | 0.33 | 0.29, 0.37 | 4.6e-67 | 0.32 | 0.29, 0.36 | 3.7e-64 | 0.30 | 0.26, 0.34 | 6.2e-55 | 0.77 |
| | 14yr FMI | 2424 | 0.33 | 0.30, 0.37 | 3.2e-63 | 0.33 | 0.29, 0.37 | 3.5e-60 | 0.30 | 0.26, 0.34 | 1.4e-51 | 0.09 |
| | 16yr FMI | 2105 | 0.35 | 0.31, 0.39 | 3.3e-61 | 0.34 | 0.30, 0.38 | 5.7e-58 | 0.31 | 0.27, 0.35 | 3.7e-49 | 0.76 |
| | 18yr FMI | 1884 | 0.35 | 0.30, 0.39 | 3.7e-53 | 0.34 | 0.29, 0.38 | 1.1e-49 | 0.31 | 0.27, 0.36 | 8.4e-44 | 0.41 |
| BiB (SA) | BW | 449 | 0.15 | 0.06, 0.25 | 1.6e-03 | 0.13 | 0.03, 0.22 | 0.01 | 0.13 | 0.03, 0.22 | 0.01 | 0.69 |
| | 1yr BMI | 401 | 0.15 | 0.05, 0.26 | 4.3e-03 | 0.14 | 0.03, 0.24 | 0.01 | 0.13 | 0.02, 0.23 | 0.02 | 0.04 |
| | 4yr BMI | 325 | 0.25 | 0.13, 0.37 | 6.2e-05 | 0.25 | 0.12, 0.37 | 1.5e-04 | 0.21 | 0.09, 0.33 | 8.6e-04 | 0.31 |
| BiB (WE) | BW | 604 | 0.21 | 0.14, 0.29 | 1.4e-08 | 0.23 | 0.15, 0.30 | 6.2e-09 | 0.22 | 0.14, 0.29 | 5.9e-08 | 0.23 |
| | 1yr BMI | 559 | 0.11 | 0.03, 0.19 | 0.01 | 0.13 | 0.04, 0.21 | 3.8e-03 | 0.12 | 0.03, 0.20 | 0.01 | 0.48 |
| | 4yr BMI | 442 | 0.14 | 0.05, 0.23 | 1.6e-03 | 0.17 | 0.07, 0.26 | 3.9e-04 | 0.13 | 0.04, 0.23 | 0.01 | 0.83 |

**SA**: South Asians, **WE**: White Europeans, **Model 1**: controlled for maternal age, offspring age and sex in the standardised exposure and outcome and 20 genetic principal components, **Model 2**: additionally adjusted for potential confounders including parity, maternal smoking during pregnancy, maternal and paternal education and parental occupation, **Model 3**: additionally adjusted for paternal BMI, $P_{sex\ int.}$: *P*-value for exposure * sex interaction (covariates as per model 3), **β**: coefficient from linear regression of outcome (age- [except for BW] and sex-standardised *z*-score) on maternal BMI (age-standardised *z*-score)

**Supplementary information S27: Confounder adjusted MV estimates for the association between maternal BMI and *offspring weight, BMI and ponderal index (PI) at birth* in ALSPAC**

| Cohort | Outcome | $N^a$ | Model 1 | | | Model 2 | | | Model 3 | | | $P_{sex\ int.}$ |
|---|---|---|---|---|---|---|---|---|---|---|---|---|
| | | | β | 95% CI | *P* | β | 95% CI | *P* | β | 95% CI | *P* | |
| ALSPAC | BW | 2635 | 0.11 | 0.07, 0.14 | 6.3e-09 | 0.11 | 0.08, 0.15 | 5.8e-10 | 0.11 | 0.07, 0.15 | 5.5e-09 | 0.87 |
| | Birth BMI | 2635 | 0.12 | 0.08, 0.15 | 1.7e-09 | 0.12 | 0.08, 0.16 | 6.7e-10 | 0.11 | 0.08, 0.15 | 2.7e-09 | 0.38 |
| | Birth PI | 2635 | 0.09 | 0.05, 0.13 | 1e-06 | 0.09 | 0.05, 0.13 | 1.5e-06 | 0.09 | 0.05, 0.13 | 2.2e-06 | 0.25 |

**Model 1**: controlled for maternal age, offspring age and sex in the standardised exposure and outcome, **Model 2**: additionally adjusted for potential confounders including parity, maternal smoking during pregnancy, maternal and paternal education and parental occupation, **Model 3**: additionally adjusted for paternal BMI, $P_{sex\ int.}$: *P*-value for exposure * sex interaction (covariates as per model 3), **a**: individuals with missing birth length data were excluded from all analyses to enable comparability between phenotypes, **β**: coefficient from linear regression of outcome (age- [except for BW] and sex-standardised *z*-score) on maternal BMI (age-standardised *z*-score)

**Supplementary information S28: MR and MV estimates from cohorts analysed separately**

| Outcome | Estimator | *N* | Effect | 95% CI | *P* | $P_{dif}$ |
|---|---|---|---|---|---|---|
| BW | MV | 3265 | 0.12 | 0.08, 0.15 | 1.8e-11 | |
| | Lassosum (MR) | 5085 | 0.11 | 0.01, 0.22 | 0.03 | 0.89 |
| | Yengo (MR) | 5085 | 0.19 | 0.01, 0.36 | 0.04 | 0.43 |
| | Locke (MR) | 5085 | 0.15 | -0.12, 0.41 | 0.28 | 0.85 |
| | Speliotes (MR) | 5085 | 0.31 | 0.01, 0.60 | 0.04 | 0.22 |
| | FTO (MR) | 5085 | 0.09 | -0.35, 0.54 | 0.68 | 0.92 |
| 1yr BMI | MV | 3145 | 0.06 | 0.02, 0.09 | 2.1e-03 | |
| | Lassosum (MR) | 4838 | 0.00 | -0.11, 0.10 | 0.95 | 0.27 |
| | Yengo (MR) | 4838 | 0.13 | -0.05, 0.31 | 0.16 | 0.43 |
| | Locke (MR) | 4838 | 0.35 | 0.05, 0.65 | 0.02 | 0.07 |
| | Speliotes (MR) | 4838 | 0.22 | -0.09, 0.54 | 0.17 | 0.31 |
| | FTO (MR) | 4838 | 0.19 | -0.31, 0.69 | 0.45 | 0.66 |
| 4yr BMI | MV | 3060 | 0.18 | 0.15, 0.22 | 7.1e-24 | |
| | Lassosum (MR) | 4670 | 0.02 | -0.09, 0.13 | 0.77 | 2.6e-03 |
| | Yengo (MR) | 4670 | 0.14 | -0.05, 0.32 | 0.14 | 0.61 |
| | Locke (MR) | 4670 | 0.17 | -0.10, 0.43 | 0.22 | 0.89 |
| | Speliotes (MR) | 4670 | 0.23 | -0.06, 0.52 | 0.13 | 0.77 |
| | FTO (MR) | 4670 | 0.21 | -0.25, 0.66 | 0.37 | 0.92 |
| 10yr BMI | MV | 3007 | 0.30 | 0.26, 0.33 | 1.2e-63 | |
| | Lassosum (MR) | 4476 | 0.10 | -0.01, 0.21 | 0.07 | 1.4e-04 |
| | Yengo (MR) | 4476 | 0.10 | -0.09, 0.29 | 0.29 | 0.04 |
| | Locke (MR) | 4476 | 0.05 | -0.21, 0.32 | 0.69 | 0.08 |
| | Speliotes (MR) | 4476 | 0.13 | -0.14, 0.40 | 0.36 | 0.24 |
| | FTO (MR) | 4476 | -0.01 | -0.44, 0.43 | 0.97 | 0.25 |
| 15yr BMI | MV | 2795 | 0.32 | 0.29, 0.36 | 3.1e-72 | |
| | Lassosum (MR) | 4112 | 0.13 | 0.01, 0.24 | 0.03 | 1e-03 |
| | Yengo (MR) | 4112 | 0.13 | -0.06, 0.33 | 0.17 | 0.05 |
| | Locke (MR) | 4112 | -0.13 | -0.45, 0.19 | 0.43 | 0.01 |
| | Speliotes (MR) | 4112 | -0.16 | -0.51, 0.18 | 0.35 | 0.01 |
| | FTO (MR) | 4112 | -0.49 | -1.10, 0.12 | 0.11 | 0.01 |

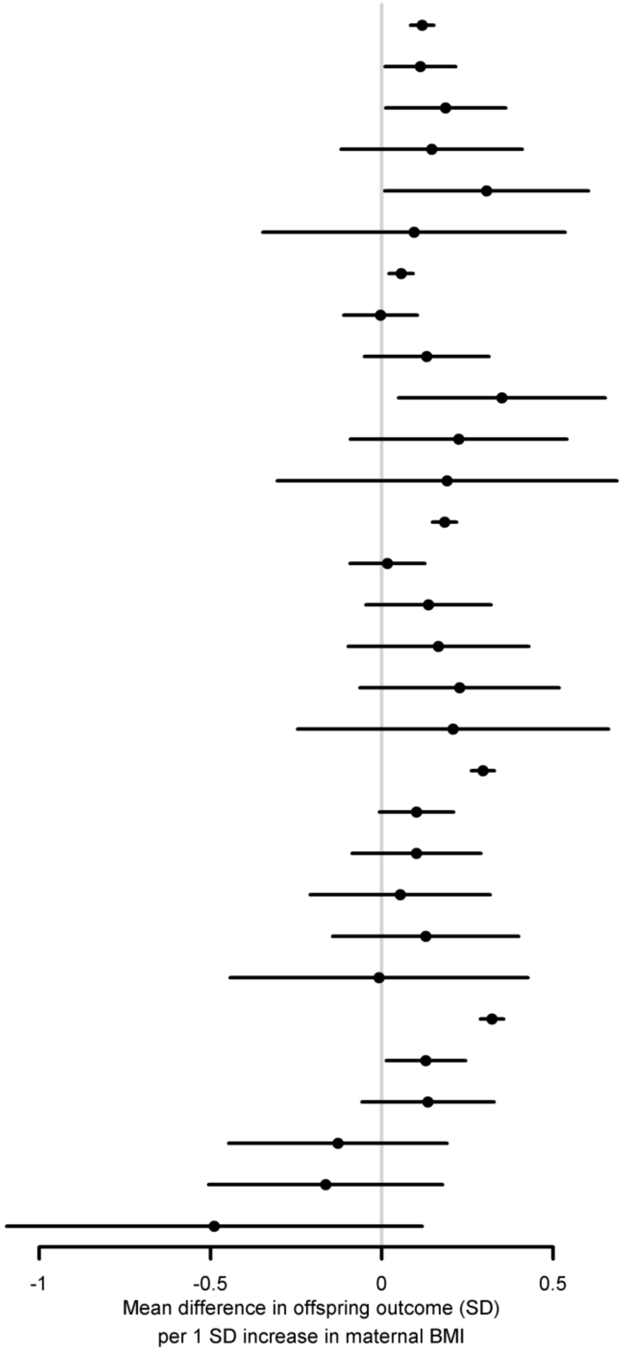

Comparison of the MR and MV estimates for all genetic IVs for ***ALSPAC, for BW and BMI***. MV estimates were from model three (**Methods**). The exposure was age standardised maternal BMI *z*-score and the outcomes were sex and age (except for BW) standardised *z*-scores for offspring phenotype. ***P***: *P*-value for the null hypothesis that the effect equals zero, $\boldsymbol{P_{dif}}$: *P*-value for the null hypothesis that MR effect equals the MV effect, **FTO**: rs9939609 at the *FTO* locus, **Speliotes, Locke, Yengo**: GWS SNPs from the GWAS with

| Outcome | Estimator | *N* | Effect | 95% CI | *P* | $P_{dif}$ |
|---|---|---|---|---|---|---|
| Birth weight | MV | 2635 | 0.11 | 0.07, 0.15 | 5.5e-09 | |
| | Lassosum (MR) | 4113 | 0.13 | 0.02, 0.24 | 0.02 | 0.73 |
| | Yengo (MR) | 4113 | 0.10 | -0.09, 0.30 | 0.30 | 0.95 |
| | Locke (MR) | 4113 | 0.11 | -0.21, 0.43 | 0.50 | 1.00 |
| | Speliotes (MR) | 4113 | 0.21 | -0.18, 0.59 | 0.30 | 0.66 |
| | FTO (MR) | 4113 | -0.03 | -0.65, 0.59 | 0.93 | 0.79 |
| Birth BMI | MV | 2635 | 0.11 | 0.08, 0.15 | 2.7e-09 | |
| | Lassosum (MR) | 4113 | 0.10 | -0.02, 0.21 | 0.10 | 0.77 |
| | Yengo (MR) | 4113 | 0.04 | -0.16, 0.24 | 0.71 | 0.46 |
| | Locke (MR) | 4113 | 0.05 | -0.28, 0.38 | 0.76 | 0.72 |
| | Speliotes (MR) | 4113 | 0.12 | -0.27, 0.52 | 0.54 | 0.96 |
| | FTO (MR) | 4113 | -0.39 | -1.11, 0.33 | 0.29 | 0.60 |
| Birth PI | MV | 2635 | 0.09 | 0.05, 0.13 | 2.2e-06 | |
| | Lassosum (MR) | 4113 | 0.06 | -0.06, 0.17 | 0.34 | 0.57 |
| | Yengo (MR) | 4113 | -0.01 | -0.21, 0.18 | 0.90 | 0.33 |
| | Locke (MR) | 4113 | 0.00 | -0.32, 0.32 | 0.99 | 0.61 |
| | Speliotes (MR) | 4113 | 0.03 | -0.36, 0.42 | 0.87 | 0.78 |
| | FTO (MR) | 4113 | -0.55 | -1.31, 0.20 | 0.15 | 0.34 |

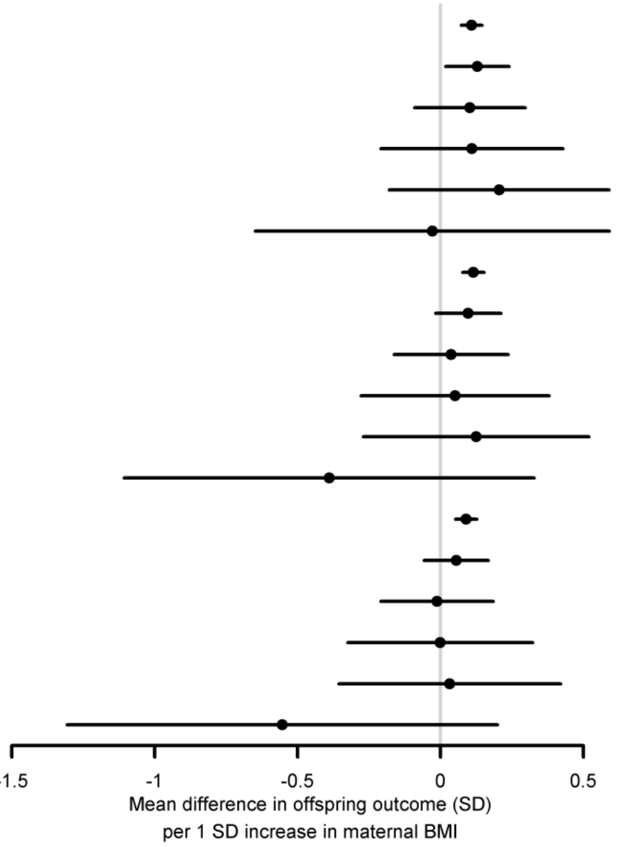

Comparison of the MR and MV estimates for all genetic IVs for ***ALSPAC, for weight, BMI and ponderal index (PI) at birth***. MV estimates were from model three (**Methods**). The exposure was age standardised maternal BMI *z*-score and the outcomes were sex and age (except for BW) standardised *z*-scores for offspring phenotype. ***P***: *P*-value for the null hypothesis that the effect equals zero, $\boldsymbol{P_{dif}}$: *P*-value for the null hypothesis that MR effect equals the MV effect, **FTO**: rs9939609 at the *FTO* locus, **Speliotes, Locke, Yengo**: GWS SNPs from the GWAS with the indicated first author, **Lassosum**: PRS calculated by the lassosum method

| Outcome | Estimator | *N* | Effect | 95% CI | *P* | $P_{dif}$ |
|---|---|---|---|---|---|---|
| BW | MV | 449 | 0.13 | 0.03, 0.22 | 0.01 | |
| | Lassosum (MR) | 2262 | 0.17 | -0.04, 0.38 | 0.12 | 0.76 |
| | Yengo (MR) | 2262 | 0.11 | -0.18, 0.41 | 0.45 | 0.91 |
| | Locke (MR) | 2262 | -0.02 | -0.37, 0.33 | 0.90 | 0.42 |
| | Speliotes (MR) | 2262 | 0.01 | -0.30, 0.32 | 0.93 | 0.49 |
| | FTO (MR) | 2262 | 0.03 | -0.41, 0.47 | 0.90 | 0.68 |
| 1yr BMI | MV | 401 | 0.13 | 0.03, 0.24 | 0.01 | |
| | Lassosum (MR) | 2023 | -0.14 | -0.36, 0.08 | 0.20 | 0.02 |
| | Yengo (MR) | 2023 | 0.04 | -0.25, 0.34 | 0.78 | 0.60 |
| | Locke (MR) | 2023 | 0.05 | -0.33, 0.43 | 0.80 | 0.68 |
| | Speliotes (MR) | 2023 | 0.04 | -0.30, 0.38 | 0.82 | 0.64 |
| | FTO (MR) | 2023 | -0.35 | -0.83, 0.13 | 0.15 | 0.10 |
| 4yr BMI | MV | 325 | 0.22 | 0.10, 0.34 | 4e-04 | |
| | Lassosum (MR) | 1566 | -0.06 | -0.28, 0.16 | 0.60 | 0.02 |
| | Yengo (MR) | 1566 | -0.08 | -0.44, 0.27 | 0.65 | 0.12 |
| | Locke (MR) | 1566 | -0.29 | -0.76, 0.17 | 0.22 | 0.06 |
| | Speliotes (MR) | 1566 | -0.04 | -0.38, 0.29 | 0.80 | 0.17 |
| | FTO (MR) | 1566 | -0.36 | -0.99, 0.26 | 0.26 | 0.16 |

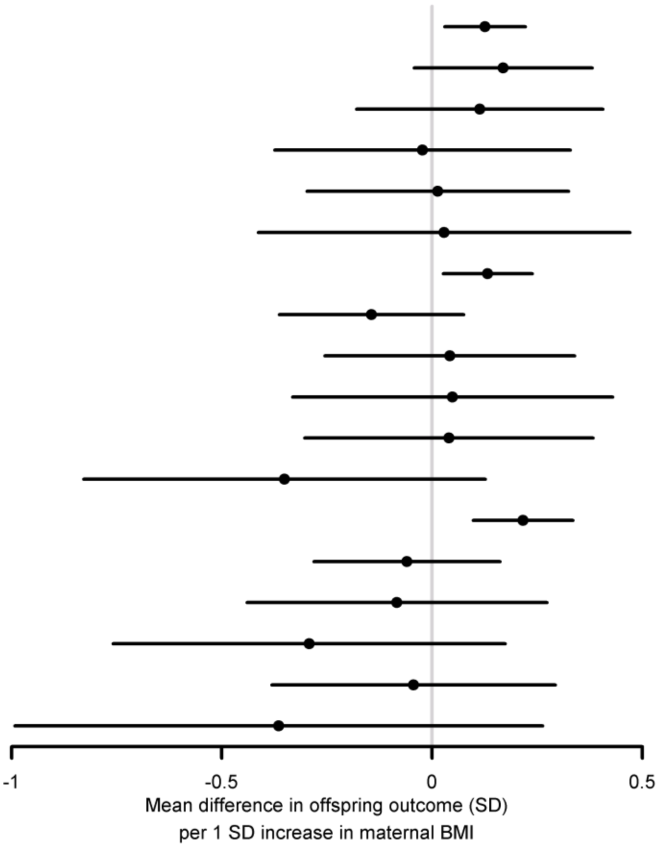

Comparison of the MR and MV estimates for all genetic IVs for ***BiB (South Asians)***. MV estimates were from model three (**Methods**). The exposure was age standardised maternal BMI *z*-score and the outcomes were sex and age (except for BW) standardised *z*-scores for offspring phenotype. ***P***: *P*-value for the null hypothesis that the effect equals zero, $\boldsymbol{P_{dif}}$: *P*-value for the null hypothesis that MR effect equals the MV effect, **FTO**: rs9939609 at the *FTO* locus, **Speliotes, Locke, Yengo**: GWS SNPs from the GWAS with the indicated first author, **Lassosum**: PRS calculated by the lassosum method

| Outcome | Estimator | *N* | Effect | 95% CI | *P* | $P_{dif}$ |
|---|---|---|---|---|---|---|
| BW | MV | 604 | 0.20 | 0.13, 0.28 | 1.3e-07 | |
| | Lassosum (MR) | 1992 | 0.19 | 0.00, 0.38 | 0.05 | 1.00 |
| | Yengo (MR) | 1992 | 0.21 | -0.11, 0.53 | 0.21 | 0.94 |
| | Locke (MR) | 1992 | 0.51 | 0.09, 0.93 | 0.02 | 0.20 |
| | Speliotes (MR) | 1992 | 0.67 | 0.17, 1.17 | 0.01 | 0.16 |
| | FTO (MR) | 1992 | 0.44 | -0.15, 1.03 | 0.15 | 0.55 |
| 1yr BMI | MV | 559 | 0.11 | 0.03, 0.20 | 0.01 | |
| | Lassosum (MR) | 1798 | 0.04 | -0.17, 0.24 | 0.72 | 0.55 |
| | Yengo (MR) | 1798 | 0.18 | -0.17, 0.52 | 0.31 | 0.66 |
| | Locke (MR) | 1798 | 0.25 | -0.21, 0.70 | 0.29 | 0.57 |
| | Speliotes (MR) | 1798 | 0.47 | -0.05, 0.99 | 0.08 | 0.23 |
| | FTO (MR) | 1798 | 0.69 | -0.01, 1.40 | 0.05 | 0.33 |
| 4yr BMI | MV | 442 | 0.14 | 0.05, 0.23 | 2e-03 | |
| | Lassosum (MR) | 1339 | 0.06 | -0.15, 0.28 | 0.55 | 0.66 |
| | Yengo (MR) | 1339 | -0.08 | -0.46, 0.30 | 0.68 | 0.42 |
| | Locke (MR) | 1339 | 0.06 | -0.39, 0.50 | 0.80 | 0.80 |
| | Speliotes (MR) | 1339 | 0.14 | -0.28, 0.56 | 0.51 | 0.95 |
| | FTO (MR) | 1339 | 0.49 | 0.00, 0.99 | 0.05 | 0.22 |

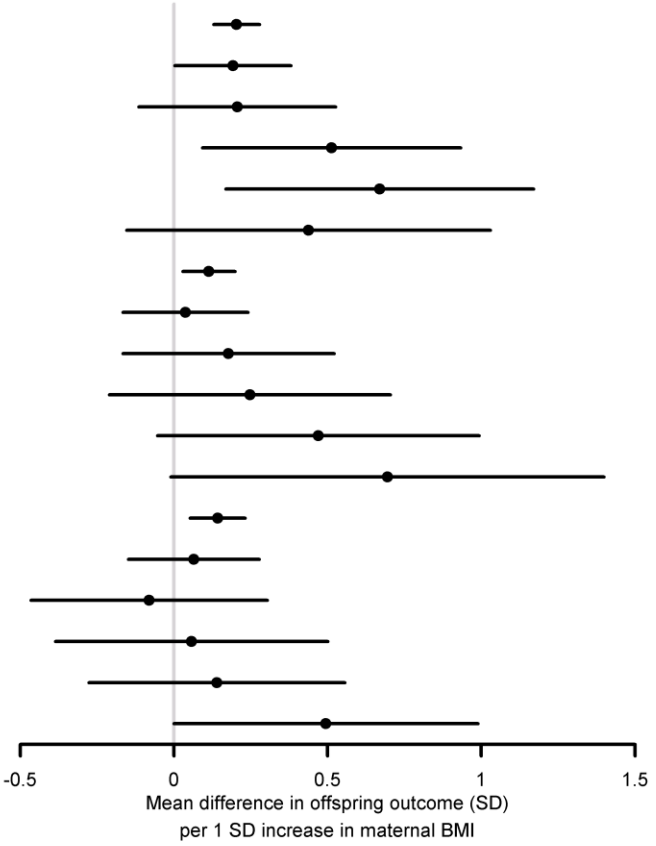

Comparison of the MR and MV estimates for all genetic IVs for ***BiB (White Europeans)***. MV estimates were from model three (**Methods**). The exposure was age standardised maternal BMI *z*-score and the outcomes were sex and age (except for BW) standardised *z*-scores for offspring phenotype. ***P***: *P*-value for the null hypothesis that the effect equals zero, $P_{dif}$: *P*-value for the null hypothesis that MR effect equals the MV effect, **FTO**: rs9939609 at the *FTO* locus, **Speliotes, Locke, Yengo**: GWS SNPs from the GWAS with the indicated first author, **Lassosum**: PRS calculated by the lassosum method

**Supplementary information S29: Meta-analysed results from ALSPAC and BiB from a linear mixed model**

| Phenotype | Estimator | *N* | Effect | 95% CI | *P* | $P_{dif}$ |
|---|---|---|---|---|---|---|
| BW | MV | 4318 | 0.13 | 0.10, 0.16 | 2.2e-18 | |
| | Lassosum (MR) | 9339 | 0.12 | 0.05, 0.19 | 4.9e-04 | 0.80 |
| | Yengo (MR) | 9339 | 0.17 | 0.05, 0.28 | 0.01 | 0.54 |
| | Locke (MR) | 9339 | 0.11 | -0.06, 0.27 | 0.20 | 0.69 |
| | Speliotes (MR) | 9339 | 0.25 | 0.05, 0.45 | 0.02 | 0.20 |
| | FTO (MR) | 9339 | 0.03 | -0.25, 0.31 | 0.83 | 0.41 |
| 1yr BMI | MV | 4105 | 0.07 | 0.04, 0.10 | 1e-05 | |
| | Lassosum (MR) | 8659 | -0.03 | -0.10, 0.05 | 0.47 | 0.03 |
| | Yengo (MR) | 8659 | 0.07 | -0.06, 0.19 | 0.28 | 0.96 |
| | Locke (MR) | 8659 | 0.22 | 0.05, 0.39 | 0.01 | 0.11 |
| | Speliotes (MR) | 8659 | 0.18 | -0.01, 0.37 | 0.07 | 0.21 |
| | FTO (MR) | 8659 | 0.16 | -0.13, 0.45 | 0.29 | 0.57 |
| 4yr BMI | MV | 3827 | 0.18 | 0.15, 0.21 | 6.1e-29 | |
| | Lassosum (MR) | 7575 | 0.02 | -0.06, 0.09 | 0.65 | 2e-04 |
| | Yengo (MR) | 7575 | 0.01 | -0.11, 0.14 | 0.86 | 0.01 |
| | Locke (MR) | 7575 | 0.10 | -0.07, 0.26 | 0.25 | 0.35 |
| | Speliotes (MR) | 7575 | 0.15 | -0.03, 0.34 | 0.10 | 0.80 |
| | FTO (MR) | 7575 | 0.33 | 0.09, 0.58 | 0.01 | 0.27 |

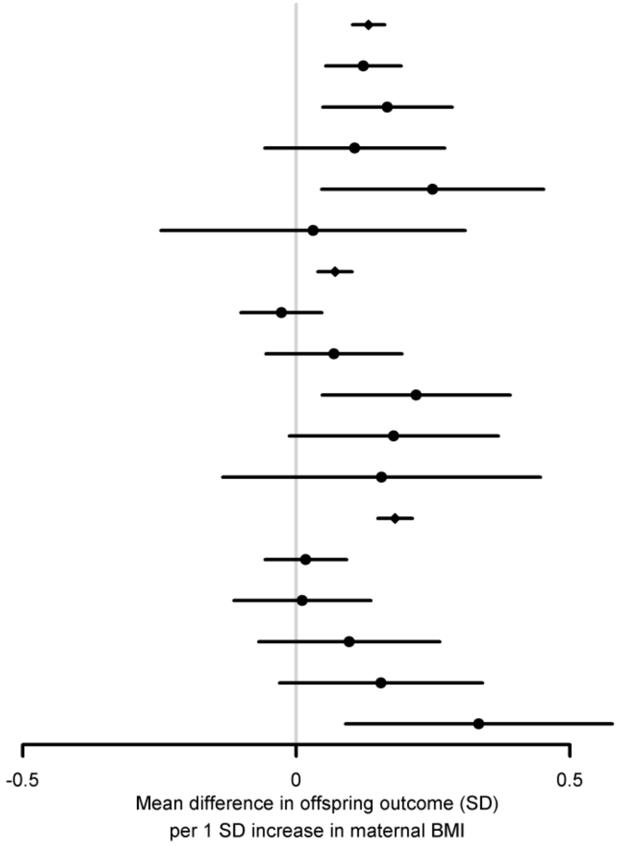

Comparison of the MR and MV estimates for all genetic IVs, ***meta-analysed for ALSPAC, BiB South Asians and BiB White Europeans***, for BW and BMI outcomes, estimated ***using a linear mixed model (LMM)*** to control for population structure (**Methods**). The LMM included 20 genetic PCs fitted as fixed effects. MV estimates were from model three (**Methods**). The exposure was age-standardised maternal BMI *z*-score and the outcomes were sex- and age- (except for BW) standardised *z*-scores for offspring phenotype. ***P***: *P*-value for the null hypothesis that the effect equals zero, $\boldsymbol{P_{dif}}$: *P*-value for the null hypothesis that MR effect equals the MV effect, **FTO**: rs9939609 at the *FTO* locus, **Speliotes, Locke, Yengo**: GWS SNPs from the GWAS with the indicated first author, **Lassosum**: PRS calculated by the lassosum method

**Supplementary information S30: Associations of the lassosum maternal non-transmitted allele BMI PRS (*z*-score) with offspring outcomes (age and sex standardised *z*-score), in ALSPAC, BiB South Asians (SA) and BiB white Europeans (WE)**

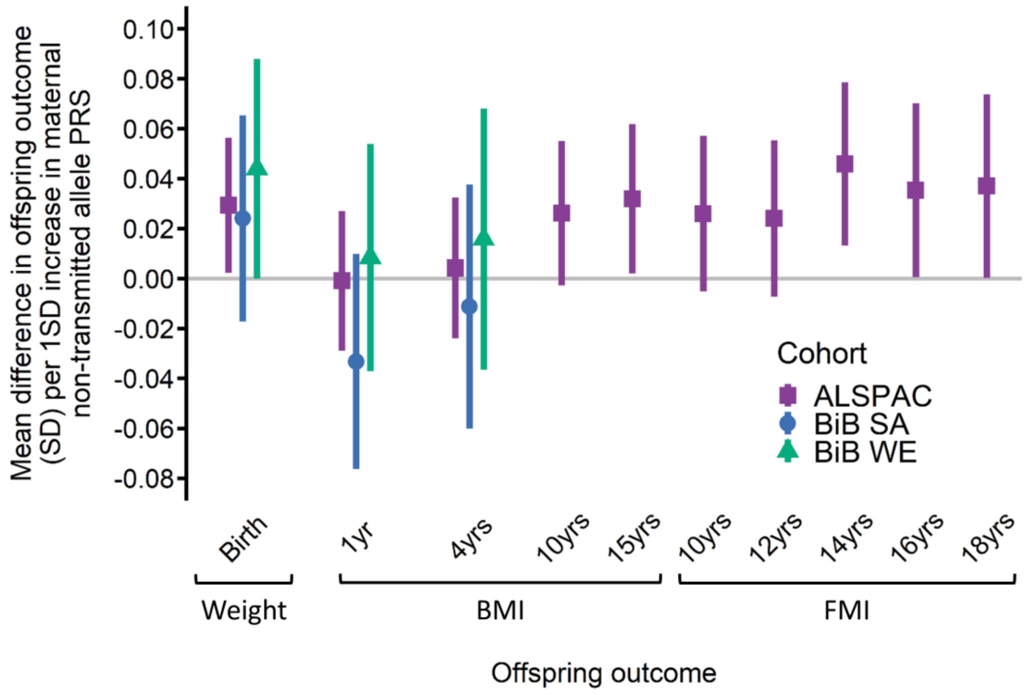

**Supplementary information S31: Associations of the lassosum maternal non-transmitted allele BMI PRS (*z*-score) with offspring outcomes (age and sex standardised *z*-score), in ALSPAC, BiB South Asians (SA) and BiB white Europeans (WE)**

| Outcome | Cohort | *N* | Beta | 95% CI | | *P* |
|---|---|---|---|---|---|---|
| BW | ALSPAC | 5085 | 0.03 | 0.00 | 0.06 | 0.034 |
| | BiB (SA) | 2262 | 0.02 | -0.02 | 0.07 | 0.252 |
| | BiB (WE) | 1992 | 0.04 | 0.00 | 0.09 | 0.050 |
| | Meta-analysis | 9339 | 0.03 | 0.01 | 0.05 | 0.002 |
| 1yr BMI | ALSPAC | 4838 | 0.00 | -0.03 | 0.03 | 0.946 |
| | BiB (SA) | 2023 | -0.03 | -0.08 | 0.01 | 0.130 |
| | BiB (WE) | 1798 | 0.01 | -0.04 | 0.05 | 0.718 |
| | Meta-analysis | 8659 | -0.01 | -0.03 | 0.01 | 0.538 |
| 4yr BMI | ALSPAC | 4670 | 0.00 | -0.02 | 0.03 | 0.771 |
| | BiB (SA) | 1566 | -0.01 | -0.06 | 0.04 | 0.652 |
| | BiB (WE) | 1339 | 0.02 | -0.04 | 0.07 | 0.555 |
| | Meta-analysis | 7575 | 0.00 | -0.02 | 0.03 | 0.784 |
| 10yr BMI | ALSPAC | 4476 | 0.03 | 0.00 | 0.05 | 0.077 |
| 15yr BMI | ALSPAC | 4112 | 0.03 | 0.00 | 0.06 | 0.036 |
| 10yr FMI | ALSPAC | 3855 | 0.03 | -0.01 | 0.06 | 0.102 |
| 12yr FMI | ALSPAC | 3807 | 0.02 | -0.01 | 0.06 | 0.133 |
| 14yr FMI | ALSPAC | 3506 | 0.05 | 0.01 | 0.08 | 0.006 |
| 16yr FMI | ALSPAC | 2996 | 0.04 | 0.00 | 0.07 | 0.047 |
| 18yr FMI | ALSPAC | 2659 | 0.04 | 0.00 | 0.07 | 0.048 |

20 Genetic PCs were included as covariates. A fixed effects model was used for the meta-analysis

**Supplementary information S32: Exploring the change in MR estimates as SNP effect sizes vary**

Because the lassosum BMI PRS included many SNPs, it is likely that some of these SNPs had effects on the offspring outcomes via horizontal pleiotropic pathways (i.e. via exposures other than maternal BMI). Furthermore, most of the SNPs included in the lassosum BMI PRS had small effect sizes, and the consequences of this for the extent of horizontal pleiotropic effects are unclear (49). Horizontal pleiotropic effects could bias our MR estimates (for example if there is directional pleiotropy, in which pleiotropic effects that increase and decrease the offspring outcome do not cancel out). We conducted sensitivity analyses to explore the presence and likely direction of pleiotropic bias induced by including many SNPs with small effect sizes in the PRS.

We explored how MR estimates varied as the SNP effect sizes varied in ALSPAC. We divided the 80939 SNPs used to calculate the lassosum BMI PRS into 50 mutually exclusive bins based on absolute effect size (i.e. absolute lassosum SNP weights). We calculated 50 separate PRS from the SNPs in each bin, and used these PRS to calculate 50 MR estimates, before regressing the MR estimates on the SNP effect size bin (using HC1 robust SEs to account for heteroscedasticity). A non-zero slope in this regression would indicate that there is heterogeneity in the MR estimates for the different bins, suggesting that the MR estimates from at least some of the SNP bins are subject to bias due to horizontal pleiotropy, and by extension that the lassosum MR estimates from the primary analyses are subject to pleiotropic bias to some extent. The figures immediately below show the regression lines for each ALSPAC phenotype.

**Below, upper panel:** MR effect estimates for ***BW*** from SNPs binned by absolute lassosum effect size, regressed on SNP bin, in ALSPAC. Purple band indicates 95% CI for regression line, based on robust SEs. MR estimates are the mean increase in offspring phenotype (SD) per 1 SD increase in maternal pre-pregnancy BMI. **Lower panel:** IV *F*-statistic for the PRS from each bin

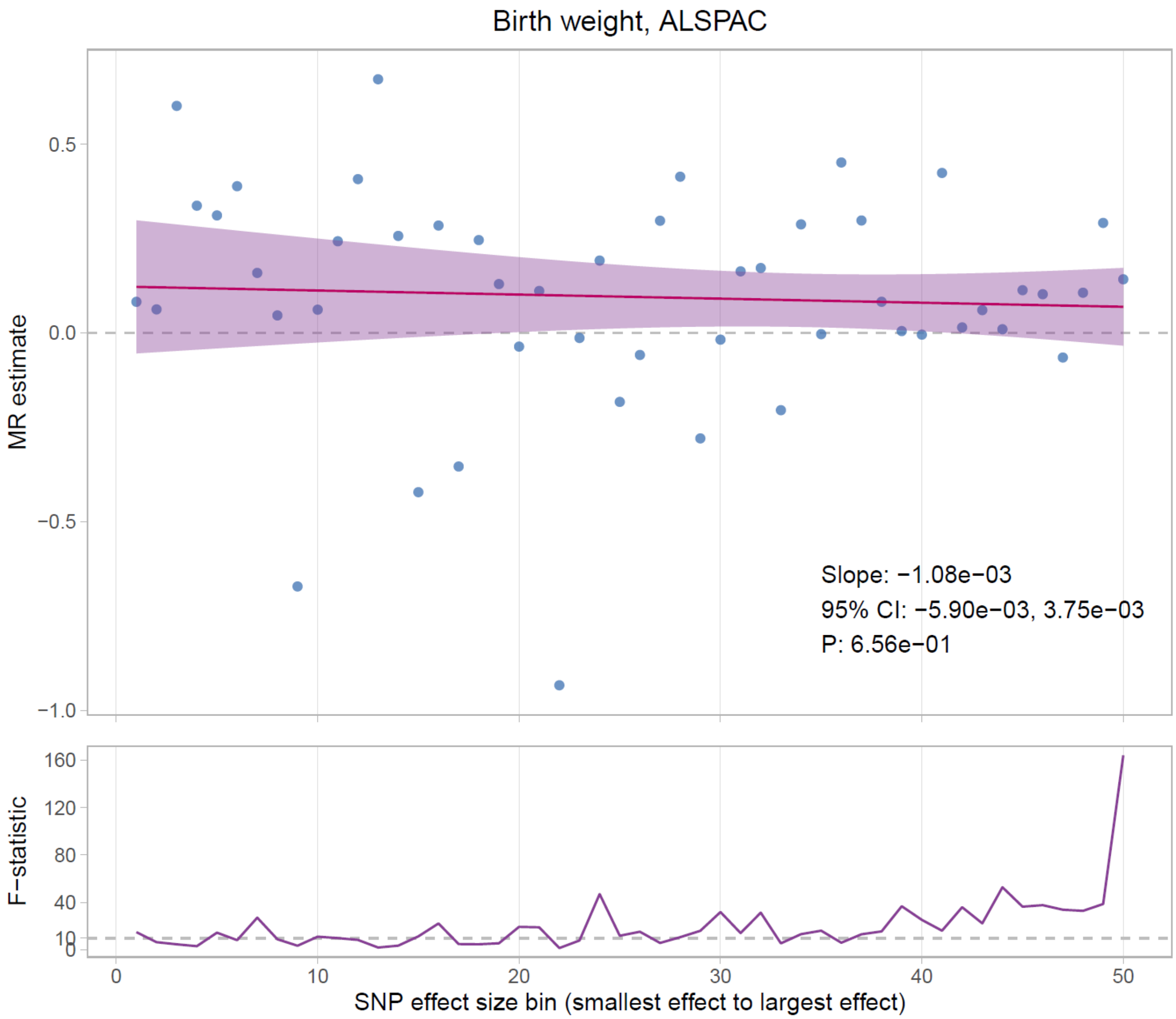

**Below, upper panel:** MR effect estimates for ***1 year BMI*** from SNPs binned by absolute lassosum effect size, regressed on SNP bin, in ALSPAC. Purple band indicates 95% CI for regression line, based on robust SEs. MR estimates are the mean increase in offspring phenotype (SD) per 1 SD increase in maternal pre-pregnancy BMI. **Lower panel:** IV *F*-statistic for the PRS from each bin

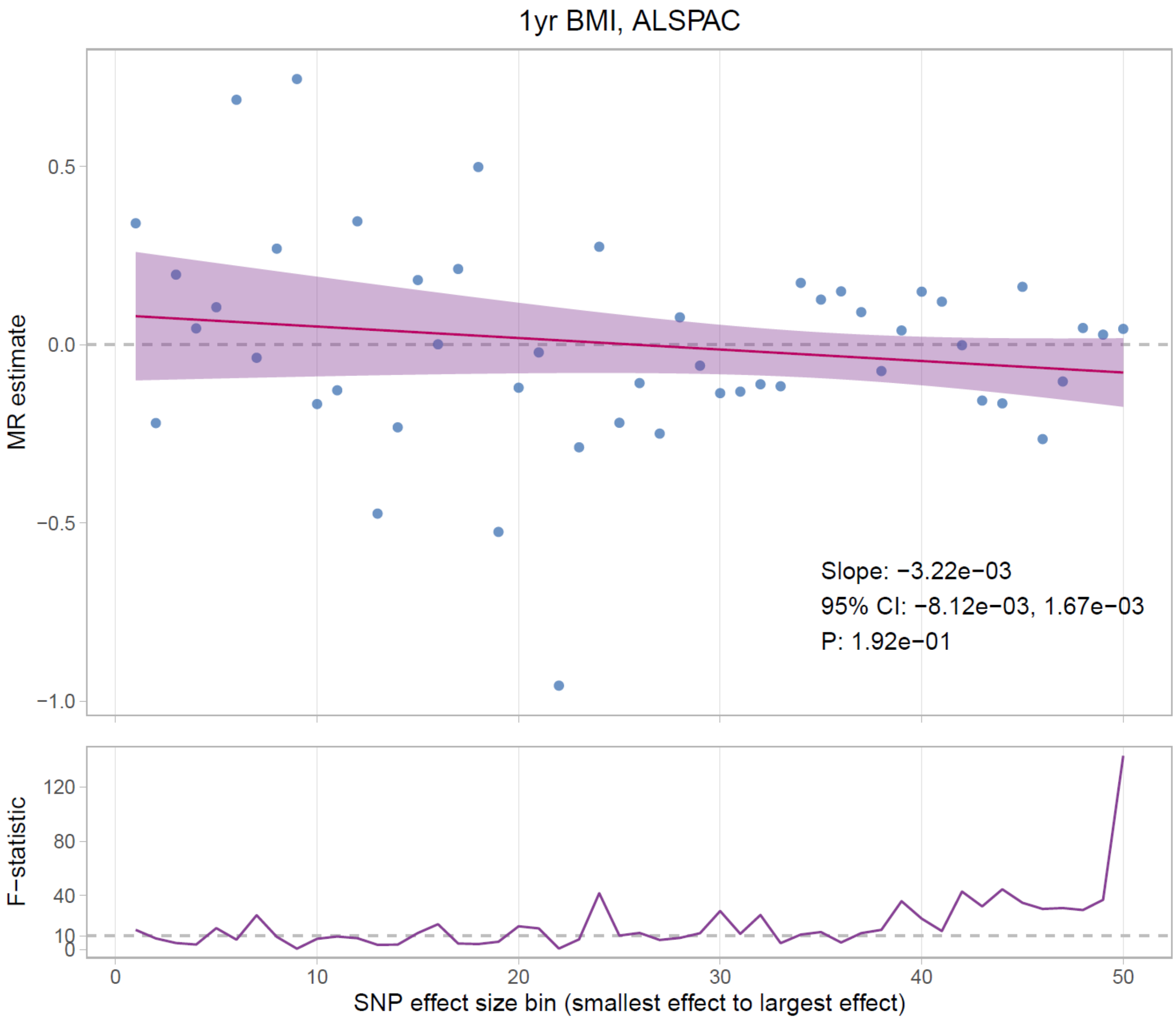

**Below, upper panel:** MR effect estimates for ***4 year BMI*** from SNPs binned by absolute lassosum effect size, regressed on SNP bin, in ALSPAC. Purple band indicates 95% CI for regression line, based on robust SEs. MR estimates are the mean increase in offspring phenotype (SD) per 1 SD increase in maternal pre-pregnancy BMI. **Lower panel:** IV *F*-statistic for the PRS from each bin

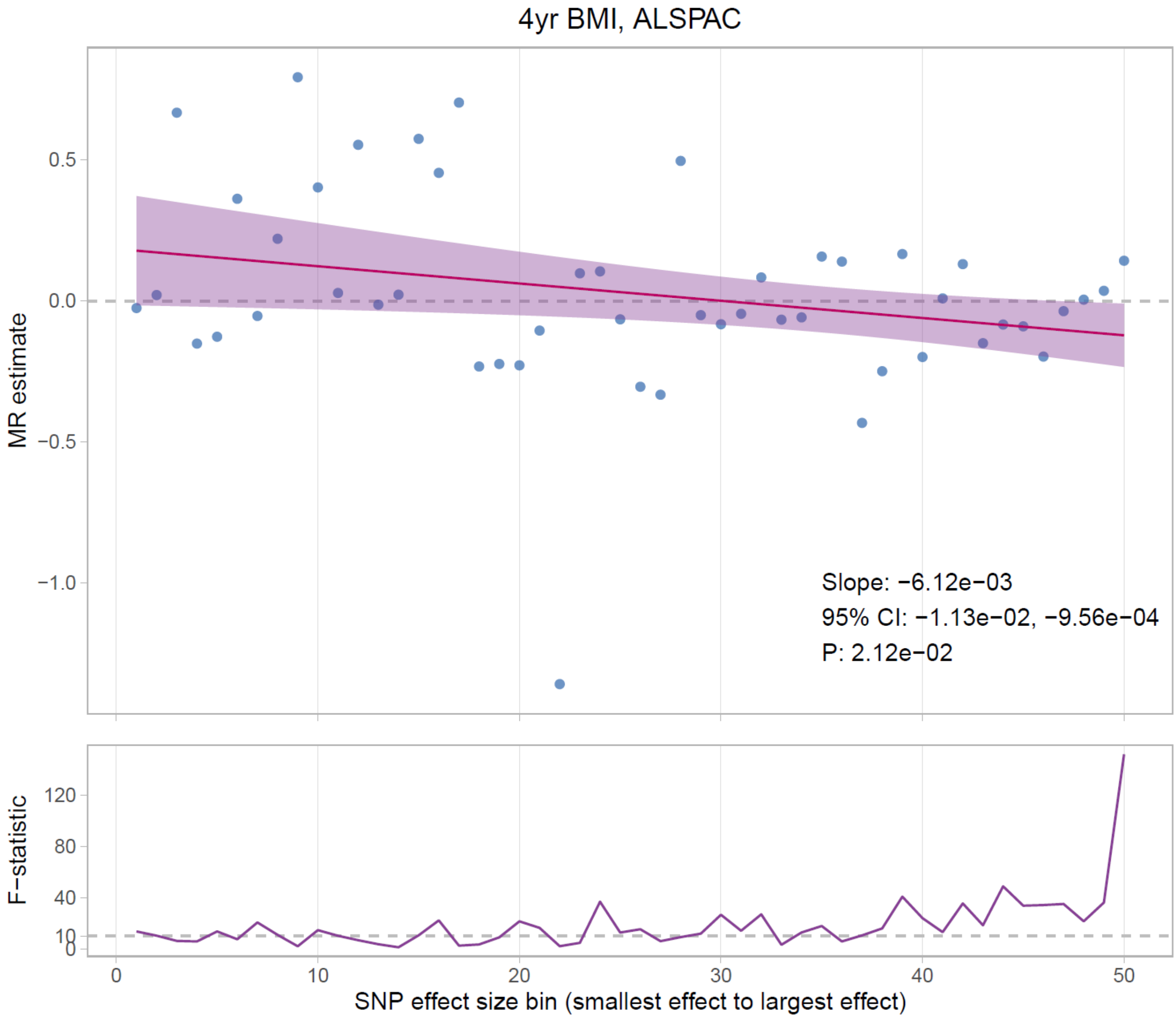

**Below, upper panel:** MR effect estimates for ***10 year BMI*** from SNPs binned by absolute lassosum effect size, regressed on SNP bin, in ALSPAC. Purple band indicates 95% CI for regression line, based on robust SEs. MR estimates are the mean increase in offspring phenotype (SD) per 1 SD increase in maternal pre-pregnancy BMI. **Lower panel:** IV *F*-statistic for the PRS from each bin

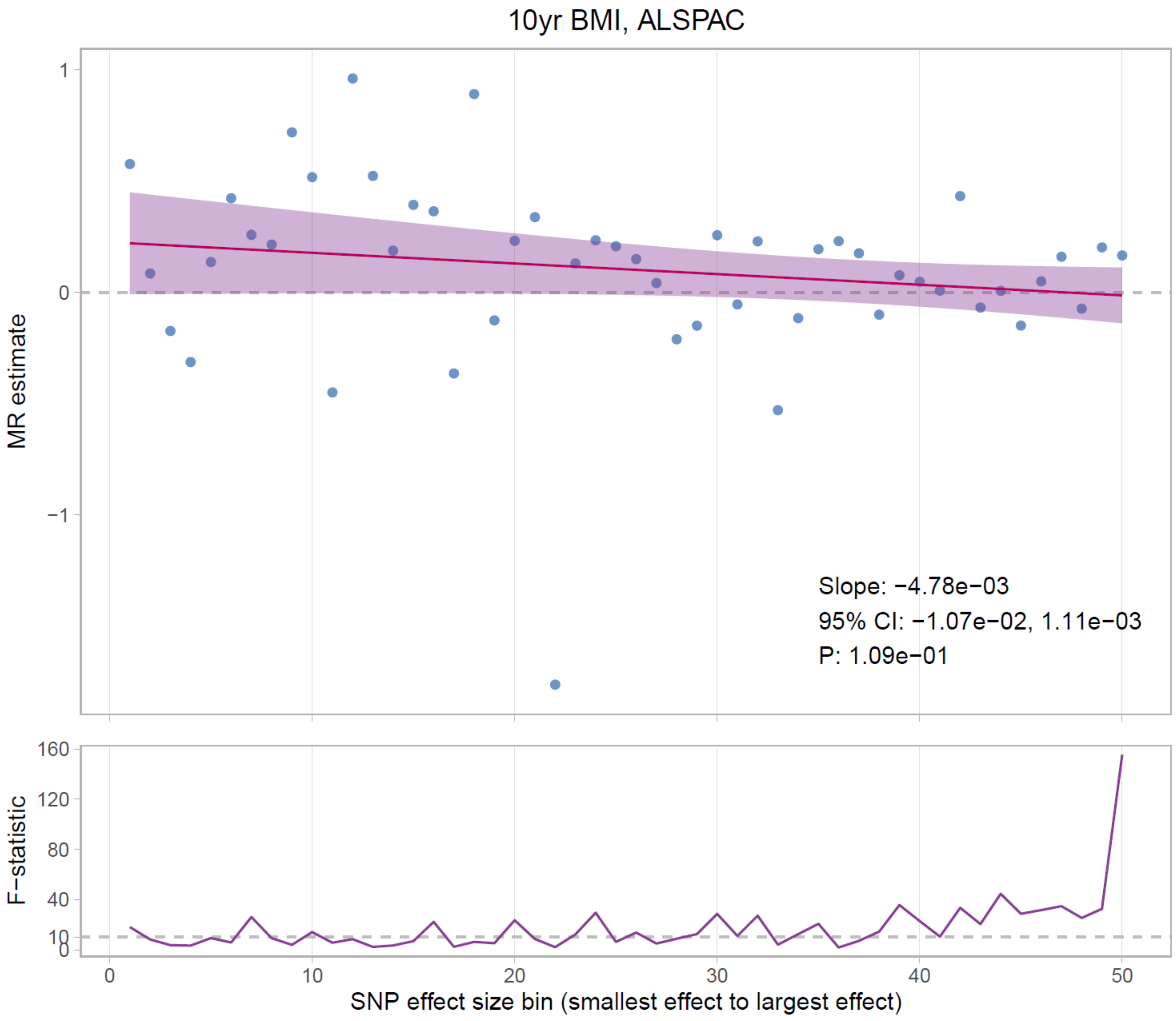

**Below, upper panel:** MR effect estimates for ***15 year BMI*** from SNPs binned by absolute lassosum effect size, regressed on SNP bin, in ALSPAC. Purple band indicates 95% CI for regression line, based on robust SEs. MR estimates are the mean increase in offspring phenotype (SD) per 1 SD increase in maternal pre-pregnancy BMI. **Lower panel:** IV *F*-statistic for the PRS from each bin

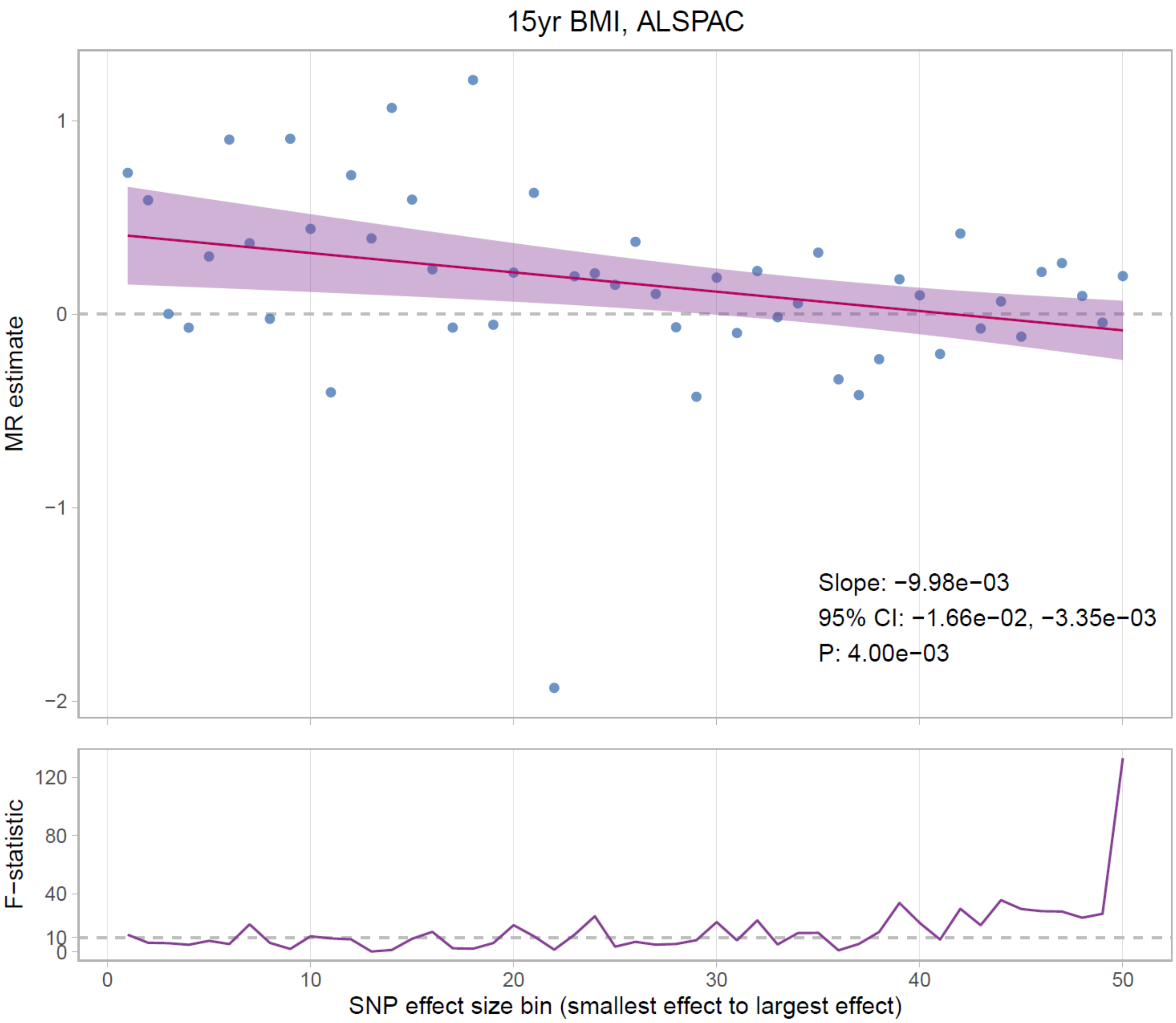

**Below, upper panel:** MR effect estimates for ***10 year FMI*** from SNPs binned by absolute lassosum effect size, regressed on SNP bin, in ALSPAC. Purple band indicates 95% CI for regression line, based on robust SEs. MR estimates are the mean increase in offspring phenotype (SD) per 1 SD increase in maternal pre-pregnancy BMI. **Lower panel:** IV *F*-statistic for the PRS from each bin

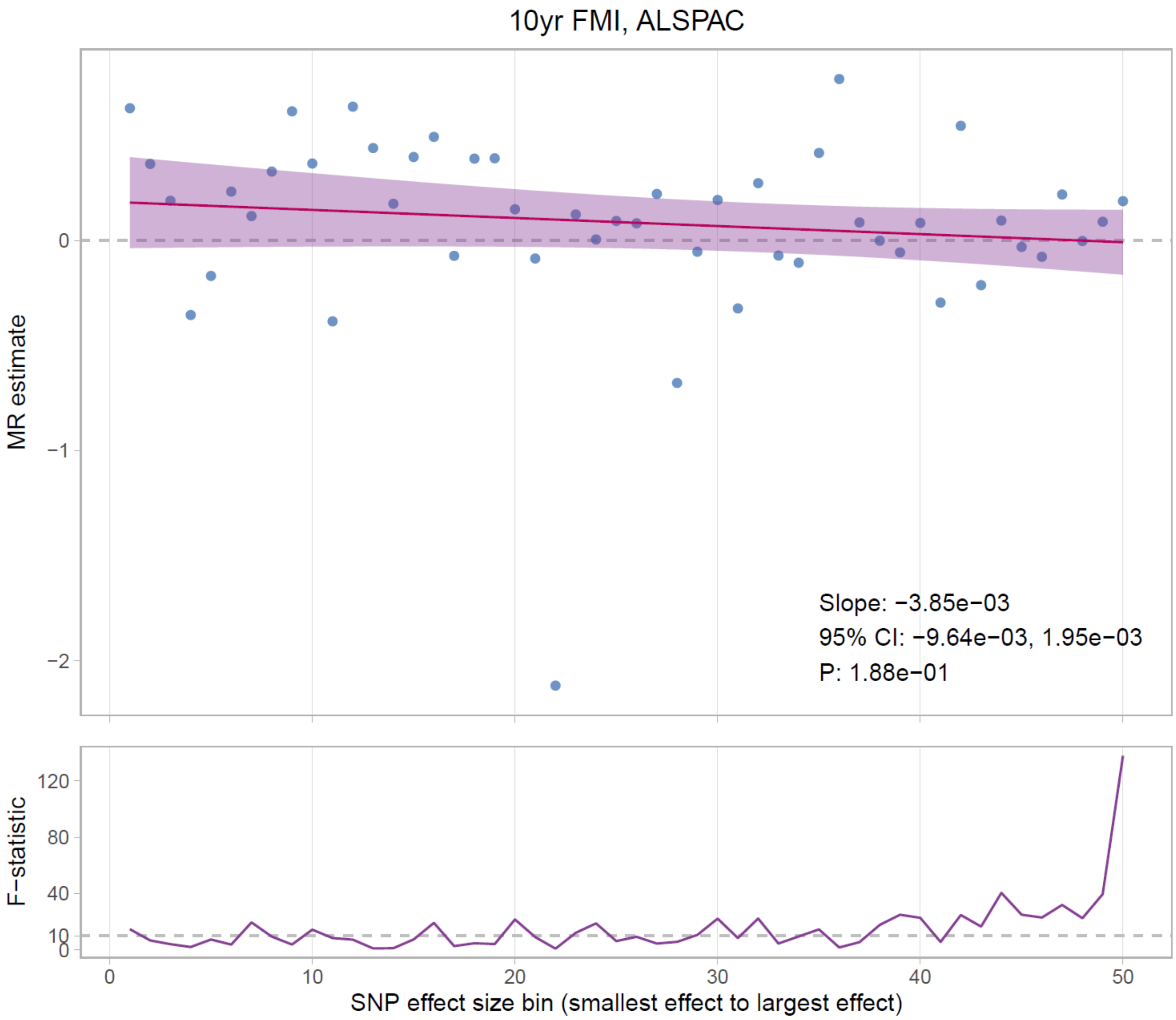

**Below, upper panel:** MR effect estimates for ***12 year FMI*** from SNPs binned by absolute lassosum effect size, regressed on SNP bin, in ALSPAC. Purple band indicates 95% CI for regression line, based on robust SEs. MR estimates are the mean increase in offspring phenotype (SD) per 1 SD increase in maternal pre-pregnancy BMI. **Lower panel:** IV *F*-statistic for the PRS from each bin

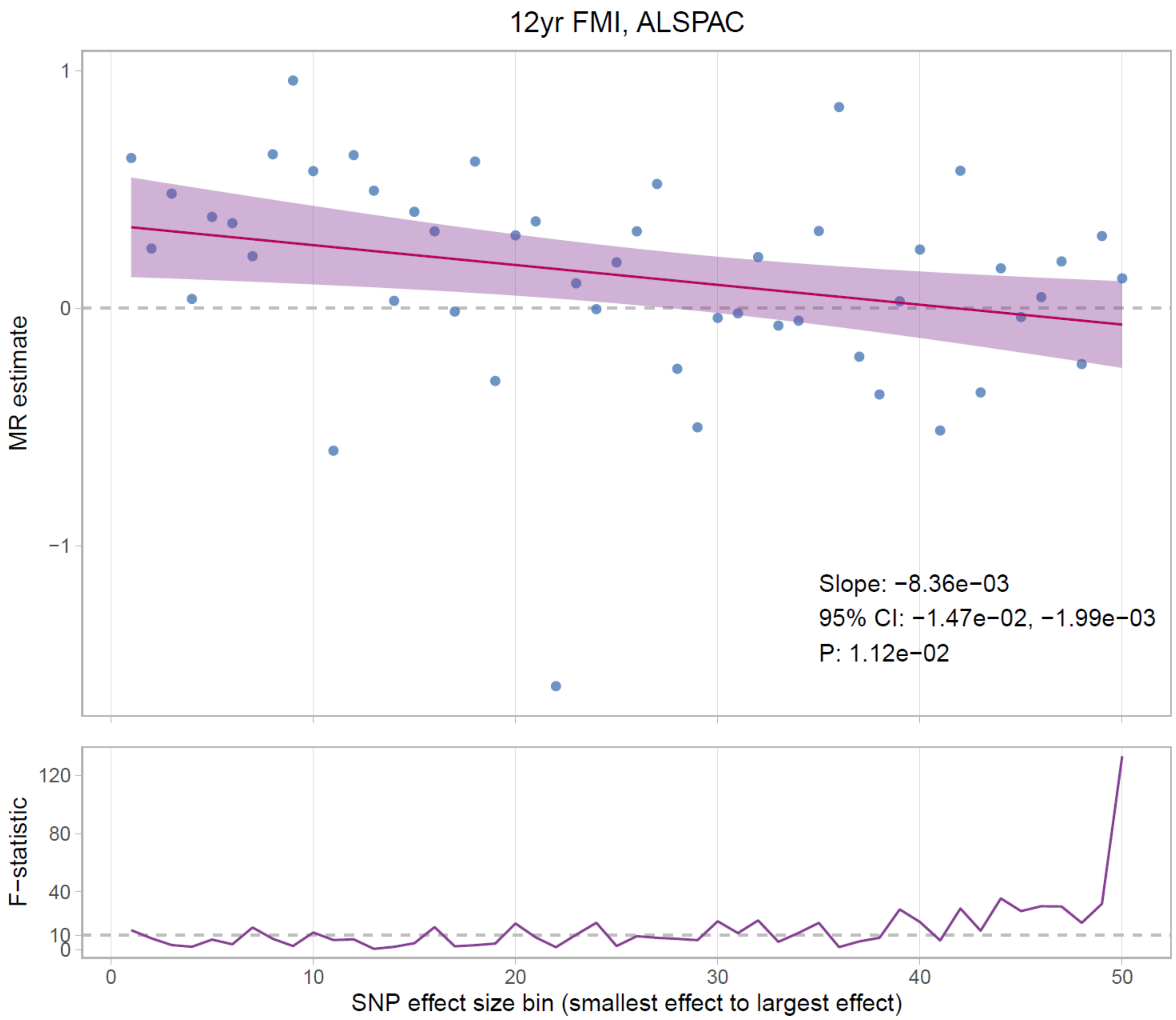

**Below, upper panel:** MR effect estimates for ***14 year FMI*** from SNPs binned by absolute lassosum effect size, regressed on SNP bin, in ALSPAC. Purple band indicates 95% CI for regression line, based on robust SEs. MR estimates are the mean increase in offspring phenotype (SD) per 1 SD increase in maternal pre-pregnancy BMI. **Lower panel:** IV *F*-statistic for the PRS from each bin

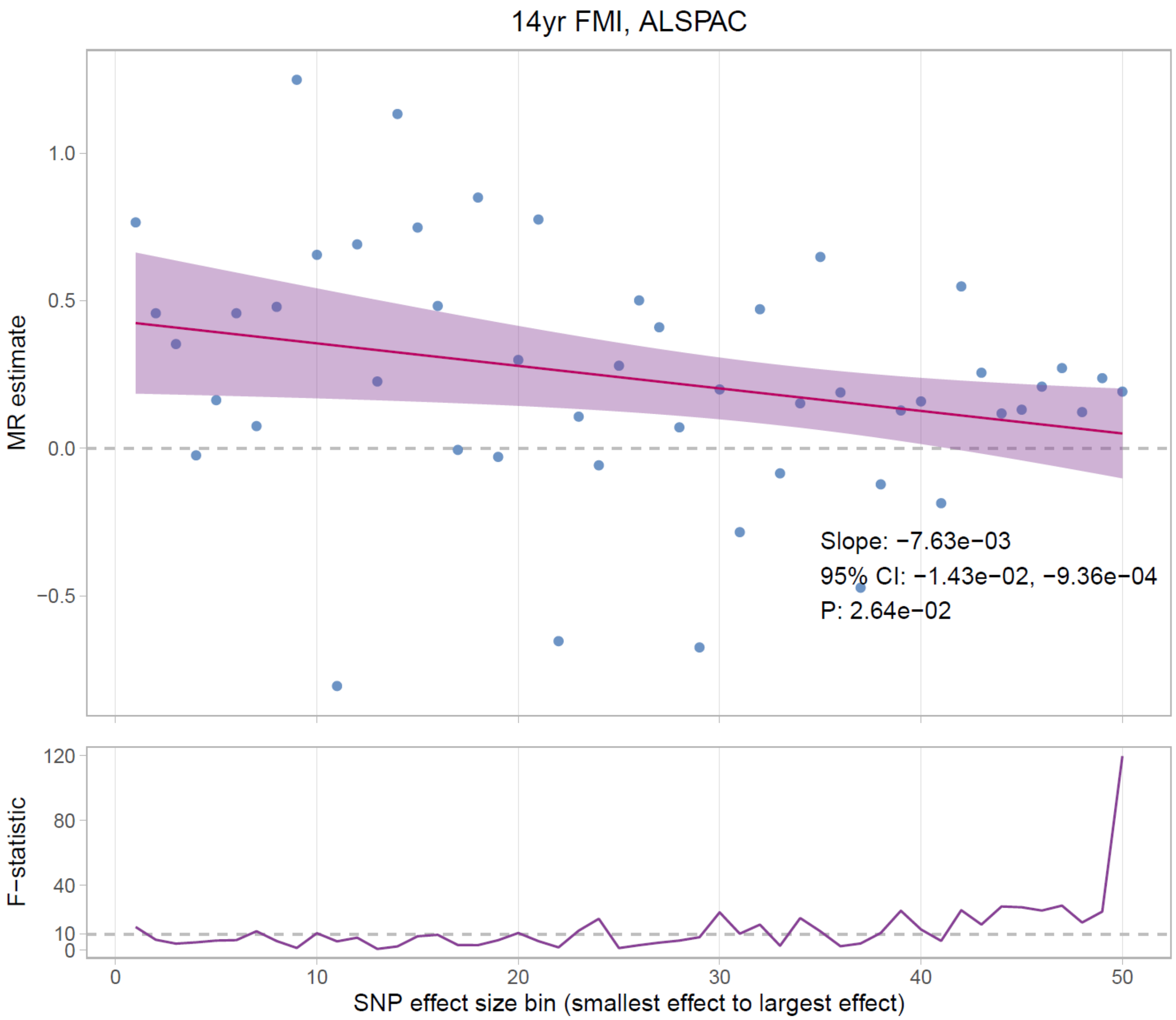

**Below, upper panel:** MR effect estimates for ***16 year FMI*** from SNPs binned by absolute lassosum effect size, regressed on SNP bin, in ALSPAC. Purple band indicates 95% CI for regression line, based on robust SEs. MR estimates are the mean increase in offspring phenotype (SD) per 1 SD increase in maternal pre-pregnancy BMI. **Lower panel:** IV *F*-statistic for the PRS from each bin

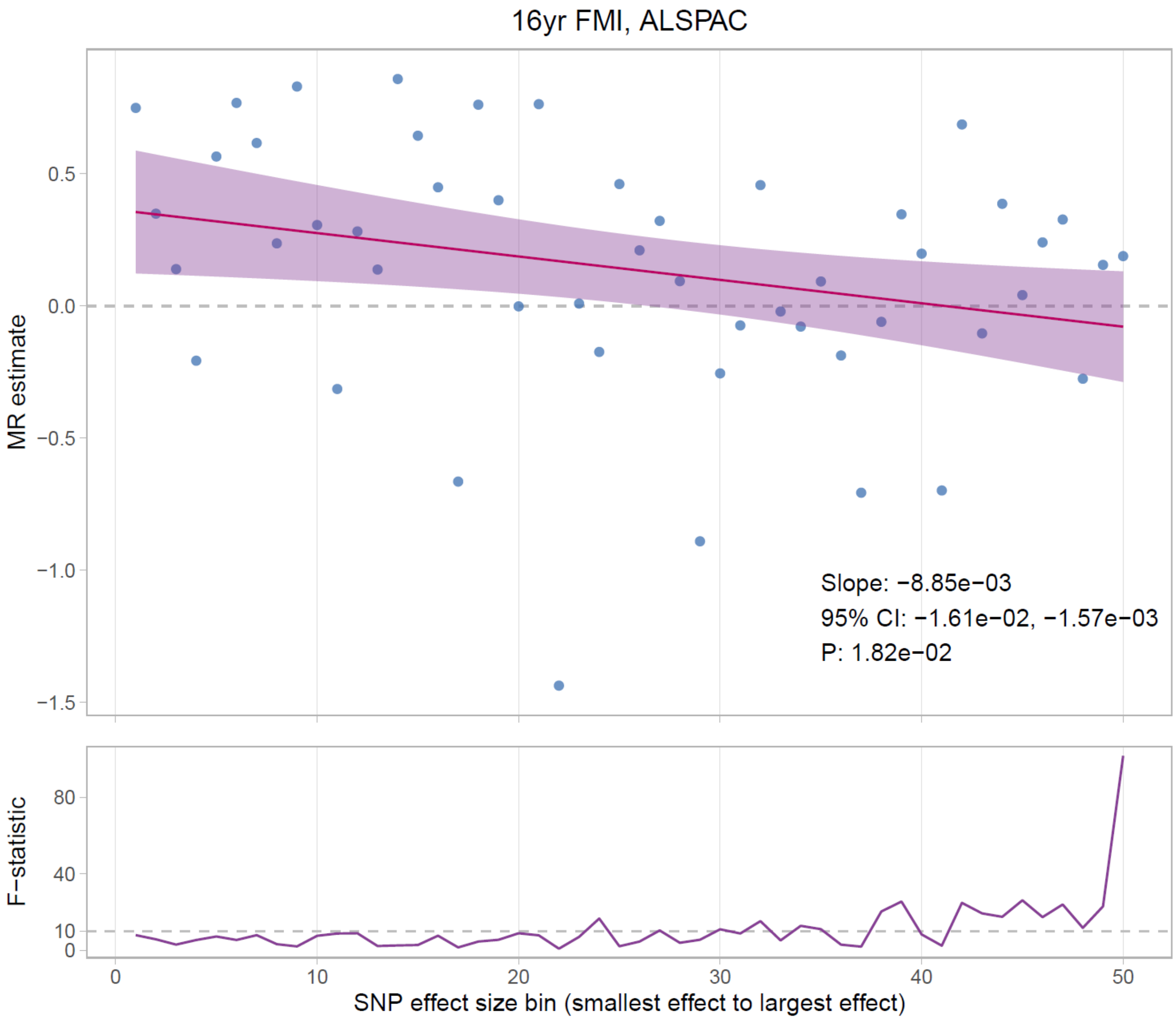

**Below, upper panel:** MR effect estimates for ***18 year FMI*** from SNPs binned by absolute lassosum effect size, regressed on SNP bin, in ALSPAC. Purple band indicates 95% CI for regression line, based on robust SEs. MR estimates are the mean increase in offspring phenotype (SD) per 1 SD increase in maternal pre-pregnancy BMI. **Lower panel:** IV *F*-statistic for the PRS from each bin

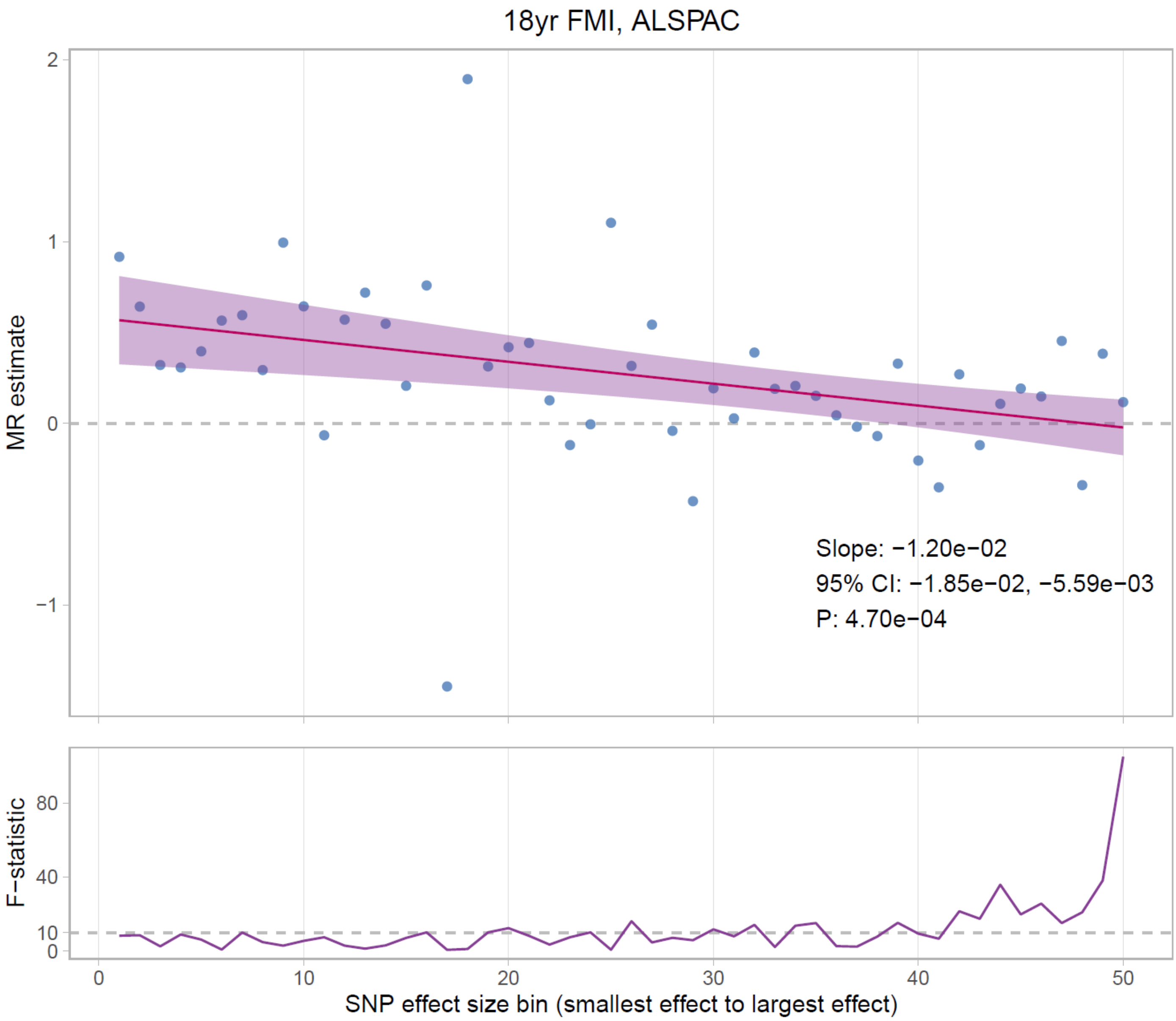

For the majority of outcomes (particularly in adolescence) there was strong statistical evidence that the regression line had a negative slope, indicating that SNPs with smaller effect sizes gave larger (more positive) MR estimates. An exception to this pattern was BW, for which the regression line was essentially flat. As SNP effect size became smaller the PRS became a weaker IV, as shown by the *F*-statistic plots in the lower panels of the figures. Because the present study used a one-sample MR design, weak instrument bias would bias MR estimates towards the phenotypic associations (50). We verified that the negative regression slopes observed were not driven by weak instrument bias by including the *F*-statistic as a covariate in the regression; results were similar and are available from the authors on request.

**Supplementary information S33: MR effect heterogeneity and MR Egger regression for large effect SNPs in ALSPAC**

The observed relationship between SNP effect size and MR estimates (**Supplementary information S32**) suggests that the degree of pleiotropic bias changes with SNP effect size. It is therefore plausible that SNPs with either small or large effect sizes produce biased MR estimates; however, based on the regression results above (**Supplementary information S32**) it is not possible to determine whether the bias primarily affects small or large effect size SNPs. We hypothesised that the true causal effect of maternal BMI on offspring adolescent adiposity is close to zero, and accordingly that MR estimates from large effect SNPs are unbiased (i.e. SNP bins at the right of the plots immediately above, and MR estimates from GWS SNPs in the primary analyses). This would imply that MR estimates from small effect SNPs are positively biased (i.e. SNP bins at the left of the plots immediately above, and MR estimates from the lassosum PRS in the primary analyses). We tested for evidence of pleiotropic bias for large effect size SNPs using a two-sample MR approach, in which the SNP-exposure associations came from our previous UKB BMI GWAS (excluding participants attending the Bristol assessment centre, as described in **Supplementary information S11**), and the SNP-outcome associations were estimated in ALSPAC using a non-transmitted allele approach as per our primary analyses. We tested for evidence of between-SNP MR estimate heterogeneity (Cochran's *Q* test) and estimated the MR Egger regression intercept (51); results are given in the table immediately below.

| SNP set | Outcome | Inverse variance weighted MR | | | | MR Egger | | | | | |
|---|---|---|---|---|---|---|---|---|---|---|---|
| | | | | | | Slope | | | Intercept | | |
| | | Beta | SE | *P* | $P_{het}$[a] | Beta | SE | *P* | Beta | SE | *P* |
| Locke | BW | 0.156 | 0.152 | 0.304 | 0.017* | 0.373 | 0.336 | 0.270 | -0.006 | 0.008 | 0.472 |
| | 1yr BMI | 0.320 | 0.141 | 0.023 | 0.261 | 0.324 | 0.312 | 0.302 | 0.000 | 0.008 | 0.990 |
| | 4yr BMI | 0.192 | 0.138 | 0.164 | 0.661 | 0.371 | 0.303 | 0.224 | -0.005 | 0.007 | 0.508 |
| | 10yr BMI | 0.069 | 0.152 | 0.652 | 0.124 | 0.173 | 0.337 | 0.608 | -0.003 | 0.008 | 0.728 |
| | 15yr BMI | -0.146 | 0.149 | 0.328 | 0.346 | -0.248 | 0.329 | 0.453 | 0.003 | 0.008 | 0.728 |
| | 10yr FMI | 0.234 | 0.162 | 0.150 | 0.180 | 0.538 | 0.357 | 0.136 | -0.008 | 0.009 | 0.342 |
| | 12yr FMI | 0.015 | 0.168 | 0.928 | 0.078 | -0.040 | 0.371 | 0.915 | 0.002 | 0.009 | 0.868 |
| | 14yr FMI | -0.063 | 0.159 | 0.690 | 0.569 | 0.034 | 0.349 | 0.922 | -0.003 | 0.009 | 0.754 |
| | 16yr FMI | -0.108 | 0.180 | 0.549 | 0.204 | -0.266 | 0.398 | 0.506 | 0.004 | 0.010 | 0.657 |
| | 18yr FMI | -0.132 | 0.181 | 0.468 | 0.448 | 0.479 | 0.396 | 0.230 | -0.017 | 0.010 | 0.087 |
| Yengo | BW | 0.158 | 0.082 | 0.054 | 0.435 | 0.351 | 0.219 | 0.109 | -0.003 | 0.004 | 0.342 |
| | 1yr BMI | 0.116 | 0.085 | 0.173 | 0.304 | 0.334 | 0.228 | 0.143 | -0.004 | 0.004 | 0.302 |
| | 4yr BMI | 0.102 | 0.088 | 0.245 | 0.269 | 0.319 | 0.234 | 0.173 | -0.004 | 0.004 | 0.316 |
| | 10yr BMI | 0.085 | 0.088 | 0.330 | 0.976 | -0.125 | 0.233 | 0.593 | 0.004 | 0.004 | 0.332 |
| | 15yr BMI | 0.113 | 0.090 | 0.210 | 0.918 | -0.398 | 0.241 | 0.100 | 0.009 | 0.004 | 0.023* |
| | 10yr FMI | 0.093 | 0.095 | 0.326 | 0.984 | 0.191 | 0.253 | 0.452 | -0.002 | 0.004 | 0.678 |
| | 12yr FMI | 0.096 | 0.095 | 0.311 | 0.951 | -0.098 | 0.252 | 0.698 | 0.003 | 0.004 | 0.407 |
| | 14yr FMI | 0.161 | 0.099 | 0.103 | 0.806 | -0.237 | 0.264 | 0.370 | 0.007 | 0.004 | 0.104 |
| | 16yr FMI | 0.094 | 0.107 | 0.383 | 0.323 | -0.438 | 0.286 | 0.127 | 0.009 | 0.005 | 0.046* |
| | 18yr FMI | 0.090 | 0.112 | 0.422 | 0.680 | -0.004 | 0.300 | 0.990 | 0.002 | 0.005 | 0.735 |

All MR estimates are for the effect of maternal BMI on the offspring's age and sex standardised outcomes. **a**: *P*-value for heterogeneity of MR estimates between SNPs (Cochran's *Q*), ***P** <0.05, **Locke**: 87 clumped GWS BMI-associated SNPs from Locke *et al*. (37), **Yengo**: 497 clumped GWS BMI-associated SNPs from Yengo *et al*. (38). Clumping was carried out using ALSPAC as the LD reference panel

In general there was not strong statistical evidence for between-SNP MR estimate heterogeneity, nor was there strong evidence that the MR-Egger intercept differed from zero. Taken together, this suggests a lack of pleiotropic bias for the large effect size SNPs, and in combination with the regression results from **Supplementary information S32** suggests that MR estimates using small effect SNPs (including our primary lassosum MR estimates) may be positively biased. Importantly, positive pleiotropic bias would weaken the apparent evidence that the MR estimates differ from the MV estimates (i.e. the bias would cause $P_{dif}$ to be bigger). As such, the evidence supporting our conclusion that observational associations between maternal BMI and offspring adolescent BMI are subject to residual confounding (from our primary lassosum MR analyses) is likely to be conservative.

#### Supplementary information S34: Power calculations

The primary analysis presented in Richmond *et al*. 2017 had 82% power to detect a true causal effect of size 0.28 (equal to their observed MV estimate, controlled for age and sex), with two-sided α = 0.05 (**large causal effect** below). However for a true causal effect of size 0.16 (**moderate causal effect** below) there was only 37% power. The present study had 79% power to detect a true causal effect of size 0.16 (for **15yr BMI** below), and had 79% power to detect a true causal effect of size 0.13 (for **4yr BMI** below). **MV**: confounder adjusted multivariable linear regression estimate. All power calculations were carried out using the online tool by Brion *et al.* (52).

| | **Richmond *et al.* 2017** | | **Present study** | |
|---|---|---|---|---|
| | **Large causal effect** | **Moderate causal effect** | **15yr BMI, ALSPAC** | **4yr BMI, ALSPAC and BiB meta-analysis** |
| Sample size | 6057 | 6057 | 4112 | 7575 |
| IV $R^2$ | 1.6% | 1.6% | 6.6% | 5.8% |
| α | 0.05 | 0.05 | 0.05 | 0.05 |
| Variance of exposure | 1 | 1 | 1 | 1 |
| Variance of outcome | 1 | 1 | 1 | 1 |
| MV effect size | 0.28 | 0.28 | 0.32 | 0.18 |
| True causal effect size | 0.28 | 0.16 | 0.16 | 0.13 |
| Power | 0.82 | 0.37 | 0.79 | 0.79 |

**Supplementary information S35: Comparison to previous analyses of ALSPAC data**

Two previous papers have used MR to investigate the causal effect of maternal pre-pregnancy BMI on offspring child/adolescent adiposity using ALSPAC data (7, 43), and one other methodological paper presented some limited MR analyses as an empirical example (22). These prior analyses of ALSPAC data are compared with the present work in the table immediately below.

| | **Lawlor *et al.* 2008 (43)** | **Richmond *et al.* 2017 (7)** | **Lawlor *et al.* 2017 (22)** | **Present study** |
|---|---|---|---|---|
| Exposure | Maternal pre-pregnancy BMI | Maternal pre-pregnancy BMI | Maternal pre-pregnancy BMI | Maternal pre-pregnancy BMI |
| Relevant outcomes (*N* in ALSPAC analysis) | | | | Birth weight (5085)<br>1yr BMI (4838)<br>4yr BMI (4670) |
| | | 7yr BMI (3720) | | |
| | 9yr fat mass[a] (3263) | | | |
| | | 10yr BMI (3657) | | 10yr BMI (4476[b]) |
| | 11yr fat mass[a] (3263) | | | |
| | | 12yr BMI (3496) | | |
| | | 14yr BMI (3227) | | |
| | | | | 15yr BMI (4112[b]) |
| | | 16yr BMI (2806) | | |
| | | 18yr BMI (2521) | 18yr BMI (2482) | |
| | | 10yr FMI[a] (3495) | | 10yr FMI[a] (3855[c]) |
| | | 12yr FMI[a] (3444) | | 12yr FMI[a] (3807[c]) |
| | | 14yr FMI[a] (3192) | | 14yr FMI[a] (3506[c]) |
| | | 16yr FMI[a] (2715) | | 16yr FMI[a] (2996[c]) |
| | | 18yr FMI[a] (2430) | 18yr FMI[a] (2392) | 18yr FMI[a] (2659[c]) |
| IVs | rs9939609 at the *FTO* locus | 31 SNP BMI PRS<br>97 SNP BMI PRS | 97 SNP BMI PRS | rs9939609<br>31 SNP BMI PRS<br>87[d] SNP BMI PRS<br>497 SNP BMI PRS<br>80939 SNP BMI PRS |
| Main analysis method | One sample MR, maternal genotype was adjusted for offspring genotype | One sample MR, maternal PRS was adjusted for offspring PRS | One sample MR, maternal non-transmitted allele PRS (presented as an empirical example in a primarily methodological paper, with few sensitivity analyses) | One sample MR, maternal non-transmitted allele PRS |
| Other cohorts analysed | None | Generation R | None | Born in Bradford |

**a**: Fat mass and fat mass index (FMI) were assessed by dual-energy X-ray absorptiometry (DXA) scan, **b**: The larger sample size for BMI outcomes in the present study versus Richmond *et al.* 2017 is due to (i) the lack of exclusion of cryptic relatedness in the present study (results were similar in sensitivity analyses with removal of cryptic relatedness), (ii) inclusion of BMI measurements from questionnaire data (**Supplementary information S6**), and (iii) inclusion of measurements from broader age windows (**Supplementary information S5**) in order to facilitate comparisons to previous work in which we took a similar approach (6), **c**: The larger sample size for FMI outcomes in the present study versus Richmond *et al*. 2017 and Lawlor *et al*. 2017 was due to the lack of exclusion of cryptic relatedness in the present study, **d**: Fewer BMI-associated SNPs were used in the present study versus Richmond *et al*. 2017 and Lawlor *et al*. 2017 because in the present study we (i) excluded three SNPs associated with BMI in men only in the Locke *et al*. GWAS, and (ii) applied more stringent filtering on SNP imputation quality score ($r^2$)
